# Inhaled budesonide for COVID-19 in people at higher risk of adverse outcomes in the community: interim analyses from the PRINCIPLE trial

**DOI:** 10.1101/2021.04.10.21254672

**Authors:** PRINCIPLE Collaborative Group, Ly-Mee Yu, Mona Bafadhel, Jienchi Dorward, Gail Hayward, Benjamin R Saville, Oghenekome Gbinigie, Oliver Van Hecke, Emma Ogburn, Philip H Evans, Nicholas PB Thomas, Mahendra G Patel, Nicholas Berry, Michelle A. Detry, Christina T. Saunders, Mark Fitzgerald, Victoria Harris, Simon de Lusignan, Monique I Andersson, Peter J Barnes, Richard EK Russell, Dan V Nicolau, Sanjay Ramakrishnan, FD Richard Hobbs, Christopher C Butler

## Abstract

**BACKGROUND:** Inhaled budesonide has shown efficacy for treating COVID-19 in the community but has not yet been tested in effectiveness trials.

**METHODS:** We performed a multicenter, open-label, multi-arm, adaptive platform randomized controlled trial involving people aged ≥65 years, or ≥50 years with comorbidities, and unwell ≤14 days with suspected COVID-19 in the community (PRINCIPLE). Participants were randomized to usual care, usual care plus inhaled budesonide (800µg twice daily for 14 days), or usual care plus other interventions. The co-primary endpoints are time to first self-reported recovery, and hospitalization/death related to COVID-19, both measured over 28 days from randomisation and analysed using Bayesian models.

**RESULTS:** The trial opened on April 2, 2020. Randomization to inhaled budesonide began on November 27, 2020 and was stopped on March 31, 2021 based on an interim analysis using data from March 4, 2021. Here, we report updated interim analysis data from March 25, 2021, at which point the trial had randomized 4663 participants with suspected COVID-19. Of these, 2617 (56.1%) tested SARS-CoV-2 positive and contributed data to this interim budesonide primary analysis; 751 budesonide, 1028 usual care and 643 to other interventions. Time to first self-reported recovery was shorter in the budesonide group compared to usual care (hazard ratio 1.208 [95% BCI 1.076 – 1.356], probability of superiority 0.999, estimated benefit [95% BCI] of 3.011 [1.134 – 5.41] days). Among those in the interim budesonide primary analysis who had the opportunity to contribute data for 28 days follow up, there were 59/692 (8.5%) COVID-19 related hospitalizations/deaths in the budesonide group vs 100/968 (10.3%) in the usual care group (estimated percentage benefit, 2.1% [95% BCI −0.7% – 4.8%], probability of superiority 0.928).

**CONCLUSIONS:** In this updated interim analysis, inhaled budesonide reduced time to recovery by a median of 3 days in people with COVID-19 with risk factors for adverse outcomes. Once 28 day follow up is complete for all participants randomized to budesonide, final analyses of time to recovery and hospitalization/death will be published. (Funded by the National Institute of Health Research/ United Kingdom Research Innovation [MC_PC_19079]; PRINCIPLE ISRCTN number, <u>ISRCTN86534580</u>.)

## INTRODUCTION

There is an urgent need for effective and safe community-based treatments for coronavirus disease 2019 (COVID-19), especially for older people and those with co-morbidities who are at higher risk of hospitalization and death.^1^

Inhaled corticosteroids are widely available, inexpensive and generally safe, and have been proposed as a COVID-19 treatment due to their targeted anti-inflammatory effects in the lungs.^2,3^ In animal and human studies, inhaled corticosteroids reduce expression of ACE-2 and TMPRSS2,^4,5^ which are used by SARS-CoV-2 for cell entry.^6^ Early in the pandemic, the low prevalence of asthma and chronic obstructive pulmonary disease (COPD) among people hospitalized with COVID-19, lead to speculation that inhaled corticosteroids used to treat these conditions may be protective.^2,7^ Furthermore, systemic corticosteroids reduce deaths in hospitalized patients with COVID-19^8,9^ likely because the hyperinflammatory state is responsible for the subsequent damage from SARS-CoV-2 infection.^10,11^ Two large, population-based studies in primary care in the United Kingdom found an increased risk of COVID-19 hospitalization and/or death among people prescribed inhaled corticosteroids,^12,13^ although residual confounding by unmeasured disease severity could not be ruled out. An efficacy trial including 146 adults with mild COVID-19 in the community found inhaled budesonide reduced COVID-19 related emergency assessments or hospitalizations.^14^ However, thus far, there are no results reported from large effectiveness trials of inhaled budesonide for COVID-19.

We therefore aimed to determine whether inhaled budesonide speeds recovery or reduces hospital admission or death from COVID-19 in people at higher risk of an adverse outcome in the community.

## METHODS

### TRIAL DESIGN AND OVERSIGHT

We assessed the effectiveness of inhaled budesonide in the UK national, multi-center, primary care, open-label, multi-arm, prospective adaptive Platform Randomised trial of INterventions against COVID-19 In older peoPLE (PRINCIPLE), which opened on April 2, 2020, and is ongoing. The protocol is available as a supplement to this manuscript, and at www.principletrial.org. A “platform trial” allows multiple treatments for the same disease to be tested simultaneously. A master protocol defines prospective decision criteria for dropping interventions for futility, declaring interventions superior, or adding new interventions.^15^ Interventions under evaluation in PRINCIPLE have included hydroxychloroquine, azithromycin,^16^ doxycycline,^17^ colchicine, and inhaled budesonide reported here with interim results.

The UK Medicines and Healthcare products Regulatory Agency and the South Central-Berkshire Research Ethics Committee (Ref: 20/SC/0158) approved the trial protocol and all recruitment processes. Online consent is obtained from all participants. The authors vouch for the accuracy and completeness of the data and for fidelity to the protocol. An independent Trial Steering Committee and Data Monitoring and Safety Committee provide trial oversight.

### PARTICIPANTS

People in the community were eligible if they were aged ≥65 years, or ≥50 years with comorbidities (see trial protocol), and had ongoing symptoms from polymerase chain reaction (PCR) confirmed or suspected COVID-19 (in accordance with the United Kingdom National Health Service definition of high temperature and/or new, continuous cough and/or change in sense of smell/taste),^18,19^ which started within the past 14 days. People were ineligible to be randomized to budesonide if they were already taking inhaled or systemic corticosteroids, were unable to use an inhaler, or if inhaled budesonide was contraindicated.^20^ Initially, eligible people were recruited, screened and enrolled through participating general medical practices, but from May 17, 2020, people across the UK could enroll online or by telephone. We undertook extensive community outreach to increase recruitment from ethnic minority and socially deprived communities, as these groups have been disproportionally affected by COVID-19.

### TRIAL PROCEDURES

Eligible, consenting participants were randomized using a secure, in-house, web-based randomization system (Sortition). When the budesonide group opened on November 27, 2020, the azithromycin, doxycycline and usual care groups were also active. Randomization probabilities were determined using response adaptive randomization via regular interim analyses, which allows allocation of more participants to interventions with better observed outcomes (see Adaptive Design Report). The azithromycin and doxycycline groups had stopped by December 14, 2020, at which point 1:1 allocation between usual care and budesonide occurred, stratified by age, and comorbidity. On March 04, 2021 the colchicine arm opened, with subsequent response adaptive randomization.

Participants were followed up through an online, daily symptom diary for 28 days after randomization, supplemented with telephone calls on days 7, 14 and 28. Participants were encouraged to nominate a trial partner to help provide follow up data. We obtained consent to ascertain healthcare use outcome data from general practice and hospital records. We aimed to provide a self-swab for SARS-CoV-2 confirmatory PCR testing, but capacity issues early in the pandemic meant testing was unavailable for some participants.

### TRIAL INTERVENTIONS

Participants received usual care plus inhaled budesonide 800µg twice daily for 14 days (Pulmicort Turbohaler, AstraZeneca), or usual care alone. This breath-actuated inhaler was chosen due to its ease of use for unwell, co-morbid, and potentially frail and older patients, and was either issued by the participant’s general medical practitioner (GP), or centrally by the study team and delivered to the participant. Participants in the budesonide arm were also sent a link to a video demonstrating current inhaler use, with further explanation available by telephone support. Usual care in the United Kingdom National Health Service for suspected COVID-19 in the community is largely focused on managing symptoms.^21^

### PRIMARY OUTCOMES

The trial commenced with the primary outcome of hospitalization or death within 28 days. However, the proportion requiring hospitalization in the UK^22^ was lower than initially expected^23^. Therefore, the Trial Management Group and Trial Steering Committee recommended amending the primary outcome to include a measure of illness duration.^24,25^ Duration of illness is an important outcome for patients and has important economic and social impacts. This received ethical approval on September 16, 2020, and was implemented before performing any interim analyses. Thus, the trial has two co-primary endpoints measured within 28 days of randomization: 1) time to first reported recovery defined as the first instance that a participant reports feeling recovered; and 2) hospitalization or death related to COVID-19.

### SECONDARY OUTCOMES

Secondary outcomes include a rating of how well participants feel (“How well are you feeling today? Please rate how you are feeling now using a scale of 1 – 10, where 1 is the worst you can imagine, and 10 is feeling the best you can imagine”), time to sustained recovery (date participant first reports feeling recovered and subsequently remains well until 28 days), binary outcome of early sustained recovery (reports feeling recovered within the first 14 days from randomization and remains recovered until day 28), time to initial alleviation of symptoms (date participant first reports all symptoms as minor or none), time to sustained alleviation of symptoms, time to initial reduction of severity of symptoms, contacts with health services, hospital assessment without admission, oxygen administration, Intensive Care Unit admission. mechanical ventilation, adherence to study treatment and the WHO-5 Well-Being Index^26^. We included secondary outcomes capturing sustained recovery due to the often recurrent nature of COVID-19 symptoms.

### STATISTICAL ANALYSIS

Sample size calculation and statistical analysis are detailed in the Adaptive Design Report and the Master Statistical Analysis Plan. We justify sample sizes by simulating the operating characteristics of the adaptive design in multiple scenarios, which explicitly account for response adaptive randomization, early stopping for futility/success and multiple interventions. In brief, for the primary outcome analyses, assuming a median time to recovery of nine days in the usual care group, approximately 400 participants per group would provide 90% power to detect a 2-day difference in median recovery time. Assuming 5% hospitalization in the usual care group, approximately 1500 participants per group would provide 90% power to detect a 50% reduction in the relative risk of hospitalization/death.

The first co-primary outcome, time to first self-reported recovery, was analyzed using a Bayesian piecewise exponential model regressed on treatment and stratification covariates, and included parameters for temporal drift. The second co-primary outcome, hospitalization/death, was analyzed using a Bayesian logistic regression model regressed on treatment and stratification covariates. The primary outcomes were evaluated using a “gate-keeping” strategy to preserve the overall Type I error of the primary endpoints without additional adjustments for multiple hypotheses. The hypothesis for the time-to-first-recovery endpoint was evaluated first, and if the null hypothesis was rejected, the hypothesis for the second co-primary endpoint of hospitalization/death was evaluated. In the context of multiple interim analyses, the master protocol specifies that each null hypothesis is rejected if the Bayesian posterior probability of superiority exceeded 0.99 for the time to recovery endpoint and 0.975 (via gate-keeping) for the hospitalization/death endpoint. Based on trials of antibiotics for lower respiratory tract infection,^27^ a minimum of 1.5 days difference in median time to first report of recovery, and 2% difference in hospitalization/mortality were pre-specified as clinically meaningful.

At the beginning of the trial, due to difficulties with community SARS-CoV-2 PCR testing early on in the pandemic in the United Kingdom, participants with suspected COVID-19 were included in the primary analysis population, irrespective of confirmatory testing. When testing became more readily available, the Trial Steering Committee recommended restricting the primary analysis population to those with confirmed COVID-19, and this change was included in protocol version 7.1 on February 22, 2021 and approved on March 15, 2021, before any interim budesonide results were disclosed to the Trial Management Group, and before those interim results had been reviewed the Trial Steering Committee. Therefore, the pre-specified primary analysis population is defined as all eligible SARS-CoV-2 positive participants randomized to budesonide, usual care, and other interventions, from the start of the platform trial until the data-extraction for this updated interim analysis, on March 25^th^, 2021. Because this population includes participants randomized to usual care before the budesonide group opened, the primary time to recovery analysis model includes parameters to adjust for temporal drift in the study population, which may occur due to changes in circulating SARS-CoV-2, usual care including vaccination, or the pandemic situation, as well as changes in the inclusion/exclusion criteria over time. The inclusion of parameters for temporal drift in the hospitalization/death models are currently being implemented, as pre-specified in the Adaptive Design Report version 3.5 which was approved on March 11, 2021, prior to review of the interim analysis by the Data Monitoring and Safety and Trial Steering Committees which led to unblinding of the budesonide arm. As this change has not yet been completed, we do not present secondary analyses of the hospitalization/death results in the overall study population in this pre-print, as this population contains a large proportion of participants randomized before the budesonide arm opened, and is therefore more susceptible to temporal drift. This pre-specified secondary analysis, accounting for temporal drift, will be presented in the final report.. We also conducted secondary analyses for time to recovery among all study participants irrespective of SARS-CoV-2 status, and pre-specified sensitivity analyses using the concurrent randomized population; defined as all participants who were eligible for budesonide and randomized to budesonide or usual care during the time period when the budesonide arm was active, important because participants already using steroid inhalers, and therefore may have had asthma or COPD, were excluded from randomization to the budesonide arm.

Analysis of the secondary outcomes, and pre-specified sub-group analyses, were conducted on the concurrent randomization and eligible SARS-CoV-2 positive population. Secondary time-to-event outcomes were analyzed using Cox proportional hazard models, and binary outcomes were analyzed using logistic regression, adjusting for comorbidity status, age, and duration of illness.

## Results

### Population

The first participant was randomized on April 2, 2020. Enrolment into the budesonide group started on November 27, 2020. On March 18, 2021, the Trial Steering Committee, after review of planned interim analyses data from March 4, 2021 provided by the Data Monitoring and Safety Committee, advised the Trial Management Group that the pre-specified superiority criterion was met on time to recovery, in both the SARS-CoV-2 positive population and the overall study population. On March 31, 2021, the Trial Steering Committee advised the Trial Management Group to stop randomization to budesonide because accumulating further data to reach pre-specified futility or superiority criteria on hospitalization/death was unlikely due to the successful vaccine rollout and lower than originally anticipated event rate. Results on the hospitalization outcome, restricted to those with complete 28 day follow-up data, were provided to the Trial Management Group on April 5, 2021. Here, we report updated interim analysis results using data extracted on March 25^th^, 2021. Final analyses of complete follow-up data for all participants randomized to inhaled budesonide with 28 day follow up will be published as soon as the data are available and analyzed.

By March 25, 2021, a total of 4663 people were enrolled; 1032 were allocated to budesonide, 1943 to usual care alone, and 1688 to other treatment groups (Figure 1). 3968 (85.1%) had a SARS-CoV-2 test result available, of which 2617 (66.0%) were positive. The Bayesian primary analysis model includes data from 2422/2617 (92.5%) eligible SARS-CoV-2 positive participants who provided some follow up data and were randomized to inhaled budesonide (n = 751), usual care alone (n = 1028), and other treatment groups (n = 643). To protect the integrity of the platform trial and other interventions, we only provide descriptive summaries of participants randomized to budesonide and usual care. The average age (range) of participants was 62.8 (50 – 100) years, of which 2474 (83.2%) had co-morbidities. The median (interquartile range) was 6 (4 to 9) days from symptom onset. Baseline characteristics were comparable between the two groups (Tables 1 and S1).

**Table 1.**
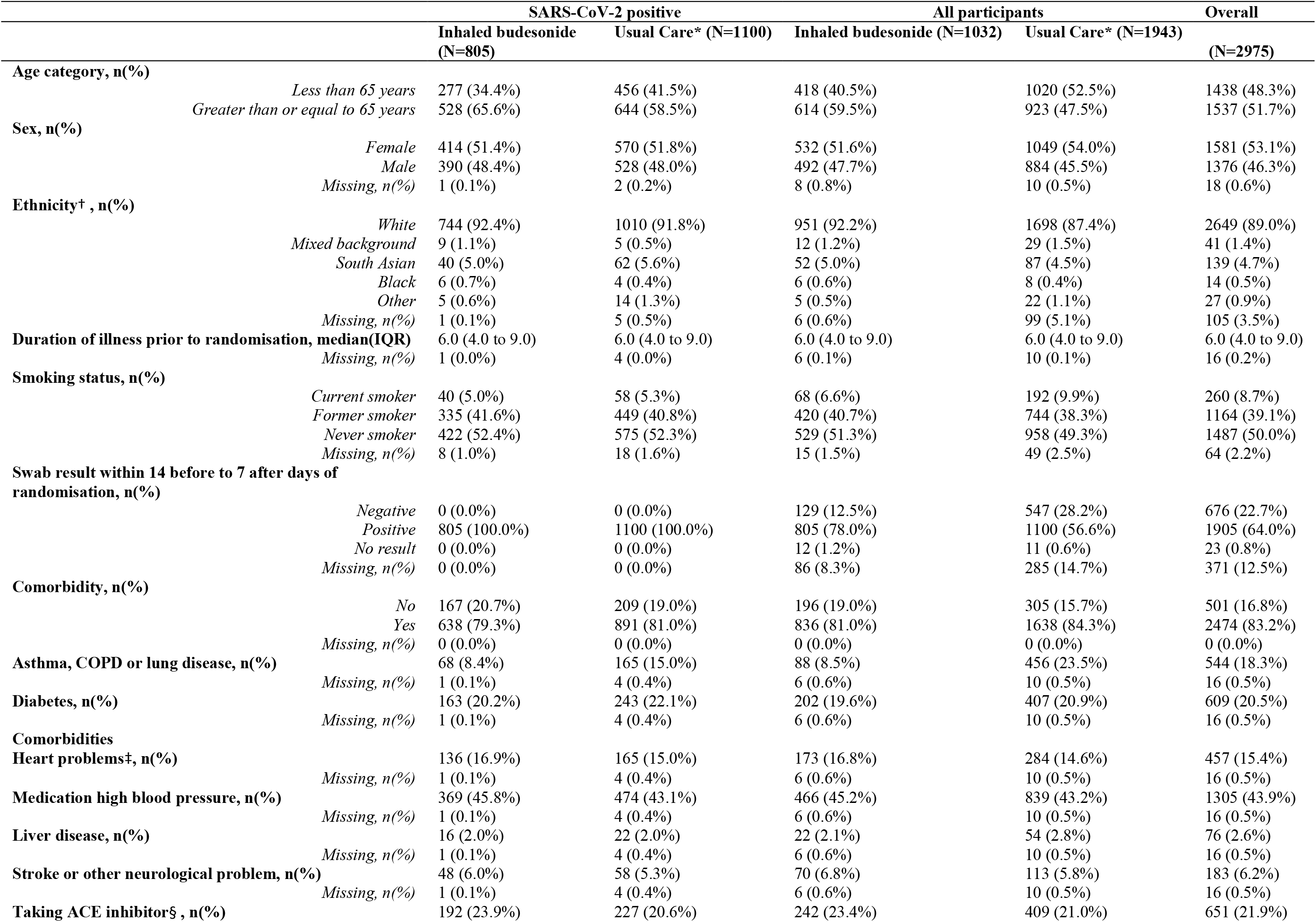
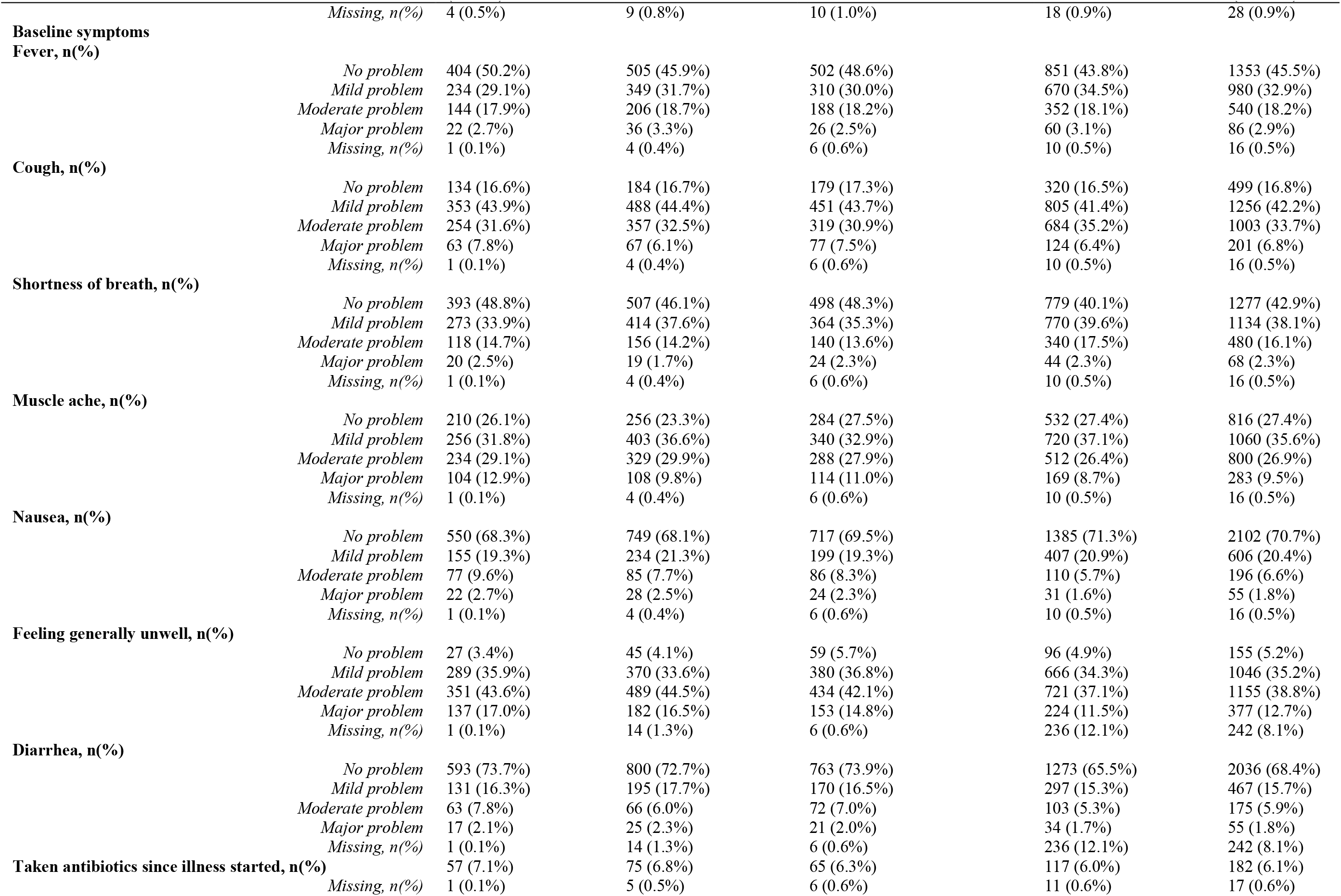
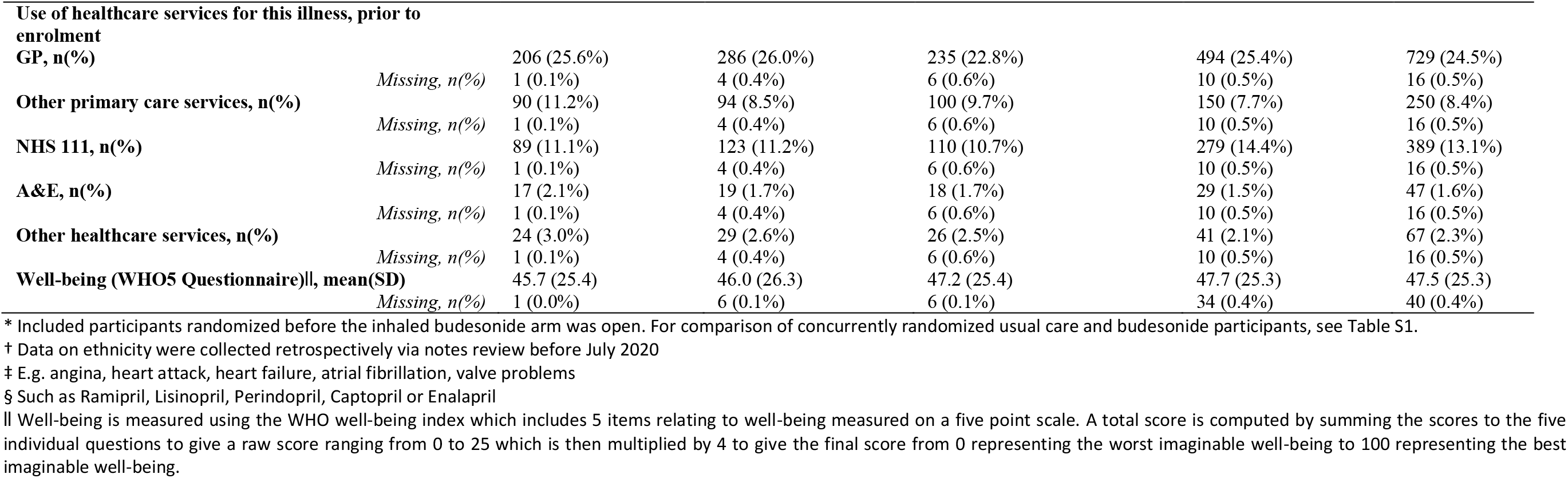
Baseline characteristics participants by treatment group

**Figure 1.**
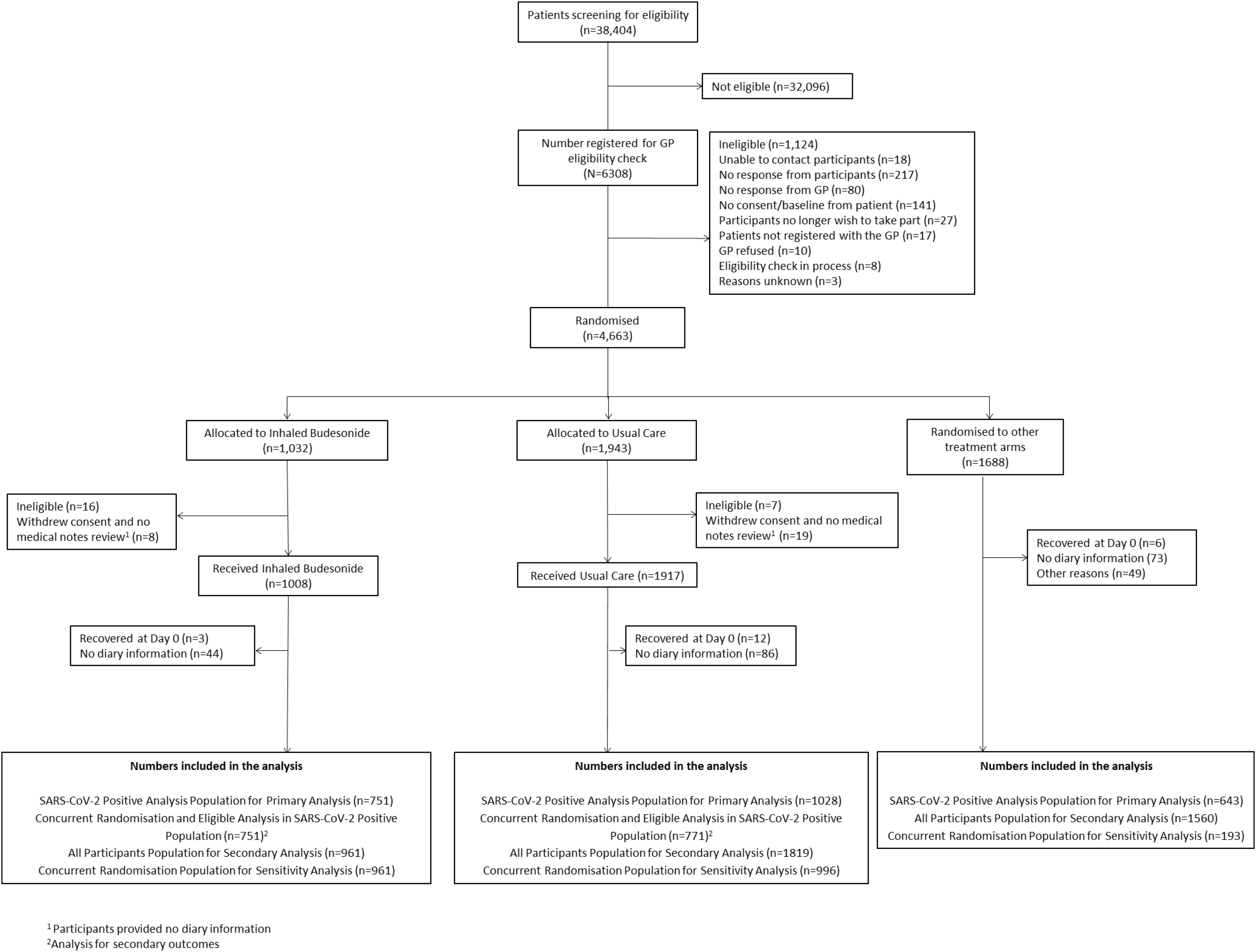
Participant flow diagram

Of 929 participants who provided information about their medication use, 79.9% of participants randomized to budesonide reported taking budesonide for at least 7 days. At the time of data-extraction for this updated analysis on March 25, 2021, 892/961 (92.8%) of all those randomized to budesonide had the opportunity to contribute data for the complete 28 days follow up. A further 13 participants were enrolled to the budesonide arm between the March 25, 2021, and March 31, 2021 when the budesonide arm closed.

### Primary Outcomes

Based on the Bayesian primary analysis model which accounts for temporal changes in time to recovery, there was evidence of a benefit in time-to-first-recovery in the budesonide arm versus usual care (hazard ratio, 1.208; 95% Bayesian Credible Interval [BCI] [1.076 – 1.356], estimated median benefit of 3.011 (95% BCI [1.134 – 5.410] days). The probability that median time to recovery was shorter in budesonide versus usual care (i.e., probability of superiority) was 0.999, which met the pre-specified 0.99 superiority threshold (Table 2). The treatment effect was consistent in the overall study population (hazard ratio, 1.160; 95% BCI [1.049 – 1.283], estimated median benefit of 2.401 (95% BCI [0.755 – 4.588] days, probability of superiority 0.998) and the concurrent randomization analysis population (hazard ratio, 1.180; 95% BCI [1.064 – 1.309], estimated median benefit of 2.586 (95% BCI [0.956 – 4.714] days, probability of superiority 0.999).

**Table 2:** Primary and Secondary Outcomes

|  | Inhaled Budesonide | Usual Care | Estimated treatment effect (95% BCI) | Pr(Superiority) |
| --- | --- | --- | --- | --- |
| **Primary outcomes (SARS-CoV-2 positive population)** |  |  |  |  |
| First reported recovery, n (%) | 534/751 (71.1%) | 666/1028 (64.7%) |  |  |
| Time to first reported recovery, median (IQR) | 11 (5, 27) | 14 (6, -) | 3.011 (1.124 – 5.410)* | 0.999* |
| Hospitalization/death at 28 days, n (%) | 59/692 (8.5%) | 100/968 (10.3%) | 2.1% (-0.7% – 4.8%)† | 0.928† |
| **Primary outcomes (Concurrent randomisation population)** |  |  |  |  |
| First reported recovery, n (%) | 703/961 (73.2%) | 663/996 (66.6%) |  |  |
| Time to first reported recovery, median (IQR) | 10 (4, 25) | 13 (4, -) | 2.59 (0.956 – 4.714)* | 0.999* |
| Hospitalization/death at 28 days, n (%) | 68/892 (7.6%) | 91/928 (9.8%) | 2.2% (-0.4% – 4.8%)† | 0.954† |
| **Secondary outcomes‡** | Inhaled Budesonide | Usual Care | Estimated treatment effect (95% CI) | P-value |
| Early sustained recovery | 221/687 (32.2%) | 156/709 (22.0%) | 1.46 (1.23 to 1.74)§ | <0.0001 |
| Rating of how well participant feels (1 worst, 10 best), mean (SD) [n] |  |  |  |  |
| Day 7 | 7.0 (1.8) [714] | 6.6 (1.9) [730] | 0.35 (0.16 to 0.54) ¶ | <0.0001 |
| Day 14 | 7.9 (1.7) [701] | 7.5 (1.7) [723] | 0.38 (0.17 to 0.59) ¶ | <0.0001 |
| Day 21 | 8.4 (1.5) [572] | 7.9 (1.6) [568] | 0.43 (0.19 to 0.67) ¶ | <0.0001 |
| Day 28 | 8.4 (1.5) [649] | 8.2 (1.5) [662] | 0.21 (-0.06 to 0.48) ¶ | 0.120 |
| Well-being (WHO5 Questionnaire), mean (SD)[n] |  |  |  |  |
| Day 14 | 42.6 (24.9) [673] | 39.1 (24.6) [689] | 3.37 (0.97 to 5.76) ¶ | 0.006 |
| Day 28 | 54.9 (25.2) [612] | 51.2 (24.9) [620] | 3.34 (0.87 to 5.81) ¶ | 0.008 |
| Self-reported contact with ≥1 healthcare service | 400/746 (53.6%) | 440/763 (57.7%) | 0.93 (0.85 to 1.01)# | 0.098 |
| GP reported contact with ≥1 healthcare service | 167/344 (48.5%) | 186/341 (54.5%) | 0.89 (0.77 to 1.03)# | 0.124 |
| Prescription of antibiotics | 31/330 (9.4%) | 28/320 (8.8%) | 1.07 (0.66 to 1.75) # | 0.787 |
| Hospital assessment without admission | 20/750 (2.7%) | 18/771 (2.3%) | 1.14 (0.61 to 2.14) # | 0.744 |
| Oxygen Administration | 43/742 (5.8%) | 64/764 (8.4%) | 0.69 (0.48 to 1.00) # | 0.057 |
| Mechanical ventilation | 11/743 (1.5%) | 11/760 (1.4%) | 1.02 (0.45 to 2.34)** | >0.99 |
| ICU admission | 9/735 (1.2%) | 17/756 (2.2%) | 0.54 (0.24 to 1.21)** | 0.166 |

\* Estimated benefit in median time to recovery derived from a Bayesian piecewise exponential model adjusted for age and comorbidity at baseline, with 95% Bayesian credible interval. A positive value corresponds to a reduction in time to recovery in days. Pr(Superiority) is the probability of superiority and treatment superiority is declared if Pr(superiority) ≥ 0.99 versus usual care.
† Estimated percentage benefit in hospitalization/death rate derived from a Bayesian logistic regression model adjusted for age and comorbidity at baseline, with 95% Bayesian credible interval. A positive value favors inhaled budesonide. Pr(Superiority) is the probability of superiority and treatment superiority is declared if Pr(superiority) ≥ 0.975 versus usual care
‡ All secondary outcome analyses were conducted on the concurrent randomization and eligible analysis population in participants with SARS-CoV-2 positive analysis population, but restricted to those in the inhaled budesonide and usual care group only.
§ Estimated relative risk derived from a logistic regression model adjusted for age, comorbidity at baseline, duration of illness, at baseline, with 95% confidence interval.
|| Estimated hazard ratio derived from a Cox proportional hazard model adjusted for age, comorbidity at baseline, duration of illness, at baseline, with 95% confidence interval.
¶ Mixed effect model adjusting age, comorbidity, duration of illness at baseline, and time. Participant was fitted as a random effect. WHO well-being score was also adjusted for the score at baseline

In the preliminary primary Bayesian analysis of hospitalizations (which do not currently account for temporal changes in hospitalizations, but will do so in the final analysis, as pre-specified in the Adaptive Design Report version 3.5), among those who had the opportunity to contribute data for 28 days follow up, the point estimate of the proportion of COVID-19 related hospitalization/deaths within 28 days follow up was slightly lower in the budesonide group compared to usual care (59/692 [8.5%] vs 100/968 [10.3%]; estimated percentage difference, 2.1%; 95% BCI −0.7 – 4.8%) (Table 2). The probability that COVID-19 hospitalizations/deaths were lower in the budesonide versus usual care (probability of superiority) was 0.928, which did not meet the predefined superiority threshold of 0.975. Results were similar in the concurrent randomized analysis population (68/892 [7.6%] vs 91/928 [9.8%]; estimated percentage difference, 2.2%; 95% BCI −0.4 – 4.8%, probability of superiority 0.954) (Table 2). Results from the overall analysis population are not reported here, as this population contains a large proportion of usual care participants randomized before the budesonide arm opened, and is therefore more sensitive to changes hospitalization rates over time. The final, complete analysis will include parameters for time in the Bayesian primary outcome models, as pre-specified in the Adaptive Design Report version 3.5.

### Secondary outcomes

Analysis of secondary outcomes, using the concurrent randomization and eligible SARS-CoV-2 positive population, showed evidence of benefit with budesonide in the daily score of how well participants felt over 28 days (Table 2 and Figure S1), the WHO-5 Wellbeing Index, early sustained recovery, time to sustained recovery, (Table 2). There was no clear evidence of differences in both participants reported or GP reported healthcare services use between groups (Table 2).

In the prespecified subgroup analyses, there was no clear evidence that symptom duration prior to randomization, baseline illness severity score, age or comorbidity modified the effect of budesonide on time to first reported recovery (Figure 3). Regarding serious adverse events, two participants reported hospitalizations unrelated to COVID-19, both in the budesonide group.

**Figure 2.**
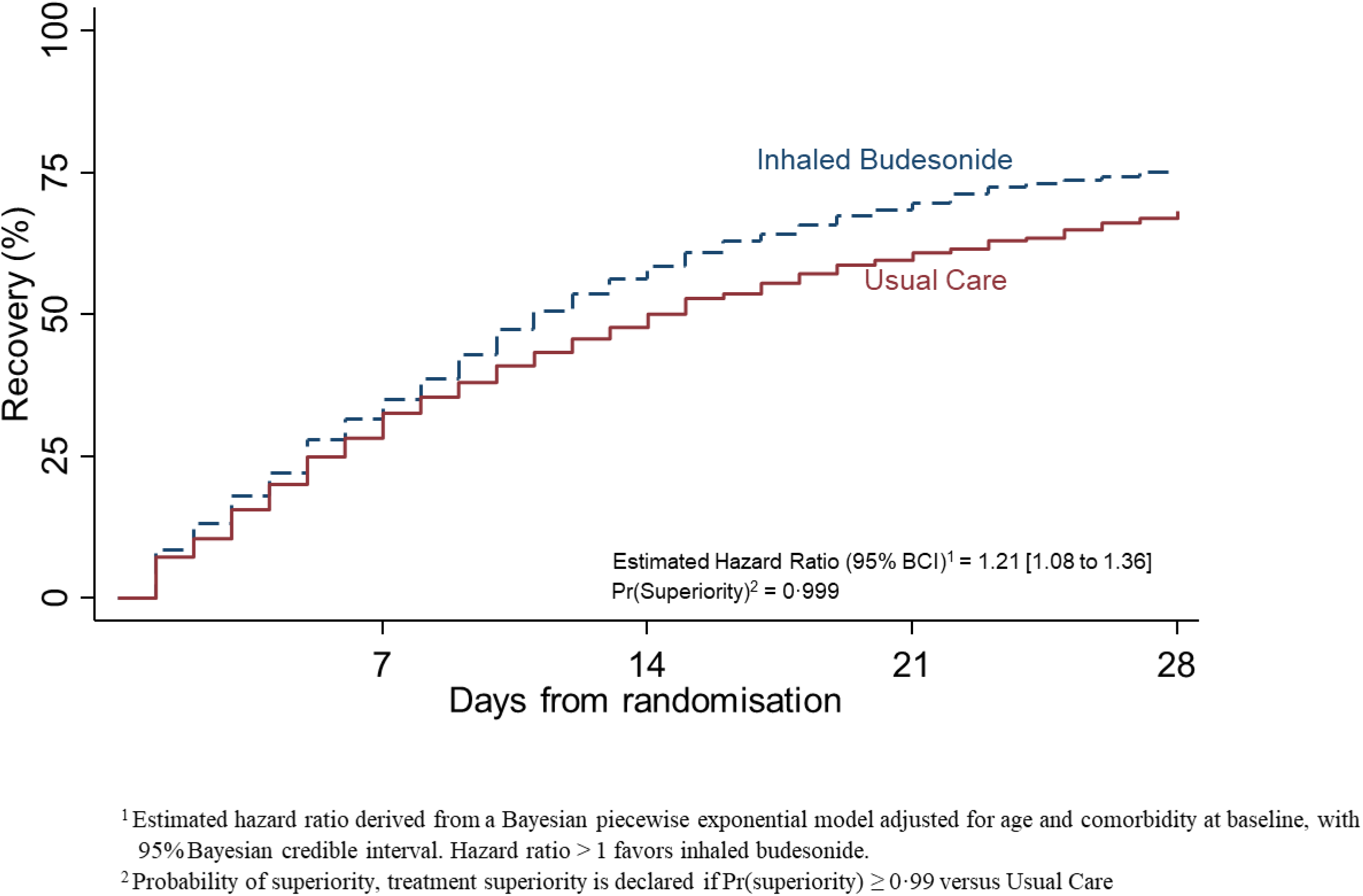
Summary and results of the time to first self-reported recovery SARS-CoV-2 positive analysis population and data extracted on March 25, 2021

**Figure 3.**
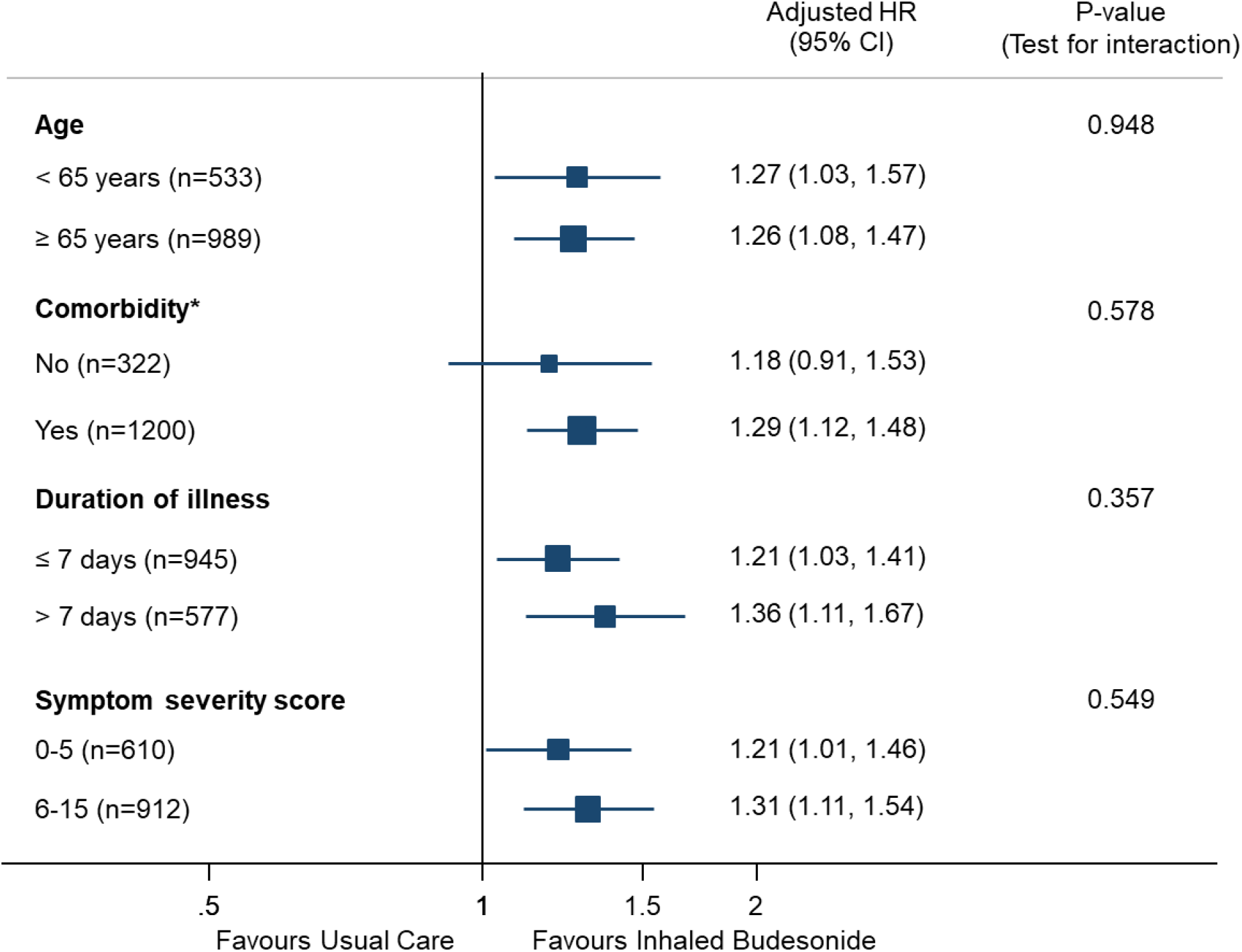
Forest plot of subgroup analysis on time-to-recovery outcome (concurrent randomisation and eligible analysis population in participant with SARS-CoV-2 positive) at time of data extracted on March 25, 2021.

## DISCUSSION

This interim analysis from a platform, randomized trial involving participants in the community with COVID-19 at increased risk of an adverse outcome, found that inhaled budesonide shortened time to first self-reported recovery. Our findings were consistent across various measures that capture symptom severity, sustained recovery, wellbeing. There was also no evidence of differential effects among pre-specified sub-groups by age, symptom duration, symptom severity and co-morbidity. Due to decreases in hospital admissions associated with the United Kingdom lockdown and vaccination program,^28,29^ and yet to be completed 28 day follow up data for some participants, the current interim analysis is not powered for the COVID-19 related hospital admission and/or death outcome. The final analysis of the effects of inhaled budesonide will be made available once all participants randomized to inhaled budesonide have completed 28 day follow up.

We identified eight ongoing randomized controlled trials of inhaled corticosteroids as treatment for COVID-19,^30^ and one completed study (the STOIC study). In the STOIC phase 2, open-label trial among adults aged 18 and over with mild, suspected COVID-19 in the community, 146 participants were randomly assigned to inhaled budesonide 800µg twice a day until symptoms resolved, or usual care.^14^ The primary outcome of COVID-19 related urgent care or emergency department assessment, or hospitalization, was achieved in 1/70 budesonide participants versus 10/69 usual care participants (difference in proportions 0.131, 95% CI 0.043 to 0.218, p=0.004). We did not combine different types of healthcare utilization into one outcome in the PRINCIPLE trial. Secondary outcomes in STOIC also favored budesonide over usual care with respect to time to self-reported recovery, symptom persistence at day 14, and resolution of fever.

PRINCIPLE is the largest randomized trial to date to evaluate inhaled budesonide for community treatment of COVID-19 and adds to the evidence base supporting this therapeutic agent following the earlier Phase 2 findings.^31^ Furthermore, recent transcriptome analysis indicates that budesonide, alongside others which include dexamethasone and ciclesonide is a novel therapeutic immunomodulatory target.^32^ Several randomized trials have demonstrated the benefits of systemic corticosteroids for treatment of people hospitalized with COVID-19.^8,9^ Our findings are immediately relevant for clinical practice as they suggest that early treatment in the community with inhaled corticosteroids is effective at speeding recovery, which has important benefits for patients and wider society. While global access to vaccines continues to be scaled up, inhaled budesonide is readily available in many primary care settings and is included in the World Health Organization List of Essential Medicines.^33^.

Strengths of our analysis include the pragmatic trial design which allows for efficient evaluation of multiple interventions as they would be used in the community, the evaluation of budesonide as a standalone, early treatment and the focus on patients at higher risk of complications. We included patients with suspected COVID-19 but without PCR confirmed SARS-CoV-2 infection in our secondary analyses as this reflects community testing conditions early in the UK pandemic, and limited SARS-CoV-2 testing may necessitate early empirical treatment in many other community and low resource hospital settings. Given the variation in PCR testing sensitivity, particularly if self-administered, some participants will have had false negative tests.^34-37^ We conducted an open label study to determine whether the addition of budesonide to usual care benefitted patients, rather than to assess benefit of budesonide compared to a placebo. The pragmatic study design therefore does not allow us to determine mechanism of effect or the degree to which a placebo effect influenced outcomes. However, we found no evidence of a placebo effect in evaluations of other treatments in this trial platform.^16,17^

In conclusion, in this updated interim analysis, inhaled budesonide improved time to recovery by a median 3 days when used to treat COVID-19 in people at higher risk of adverse outcomes in the community.

## Supporting information

CONSORT Checklist

Study protocol

## Data Availability

Data can be shared with qualifying researchers who submit a proposal with a valuable research question as assessed by a committee formed from the TMG including senior statistical and clinical representation. A contract should be signed.

## Writing group contributions

CCB and FRDH had full access to all of the data in the study and take responsibility for the integrity of the data and the accuracy of the data analysis. CCB and FDRH decided to publish the paper. MB, PJB, REKR, DVN, and SR, contributed to the development of the intervention specific appendix for inhaled budesonide. BS, NB, L-MY, CCB, FDRH, GH, OVH, OG, JD, contributed to trial design. SdeL, PHE, NT and MPG helped plan the trial and ongoing recruitment. EO, NT, SdeL, were responsible for acquisition of data. CCB, JD, BS, LMY, FDRH, GH, OVH and OG drafted the manuscript. BS, NB, L-MY, MAD, MF, CS, and VH contributed to statistical analysis. All members of the PRINCIPLE writing group critically revised the manuscript. The members of the PRINCIPLE Collaborative Group and their roles in the conduct of the trial are listed at the end of the manuscript.

## Acknowledgments

We thank the patients who participated in this study. We also thank the many health, and social care professionals and who contributed. The PRINCIPLE trial platform is led from the Primary Care and Vaccines Collaborative Clinical Trials Unit at the University of Oxford’s Nuffield Department of Primary Care Health Sciences. PRINCIPLE is supported by a large network of care homes, pharmacies, NHS 111 Hubs, hospitals, and 1,401 GP practices across England, Wales, Scotland, and Northern Ireland. The trial is integrated with the Oxford-Royal College of General Practitioners (RCGP) Research and Surveillance Centre (RSC) ORCHID digital platform. PRINCIPLE has been supported by the NIHR and its Clinical Research Network, NHS DigiTrials, Public Health England, Health and Care Research Wales, NHS Research Scotland, the Health and Social Care Board in Northern Ireland, and the Therapeutics Task Force.

CCB acknowledges part support as Senior Investigator of the National Institute of Health Research, the NIHR Community Healthcare Medtech and In-Vitro Diagnostics Co-operative (MIC), and the NIHR Health Protection Research Unit on Health Care Associated Infections and Antimicrobial Resistance. FDRH acknowledges his part-funding from the National Institute for Health Research (NIHR) School for Primary Care Research, the NIHR Applied Research Collaboration (ARC) Oxford, the NIHR Oxford Biomedical Research Centre (BRC, UHT), and the NIHR Community Healthcare Medtech and In-Vitro Diagnostics Co-operative (MIC). JD and OG are funded by the Wellcome Trust PhD Programme for Primary Care Clinicians (216421/Z/19/Z and 203921/Z/16/Z respectively). GH is funded by an NIHR Advanced Fellowship and by the NIHR Community Healthcare Medtech and In-Vitro Diagnostics Co-operative (MIC).For the purpose of Open Access, the author has applied a CC BY public copyright license to any Author Accepted Manuscript version arising from this submission.

## PRINCIPLE COLLABORATIVE GROUP

### PRINCIPLE Trial Management Group

Julie Allen^1^, Monique Andersson^2^, Nick Berry^3*^, Emily Bongard^1^, Aleksandra Borek^1^, Christopher C Butler^1^ (Chair), Simon de Lusignan^1^, Jienchi Dorward^1,4^, Philip H Evans^5,6^, Filipa Ferreira^1^, Oghenekome Gbinigie^1^, Jenna Grabey^1^, Gail Hayward^1^, FD Richard Hobbs^1^, Susan Hopkins^7^, David Judge^1^, Mona Koshkouei^1^, Martin J Llewelyn^8^, Emma Ogburn^1^, Mahendra G. Patel^1^, Dan Richards-Doran^1^, Heather Rutter^1^, Benjamin R. Saville^2,9^, Hannah Swayze^1^, Nicholas PB Thomas^5,10^, Manasa Tripathy^1^, Sarah Tonkin-Crine^1,11^, Jared Robinson^1^, Oliver Van Hecke^1^, Ly-Mee Yu^1^

* TMG membership ended when NB became unblinded and joined the SAC

Trial Management Group affiliations

1. Nuffield Department of Primary Care Health Sciences, University of Oxford, Oxford, UK
2. Department of Microbiology, Oxford University Hospitals NHS Trust, Oxford, UK
3. Berry Consultants, Texas, USA,
4. Centre for the AIDS Programme of Research in South Africa (CAPRISA), University of KwaZulu– Natal, Durban, South Africa.
5. National Institute for Health Research, Clinical Research Network
6. College of Medicine and Health, University of Exeter
7. Public Health England, London, UK
8. Brighton and Sussex Medical School, University of Sussex, Brighton, UK
9. Department of Biostatistics, Vanderbilt University School of Medicine, Tennessee, USA
10. Royal College of General Practitioners, London, UK
11. National Institute for Health Protection Research Unit in Healthcare Associated Infections and Antimicrobial Resistance, Oxford, UK

### Statistical Analysis Committee

Nick Berry, Michelle A. Detry (Chair), Christina Saunders, Mark Fitzgerald

### Trial Steering Committee

Carol Green, Phil Hannaford, Paul Little (Chair), Tim Mustill, Matthew Sydes

### Data Monitoring and Safety Committee

Deborah Ashby (Chair), Nick Francis, Simon Gates, Gordon Taylor, Patrick White

### Principle Trial Coordinating Office

Co-Study Leads: CC Butler, FDR Hobbs

Trial management: E Ogburn (coordinator), H Swayze, E Bongard, J Allen, S Tonner, R Edeson, J Brooks, R Edwards, N Maeder, S Barrett, S Brann, A Maloney, K Dempster, J de Henau, J Robinson, N Begum

Clinical Team: H Rutter (Coordinator), K Madronal, B Mundy, B Ianson, I Noel, B Thompson, O Gbinigie, J Dorward, G Hayward, O van Hecke, N Jones, H van der Westhuizen, K Kotze

Data and Programming Team: D Judge, J Grabey, L Castello, D Watt, R Zhao

Statistics: B Saville, L-M Yu, N Berry, M Detry, V Harris, C Saunders, M Fitzgerald, J Mollison, M Shanyinde

Oxford-Royal College of General Practitioner’s Research and Surveillance Centre: S de Lusignan (coordinator) M Tripathy, F Ferreira

National Institute for Health Research Clinical Research Network Coordinating Centre: PH Evans, NPB Thomas, H Collins, K Priddis, L Owen, K Hannaby, B Drew

## Supplementary Figures and Tables (using data from March 25^th^, 2021)

**Figure S1.**
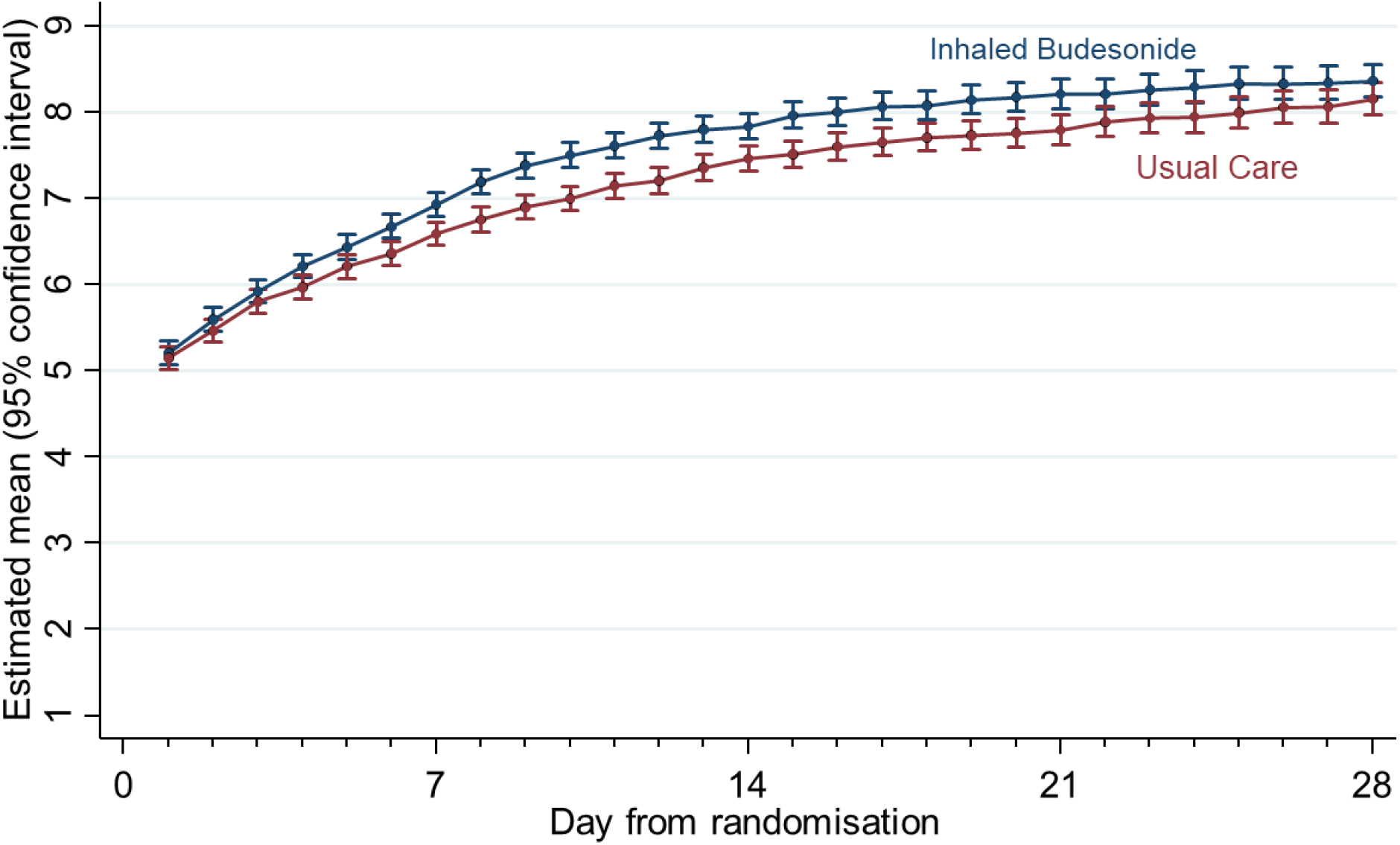
Estimated mean and 95% confidence interval of daily rating of feeling well over the 28 days follow-up by treatment arm using data extracted on 25^th^ March 2021 (Concurrent Randomisation and Eligible Analysis population in participants with SARS-CoV-2 positive)

**Table S1.** Baseline characteristics for concurrent randomization analysis population

|  | SARS-CoV-2 positive |  | All participants |  | Overall<br>GROUP Overall<br>(N=2112) |
| --- | --- | --- | --- | --- | --- |
|  | GROUP Budesonide<br>(N=805) | GROUP Usual Care<br>(N=860) | GROUP Budesonide<br>(N=1032) | GROUP Usual Care<br>(N=1080) |  |
| **Age category, n(%)** |  |  |  |  |  |
| *Less than 65 years* | 277 (34.4%) | 303 (35.2%) | 418 (40.5%) | 442 (40.9%) | 860 (40.7%) |
| *Greater than or equal to 65 years* | 528 (65.6%) | 557 (64.8%) | 614 (59.5%) | 638 (59.1%) | 1252 (59.3%) |
| **Sex, n(%)** |  |  |  |  |  |
| *Female* | 414 (51.4%) | 439 (51.0%) | 532 (51.6%) | 561 (51.9%) | 1093 (51.8%) |
| *Male* | 390 (48.4%) | 419 (48.7%) | 492 (47.7%) | 510 (47.2%) | 1002 (47.4%) |
| *Missing, n(%)* | 1 (0.1%) | 2 (0.2%) | 8 (0.8%) | 9 (0.8%) | 17 (0.8%) |
| **Ethnicity\*, n(%)** |  |  |  |  |  |
| *White* | 744 (92.4%) | 792 (92.1%) | 951 (92.2%) | 992 (91.9%) | 1943 (92.0%) |
| *Mixed background* | 9 (1.1%) | 4 (0.5%) | 12 (1.2%) | 7 (0.6%) | 19 (0.9%) |
| *South Asian* | 40 (5.0%) | 46 (5.3%) | 52 (5.0%) | 55 (5.1%) | 107 (5.1%) |
| *Black* | 6 (0.7%) | 3 (0.3%) | 6 (0.6%) | 3 (0.3%) | 9 (0.4%) |
| *Other* | 5 (0.6%) | 11 (1.3%) | 5 (0.5%) | 13 (1.2%) | 18 (0.9%) |
| *Missing, n(%)* | 1 (0.1%) | 4 (0.5%) | 6 (0.6%) | 10 (0.9%) | 16 (0.8%) |
| **Duration of illness prior to randomization,**<br>**median(IQR)** | 6.0 (4.0 to 9.0) | 6.0 (4.0 to 9.0) | 6.0 (4.0 to 9.0) | 6.0 (4.0 to 9.0) | 6.0 (4.0 to 9.0) |
| *Missing, n(%)* | 1 (0.0%) | 4 (0.1%) | 6 (0.1%) | 10 (0.2%) | 16 (0.3%) |
| **Smoking status, n(%)** |  |  |  |  |  |
| *Current smoker* | 40 (5.0%) | 43 (5.0%) | 68 (6.6%) | 76 (7.0%) | 144 (6.8%) |
| *Former smoker* | 335 (41.6%) | 352 (40.9%) | 420 (40.7%) | 426 (39.4%) | 846 (40.1%) |
| *Never smoker* | 422 (52.4%) | 451 (52.4%) | 529 (51.3%) | 555 (51.4%) | 1084 (51.3%) |
| *Missing, n(%)* | 8 (1.0%) | 14 (1.6%) | 15 (1.5%) | 23 (2.1%) | 38 (1.8%) |
| **Swab result, n(%)** |  |  |  |  |  |
| *Negative* | 0 (0.0%) | 0 (0.0%) | 129 (12.5%) | 138 (12.8%) | 267 (12.6%) |
| *Positive* | 805 (100.0%) | 860 (100.0%) | 805 (78.0%) | 860 (79.6%) | 1665 (78.8%) |
| *No result* | 0 (0.0%) | 0 (0.0%) | 12 (1.2%) | 5 (0.5%) | 17 (0.8%) |
| *Missing, n(%)* | 0 (0.0%) | 0 (0.0%) | 86 (8.3%) | 77 (7.1%) | 163 (7.7%) |
| **Comorbidity, n(%)** |  |  |  |  |  |
| *No* | 167 (20.7%) | 180 (20.9%) | 196 (19.0%) | 203 (18.8%) | 399 (18.9%) |
| *Yes* | 638 (79.3%) | 680 (79.1%) | 836 (81.0%) | 877 (81.2%) | 1713 (81.1%) |
| **Asthma, COPD or lung disease, n(%)** | 68 (8.4%) | 87 (10.1%) | 88 (8.5%) | 127 (11.8%) | 215 (10.2%) |
| *Missing, n(%)* | 1 (0.1%) | 4 (0.5%) | 6 (0.6%) | 10 (0.9%) | 16 (0.8%) |
| **Diabetes, n(%)** | 163 (20.2%) | 192 (22.3%) | 202 (19.6%) | 246 (22.8%) | 448 (21.2%) |
| *Missing, n(%)* | 1 (0.1%) | 4 (0.5%) | 6 (0.6%) | 10 (0.9%) | 16 (0.8%) |
| **Comorbidities** |  |  |  |  |  |
| **Heart problems†, n(%)** | 136 (16.9%) | 128 (14.9%) | 173 (16.8%) | 158 (14.6%) | 331 (15.7%) |
| *Missing, n(%)* | 1 (0.1%) | 4 (0.5%) | 6 (0.6%) | 10 (0.9%) | 16 (0.8%) |
|  | GROUP Budesonide<br>(N=805) | GROUP Usual Care<br>(N=860) | GROUP Budesonide<br>(N=1032) | GROUP Usual Care<br>(N=1080) |  |
| **Medication for high blood pressure, n(%)** | 369 (45.8%) | 376 (43.7%) | 466 (45.2%) | 470 (43.5%) | 936 (44.3%) |
| Missing, n(%) | 1 (0.1%) | 4 (0.5%) | 6 (0.6%) | 10 (0.9%) | 16 (0.8%) |
| **Liver disease, n(%)** | 16 (2.0%) | 20 (2.3%) | 22 (2.1%) | 31 (2.9%) | 53 (2.5%) |
| Missing, n(%) | 1 (0.1%) | 4 (0.5%) | 6 (0.6%) | 10 (0.9%) | 16 (0.8%) |
| **Stroke or other neurological problem, n(%)** | 48 (6.0%) | 42 (4.9%) | 70 (6.8%) | 61 (5.6%) | 131 (6.2%) |
| Missing, n(%) | 1 (0.1%) | 4 (0.5%) | 6 (0.6%) | 10 (0.9%) | 16 (0.8%) |
| **Taking ACE inhibitor‡, n(%)** | 192 (23.9%) | 177 (20.6%) | 242 (23.4%) | 229 (21.2%) | 471 (22.3%) |
| Missing, n(%) | 4 (0.5%) | 6 (0.7%) | 10 (1.0%) | 14 (1.3%) | 24 (1.1%) |
| **Baseline symptoms** |  |  |  |  |  |
| **Fever, n(%)** |  |  |  |  |  |
| No problem | 404 (50.2%) | 394 (45.8%) | 502 (48.6%) | 479 (44.4%) | 981 (46.4%) |
| Mild problem | 234 (29.1%) | 280 (32.6%) | 310 (30.0%) | 372 (34.4%) | 682 (32.3%) |
| Moderate problem | 144 (17.9%) | 152 (17.7%) | 188 (18.2%) | 181 (16.8%) | 369 (17.5%) |
| Major problem | 22 (2.7%) | 30 (3.5%) | 26 (2.5%) | 38 (3.5%) | 64 (3.0%) |
| Missing, n(%) | 1 (0.1%) | 4 (0.5%) | 6 (0.6%) | 10 (0.9%) | 16 (0.8%) |
| **Cough, n(%)** |  |  |  |  |  |
| No problem | 134 (16.6%) | 132 (15.3%) | 179 (17.3%) | 165 (15.3%) | 344 (16.3%) |
| Mild problem | 353 (43.9%) | 371 (43.1%) | 451 (43.7%) | 468 (43.3%) | 919 (43.5%) |
| Moderate problem | 254 (31.6%) | 293 (34.1%) | 319 (30.9%) | 368 (34.1%) | 687 (32.5%) |
| Major problem | 63 (7.8%) | 60 (7.0%) | 77 (7.5%) | 69 (6.4%) | 146 (6.9%) |
| Missing, n(%) | 1 (0.1%) | 4 (0.5%) | 6 (0.6%) | 10 (0.9%) | 16 (0.8%) |
| **Shortness of breath, n(%)** |  |  |  |  |  |
| No problem | 393 (48.8%) | 413 (48.0%) | 498 (48.3%) | 508 (47.0%) | 1006 (47.6%) |
| Mild problem | 273 (33.9%) | 307 (35.7%) | 364 (35.3%) | 388 (35.9%) | 752 (35.6%) |
| Moderate problem | 118 (14.7%) | 118 (13.7%) | 140 (13.6%) | 149 (13.8%) | 289 (13.7%) |
| Major problem | 20 (2.5%) | 18 (2.1%) | 24 (2.3%) | 25 (2.3%) | 49 (2.3%) |
| Missing, n(%) | 1 (0.1%) | 4 (0.5%) | 6 (0.6%) | 10 (0.9%) | 16 (0.8%) |
| **Muscle ache, n(%)** |  |  |  |  |  |
| No problem | 210 (26.1%) | 196 (22.8%) | 284 (27.5%) | 265 (24.5%) | 549 (26.0%) |
| Mild problem | 256 (31.8%) | 317 (36.9%) | 340 (32.9%) | 386 (35.7%) | 726 (34.4%) |
| Moderate problem | 234 (29.1%) | 254 (29.5%) | 288 (27.9%) | 317 (29.4%) | 605 (28.6%) |
| Major problem | 104 (12.9%) | 89 (10.3%) | 114 (11.0%) | 102 (9.4%) | 216 (10.2%) |
| Missing, n(%) | 1 (0.1%) | 4 (0.5%) | 6 (0.6%) | 10 (0.9%) | 16 (0.8%) |
| **Nausea, n(%)** |  |  |  |  |  |
| No problem | 550 (68.3%) | 573 (66.6%) | 717 (69.5%) | 739 (68.4%) | 1456 (68.9%) |
| Mild problem | 155 (19.3%) | 186 (21.6%) | 199 (19.3%) | 226 (20.9%) | 425 (20.1%) |
| Moderate problem | 77 (9.6%) | 73 (8.5%) | 86 (8.3%) | 81 (7.5%) | 167 (7.9%) |
| Major problem | 22 (2.7%) | 24 (2.8%) | 24 (2.3%) | 24 (2.2%) | 48 (2.3%) |
| Missing, n(%) | 1 (0.1%) | 4 (0.5%) | 6 (0.6%) | 10 (0.9%) | 16 (0.8%) |
|  | GROUP Budesonide<br>(N=805) | GROUP Usual Care<br>(N=860) | GROUP Budesonide<br>(N=1032) | GROUP Usual Care<br>(N=1080) |  |
| **Feeling generally unwell, n(%)** |  |  |  |  |  |
| *No problem* | 27 (3.4%) | 30 (3.5%) | 59 (5.7%) | 53 (4.9%) | 112 (5.3%) |
| *Mild problem* | 289 (35.9%) | 281 (32.7%) | 380 (36.8%) | 374 (34.6%) | 754 (35.7%) |
| *Moderate problem* | 351 (43.6%) | 375 (43.6%) | 434 (42.1%) | 456 (42.2%) | 890 (42.1%) |
| *Major problem* | 137 (17.0%) | 170 (19.8%) | 153 (14.8%) | 187 (17.3%) | 340 (16.1%) |
| *Missing, n(%)* | 1 (0.1%) | 4 (0.5%) | 6 (0.6%) | 10 (0.9%) | 16 (0.8%) |
| **Diarrhea, n(%)** |  |  |  |  |  |
| *No problem* | 593 (73.7%) | 633 (73.6%) | 763 (73.9%) | 795 (73.6%) | 1558 (73.8%) |
| *Mild problem* | 131 (16.3%) | 147 (17.1%) | 170 (16.5%) | 186 (17.2%) | 356 (16.9%) |
| *Moderate problem* | 63 (7.8%) | 55 (6.4%) | 72 (7.0%) | 65 (6.0%) | 137 (6.5%) |
| *Major problem* | 17 (2.1%) | 21 (2.4%) | 21 (2.0%) | 24 (2.2%) | 45 (2.1%) |
| *Missing, n(%)* | 1 (0.1%) | 4 (0.5%) | 6 (0.6%) | 10 (0.9%) | 16 (0.8%) |
| **Taken antibiotics since illness started, n(%)** | 57 (7.1%) | 68 (7.9%) | 65 (6.3%) | 80 (7.4%) | 145 (6.9%) |
| *Missing, n(%)* | 1 (0.1%) | 4 (0.5%) | 6 (0.6%) | 10 (0.9%) | 16 (0.8%) |
| **Use of healthcare services** |  |  |  |  |  |
| **GP, n(%)** | 206 (25.6%) | 215 (25.0%) | 235 (22.8%) | 251 (23.2%) | 486 (23.0%) |
| *Missing, n(%)* | 1 (0.1%) | 4 (0.5%) | 6 (0.6%) | 10 (0.9%) | 16 (0.8%) |
| **Other primary care services, n(%)** | 90 (11.2%) | 86 (10.0%) | 100 (9.7%) | 96 (8.9%) | 196 (9.3%) |
| *Missing, n(%)* | 1 (0.1%) | 4 (0.5%) | 6 (0.6%) | 10 (0.9%) | 16 (0.8%) |
| **NHS 111, n(%)** | 89 (11.1%) | 92 (10.7%) | 110 (10.7%) | 117 (10.8%) | 227 (10.7%) |
| *Missing, n(%)* | 1 (0.1%) | 4 (0.5%) | 6 (0.6%) | 10 (0.9%) | 16 (0.8%) |
| **A&E, n(%)** | 17 (2.1%) | 15 (1.7%) | 18 (1.7%) | 15 (1.4%) | 33 (1.6%) |
| *Missing, n(%)* | 1 (0.1%) | 4 (0.5%) | 6 (0.6%) | 10 (0.9%) | 16 (0.8%) |
| **Other healthcare services, n(%)** | 24 (3.0%) | 24 (2.8%) | 26 (2.5%) | 27 (2.5%) | 53 (2.5%) |
| *Missing, n(%)* | 1 (0.1%) | 4 (0.5%) | 6 (0.6%) | 10 (0.9%) | 16 (0.8%) |
| **Well-being (WHO5 Questionnaire) §, mean(SD)** | 45.7 (25.4) | 44.9 (26.5) | 47.2 (25.4) | 46.1 (26.1) | 46.7 (25.7) |
| *Missing, n(%)* | 1 (0.0%) | 4 (0.1%) | 6 (0.1%) | 10 (0.2%) | 16 (0.3%) |

\* Data on ethnicity were collected retrospectively via notes review before July 2020
† E.g. angina, heart attack, heart failure, atrial fibrillation, valve problems
‡ Such as Ramipril, Lisinopril, Perindopril, Captopril or Enalapril
§ Well-being is measured using the WHO well-being index which includes 5 items relating to well-being measured on a five point scale. A total score is computed by summing the scores to the five individual questions to give a raw score ranging from 0 to 25 which is then multiplied by 4 to give the final score from 0 representing the worst imaginable well-being to 100 representing the best imaginable well-being.

