## Supplementary material for "Inhaled budesonide for COVID-19 in people at higher risk of adverse outcomes in the community: interim analyses from the PRINCIPLE trial": Study protocol

**Trial Title:** Platform Randomised trial of INterventions against COVID-19 In older peoPLE

**Internal Reference Number / Short title:** PRINCIPLE
**Ethics Ref:** 20/SC/0158
**IRAS Project ID:** 281958
**EudraCT Number**: 2020-001209-22
**Date and Version No**: 22nd February 2021 version 7.1

| | |
|---|---|
| **Chief Investigator and trial leader:** | Professor Chris Butler, Department of Primary Care Health Sciences University of Oxford |
| **Co-Principal Investigator and Co-trial lead:** | Prof Richard Hobbs, Department of Primary Care Health Sciences University of Oxford |
| **Co-Principal Investigators:** | Prof Simon de Lusignan, RCGP Research Surveillance Centre, University of Oxford<br><br>Prof Gail Hayward, Department of Primary Care Health Sciences University of Oxford |
| **Investigators:** | Dr Ly-Mee Yu, Primary Care Clinical Trials Unit, Department of Primary Care Health Sciences, University of Oxford<br><br>Dr Emma Ogburn, Primary Care Clinical Trials Unit, Department of Primary Care Health Sciences, University of Oxford<br><br>Dr Oliver Van Hecke, Department of Primary Care Health Sciences, University of Oxford<br><br>Ms Julie Allen, Primary Care Clinical Trials Unit, Department of Primary Care Health Sciences, University of Oxford<br><br>Dr Emily Bongard, Primary Care Clinical Trials Unit, Department of Primary Care Health Sciences, University of Oxford<br><br>Dr Hannah Swayze, Primary Care Clinical Trials Unit, Department of Primary Care Health Sciences, University of Oxford<br><br>Ben Saville, PhD, Berry Consultants, Texas, USA, & Department of Biostatistics, Vanderbilt University School of Medicine, Tennessee, USA<br><br>Prof Martin Llewellyn, Professor in Infectious Diseases, Medical Research Building, Room 1.08, BSMS, University of Sussex<br><br>Dr Oghenekome Gbinigie, Department of Primary Care Health Sciences University of Oxford<br><br>Dr Jienchi Dorward, Department of Primary Care Health Sciences University of Oxford<br><br>Dr Monique Andersson, Oxford University Hospital NHS Trust |

© Copyright: The University of Oxford and Oxford University Hospitals NHS Foundation Trust 2019

Dr Susan Hopkins, Incident Director for COVID-19, Public Health England

Dr Sarah Tonkin Crine, Department of Primary Care Health Sciences, University of Oxford

Dr Aleksandra Borek, Department of Primary Care Health Sciences, University of Oxford

Prof. Mahendra G Patel, Honorary Visiting Professor of Pharmacy University of Bradford , Honorary Senior Lecturer Academic Unit Primary Care Medical School University of Sheffield

**Sponsor:** University of Oxford
Joint Research Office
1st floor, Boundary Brook House
Churchill Drive,
Headington
Oxford
OX3 7GB

**Funder:** UKRI, DHSC, NIHR

**Chief Investigator Signature:**

**Statistician Signature:**

No potential conflict of interest

**Confidentiality Statement**
This document contains confidential information that must not be disclosed to anyone other than the Sponsor, the Investigator Team, HRA, host organisation, and members of the Research Ethics Committee and Regulatory Authorities unless authorised to do so.

See *supplementary material B* for **Key Trial Contacts**.

© Copyright: The University of Oxford and Oxford University Hospitals NHS Foundation Trust 2019

### Platform Randomised trial of INterventions against COVID-19 In older peoPLE (PRINCIPLE): Overview

**Background:** There is an urgent need to identify effective treatments for SARS-CoV-2 infection that helps people recover quicker and reduces the need for hospital admission. We have established an open, adaptive, platform trial to evaluate treatments suitable for use in the community for treating COVID-like-illness that might help people recover sooner and prevent hospitalisation.

**Eligibility and randomisation:** This protocol describes a randomised trial for people in the community at higher risk of an adverse outcome from possible or confirmed SARS-CoV-2 infection, defined in accordance with the United Kingdom's National Health Service syndromic case definition (https://www.nhs.uk/conditions/coronavirus-covid-19/symptoms/). Participants are randomised to receive either usual care or a trial treatment in addition to usual care (see Intervention Specific Appendices). Participants can take part in the study if they are eligible to be randomised to at least one intervention arm, as well as the Usual Care arm.

**Platform trial:** A "platform trial" is a trial in which multiple treatments for the same disease are tested simultaneously. New interventions can be added or replace existing ones during the course of the trial in accordance with pre-specified criteria.

**Response adaptive randomisation:** The initial randomisation ratio is fixed 1:1 for a comparison between two trial arms, but the trial has the capability for these proportions to be altered according to participants' responses to interventions. Pre-specified decision criteria allow for dropping a treatment for futility, declaring a treatment superior, or adding a new treatment to be tested. If at any point a treatment is deemed superior to the usual care arm, the superior treatment may replace the usual care arm as the new standard of care. In the context of the COVID-19 pandemic, the trial may continue as long as the pandemic persists.

**Outcomes:** The trial has co-primary endpoints: 1) Time taken to self-reported recovery from randomisation; and 2) hospitalisation and/or death. The main objective of the trial is to assess the effectiveness of the interventions in reducing time to recovery and in reducing the incidence of hospitalisation and/or death.

Key secondary outcomes include: Hospital assessment without admission; Oxygen administration; Intensive Care Unit admission; Mechanical ventilation (components of the WHO Clinical Progression Ordinal Scale); Duration of hospital admission; Duration of severe symptoms; Sustained recovery; Contacts with the health services; Consumption of antibiotics; Effects in those with a positive test for COVID-19 infection; WHO Well-being Index; daily rating of how well participant feels; Safety.

See *supplementary material C* for details of objectives and outcome measures.

**Efficient study design:** All enrolment (screening, informed consent, eligibility review and baseline data) and follow-up procedures (daily diary, data capture of hospitalisations and deaths) can be performed and captured online on the trial website or by telephone with a member of the trial

© Copyright: The University of Oxford and Oxford University Hospitals NHS Foundation Trust 2019

team. Randomisation is online and automatic following eligibility confirmation. Participant packs and medications are sent from the central study team directly to the participant.

**Data to be recorded:** We will capture demographic features including ethnicity and care home residency at baseline. In the online daily diary (completed for 28 days)/ during telephone calls, participants or their Study Partners will rate the severity of symptoms including how well they are feeling, record contacts with the health services (including hospital admission), record medication use, and new infections in the household. The WHO-5 Wellbeing Index, a five-question instrument, will assess wellbeing at baseline and on days 14 and 28. Follow-up beyond 28 days after randomisation will be by accessing electronic medical records and by participant questionnaire for information relevant to the longer term consequences of COVID-19.

**Numbers to be randomised:** The trial will continue until either superiority or futility is claimed for each intervention. We estimate that approximately 400 participants per arm (800 participants total if only a single intervention vs. Usual Care) will be required to provide 90% power for detecting an approximate difference of 2 days in median recovery time in the primary analysis population. We estimate that approximately 1500 participants per arm (3000 participants total if only a single intervention vs. usual care) will be required to provide 90% power for detecting a 50% reduction in the relative risk of hospitalisation and/or death in the primary analysis population.

**To enquire about the trial, contact the PRINCIPLE Trial Team:**

PRINCIPLE Trial
Nuffield Department of Primary Care Health Sciences
Radcliffe Primary Care
Radcliffe Observatory Quarter, Woodstock Road
Oxford
OX2 6GG

Website: www.principletrial.org

© Copyright: The University of Oxford and Oxford University Hospitals NHS Foundation Trust 2019

© Copyright: The University of Oxford and Oxford University Hospitals NHS Foundation Trust 2019

17 APPENDIX B: AMENDMENT HISTORY ... 39
18 APPENDIX C: USUAL CARE ARM ... 42
1. Background and rationale ... 42
2. Changes to outcome measures ... 42
3. Detail of intervention ... 42
a. Investigational Medicinal Product (IMP) description ... 42
b. Storage of IMP ... 42
4. Safety reporting ... 42
19 APPENDIX D: USUAL CARE PLUS HYDROXYCHLOROQUINE ARM (DISCONTINUED) ... 43
1. Background and rationale ... 43
a. Evidence for potential Hydroxychloroquine benefits in COVID-19 ... 43
2. Eligibility criteria specifically related to hydroxychloroquine ... 47
3. Outcome measures related to hydroxychloroquine ... 48
4. Detail of intervention ... 48
a. Investigational Medicinal Product (IMP) description ... 48
b. Storage of IMP ... 48
c. SmPC precautions and concomitant medication ... 49
*i.* Precautions ... 49
ii. Concomitant medication ... 49
iii. Pregnancy and Breastfeeding ... 50
5. Safety reporting ... 50
20 APPENDIX E: USUAL CARE PLUS AZITHROMYCIN ARM (DISCONTINUED) ... 51
1. Background and rationale ... 51
a. Evidence for potential Azithromycin benefits in COVID-19 ... 51
b. Importance of treating CAP or CAP risk in the elderly or immuno-compromised ... 51
2 Changes to outcome measures ... 52
3 Eligibility criteria specifically related to azithromycin ... 52
4 Detail of intervention ... 52
a. Investigational Medicinal Product (IMP) description ... 53
b. Storage of IMP ... 53
c. SmPC precautions and concomitant medication ... 53
i. Precautions ... 53
ii. Concomitant medications ... 53
iii. Fertility, pregnancy and lactation ... 57
5 Safety reporting ... 57
21 APPENDIX F: USUAL CARE PLUS DOXYCYCLINE ARM (DISCONTINUED) ... 58
1. Background and rationale ... 58

© Copyright: The University of Oxford and Oxford University Hospitals NHS Foundation Trust 2019

© Copyright: The University of Oxford and Oxford University Hospitals NHS Foundation Trust 2019

### 1. BACKGROUND and RATIONALE

We urgently need to know whether potential treatments for COVID-19-like-illness that are suitable for use in the community might help affected individuals recover more quickly and reduce the risk of hospitalisation and/or death.(1) PRINCIPLE is a platform trial designed to efficiently

© Copyright: The University of Oxford and Oxford University Hospitals NHS Foundation Trust 2019

evaluate potential treatments for people with COVID-19-like-illness in the community, and who may be at higher risk of poorer outcomes. Eligible participants are those who meet the United Kingdom's National Health Service syndromic case definition (https://www.nhs.uk/conditions/coronavirus-covid-19/symptoms/), who are being managed in the community, and who are aged 65 and over, or 18 to 64 and experiencing shortness of breath as part of COVID-19 illness, or aged 18-64 with certain comorbidities (2-6)

The platform trial has the flexibility to allow additional interventions to be added in, or to replace existing interventions according to pre-specified criteria. If at any point a treatment is deemed superior to the usual care arm, the superior treatment may replace the usual care arm as the new standard of care. All approved intervention arms are outlined in Intervention Specific Appendices (ISAs).

The trial has co-primary endpoints: 1) Time taken to self-reported recovery from randomisation; and 2) Hospitalisation and/or death. The main objective of the trial is to assess the effectiveness of the respective interventions in reducing time to recovery and in reducing the incidence of hospitalisation and/or death.

The primary analysis will include all participants as specified in the master statistical analysis plan and the adaptive design report. Clinical data, and information from swab and blood tests, where available, will be used to classify participants according to aetiology.

#### 2. TRIAL DESIGN AND PROCEDURES

PRINCIPLE is an open, prospective, individually randomised, platform, response adaptive, controlled clinical trial in community care.

##### 2.1 Participant Identification

###### 2.1.1 Trial Participants

The trial aims to include symptomatic participants with confirmed, or possible COVID-19 who meet the current NHS case definition for possible COVID-19, and who are well enough to remain in the community. This definition can be found here: https://www.nhs.uk/conditions/coronavirus-covid-19/symptoms/. Participants must be aged 65 and over, **OR** aged 18 to 64 and experiencing shortness of breath as part of COVID-19 illness, **OR** aged 18-64 with certain comorbidities.

Participants experiencing shortness of breath have a greater risk of severe and critical disease outcomes with COVID-19 (5).

The study is for people who have ongoing symptoms.

###### 2.1.2 Inclusion Criteria

Inclusion requires the following:

© Copyright: The University of Oxford and Oxford University Hospitals NHS Foundation Trust 2019

1. Participant or their legal representative, is willing and able to give informed consent for participation in the study, and is willing to comply with all trial procedures

2. Suspected COVID-19 using the NHS syndromic definition, OR symptoms consistent with COVID-19* and with a positive test for SARS-CoV-2 infection within the past 14 days

3. Symptoms must have started within the past 14 days and be ongoing

**AND**

4. Participant is aged 65 or over **OR**

Participant is aged 18- 64, and is experiencing shortness of breath as part of COVID-19 illness **OR**

Participant is aged 18-64 and has any of the following underlying health conditions

a) Known weakened immune system due to a serious illness or medication (e.g. chemotherapy);
b) Known heart disease and/or a diagnosis of high blood pressure
c) Known chronic lung disease (e.g. asthma)
d) Known diabetes
e) Known mild hepatic impairment;
f) Known stroke or neurological problem;
g) Self-report obesity or body mass index ≥35 $kg/m^2$

*These symptoms may include, but are not limited to, shortness of breath, general feeling of being unwell, muscle pain, diarrhoea and vomiting.

##### 2.1.3 Exclusion Criteria

- Patient currently admitted in hospital
- Almost recovered (generally much improved and symptoms now mild or almost absent)
- Judgement of the recruiting clinician deems ineligible.
- Previous randomisation to an arm of the PRINCIPLE trial

Additional exclusions specific to each intervention arm are listed in the ISAs. For participation, participants must be eligible to be randomised to at least one intervention arm as well as the Usual Care arm.

#### 2.2 Trial procedures

##### 2.2.1 Recruitment

Recruitment is possible through a variety of mechanisms:

##### 2.2.2 Face to face

Attending clinicians including video consultations, including research nurses or other health care professionals, at general medical practices, paramedic services, hospital emergency departments,

© Copyright: The University of Oxford and Oxford University Hospitals NHS Foundation Trust 2019

clinical care hubs, Hospital at Home facilities, care of the elderly services, pharmacies, social care services, residential and nursing homes, or any health and social care facility, can facilitate recruitment into the trial. They can do this by discussing the study with potentially eligible participants, guiding them through informed consent procedures, collection of baseline data, completion of screening questions, collecting information for eligibility assessment, and randomising the participant. If required and appropriate, licensed prescribers may prescribe the medication appropriate to the group to which the participant is randomised. Alternatively, health care professionals may revert to the PC-CTU to complete the activity, including eligibility confirmation and issue of study medication and materials.

##### 2.2.3 Remote recruitment

i) All Health, health related, and Social Care professionals will be able to give information verbally or via a trial text, email, poster, social media post, adverts, media release, leaflet or letter, to potential study participants and their study partners. They may also direct patients to the online study information and the study website.

ii) Potential participants may present directly to the study team via the website or by the study telephone contact. The study team can provide information about joining the trial and guide them through the consent and enrolment process.

iii) A General Practice may be contacted by a potential participant or the practice may contact patients, by text (or by letter), who may match the trial eligibility criteria, through running searches of their database. They will then direct patients to the trial enrolment website or seek verbal consent to be contacted by the trial team.

iv) NHS Digital will provide the PRINCIPLE trial with a daily list of contact details from the COVID-19 testing Pillar 2 data, for patients receiving a positive test result for SARS-Co-V2 infection, via a secure transfer system. NHS Digital will apply an age filter to ensure only the details of those within the age range of the trial are passed on to PRINCIPLE. The trial team will make a limited number (maximum of 3) of attempts to telephone, text or email these patients to provide them with information about the trial, to invite them to consider taking part, and to enrol them if they provide full informed consent and are deemed eligible at screening.

Patient details will be provided in accordance with section 251 under the General Notice under the Health Service Control of Patient Information Regulations 2002, which applies only in England and Wales, providing patient information without consent for COVID-19 public health, surveillance and research purposes. The notice provides a temporary legal basis to avoid a breach of confidentiality for COVID-19 purposes.

v) Join Dementia Research (JDR) - We will also be using JDR as a recruitment tool. This is an on-line self-registration service that enables volunteers with memory problems or dementia, carers of those with memory problems or dementia and healthy volunteers to register their interest in taking part in research.

For all recruitment models:

- Study Partner: at screening the potential participant will be asked to provide contact details for a Study Partner, to assist in completing trial procedures and to provide

© Copyright: The University of Oxford and Oxford University Hospitals NHS Foundation Trust 2019

information on their behalf where necessary, but this is not a requirement for trial participation. However, it is strongly encouraged that participants who may be frailer and/or lack capacity to consent make use of a study partner to facilitate their participation. In addition to family member or friend, the study partner may also be a carer or other suitable person.

- Participants may be asked if they wish to enrol in additional studies that do not conflict with the main PRINCIPLE trial. Those who do not screen as eligible for PRINCIPLE may be alerted to the possibility of participating in other approved trials.

#### 2.3 Screening

An online screening, eligibility and consent procedure is used. If online access is not possible, a member of the trial team collects this information during a telephone call. A trial free-phone number enables participants to contact the trial team for further information and study participation support. Participants are screened after they have read the PIS by completing an online eligibility questionnaire.

#### 2.4 Informed Consent

If participants meet the screening criteria, they will be asked to provide informed consent and a screening trial ID number will be assigned to them. Remote, paperless online/telephone consent is required, and appropriate during the pandemic. Participants will be able to download their consent form, or it may be printed by the central study team and delivered to participants with their study materials if they so prefer.

Written and summary versions of the PIS and ICF will be presented to participants detailing no less than: the exact nature of the trial; the known side-effects and risks involved in taking part. It will be clear that the participant is free to withdraw from the study at any time. A summary, pictorial PIS is available which can be read by those feeling very unwell, lack capacity or have low reading comprehension skills. Adequate time will be given to the participant to consider the information and to ask any questions about the trial before deciding whether to participate. After consent, the participant will enter online baseline information, including their address, contact details and those of a Study Partner.

Population groups such as care home residents have been amongst those hardest hit by the pandemic and therefore stand to benefit the most from any effective treatments. However, some care home residents lack capacity to consent to research themselves. If the recruiting clinician deems a care home resident lacks capacity to consent then a personal or professional legal representative (England and Wales only) will be asked to provide consent for those lacking capacity to consent for themselves. A personal legal representative is defined as a person not connected with the conduct of the trial who is suitable to act as the legal representative by virtue of their relationship with the adult. A professional legal representative may be a doctor responsible for the medical treatment of the adult if they are independent of the study, or a person nominated by the healthcare provider. In all instances, a personal legal representative will be sought first and a professional legal representative sought only if a personal legal representative cannot be identified. A professional legal representative will be sought in order not to deny access to research to older adults who may not have personal legal representatives.

© Copyright: The University of Oxford and Oxford University Hospitals NHS Foundation Trust 2019

Only residents of care homes who lack capacity to consent will be recruited, adults who lack capacity to consent will not be recruited from the wider community. Legal Guardians and recruiting clinicians will not endeavour to obtain consent for or recruit into the trial residents who, in addition to their lack of capacity, have a quality of life which can reasonably be considered as not acceptable to the potential participant.

The legal representative will be provided with information about the trial and made aware of the following:

- They are being asked to give consent on behalf of the incapacitated adult,
- They are free to decide whether they wish to make this decision or not, and
- They are being asked to consider what the adult would want, and to set aside their own personal views when making this decision.

#### 2.5 Eligibility Assessment

Eligibility of those who have provided appropriate consent can be checked at study sites or centrally by a medically qualified clinician or a research nurse, who is suitably trained and experienced and has been delegated this responsibility, and who has appropriate access to the participant's summary care record or relevant medical information. If a participant's summary care record cannot be accessed centrally, the clinician/delegate will contact the participant's primary care medical practice for information relevant to confirming eligibility. Participants will not be randomised to an arm if an exclusion criterion to that arm applies to them, but will need to have no exclusions relevant to at least one intervention and the usual care arm.

#### 2.6 Randomisation

Participants will be randomised using a fully validated and compliant web-based randomisation system called Sortition. Once deemed eligible, the clinician or a member of the trial team will randomise the participant, to one of the arms they are eligible for (at least two arms, usual care and at least one intervention), automatically by Sortition. Full details of response adaptive randomisation are described in section 5.2.2.

The participant, legal representative if applicable, trial team and participant's GP will be notified electronically of the treatment allocation. If the participant does not have an email address, they will be notified by telephone. The research team may also send the GP or Care Home an email or letter via secure systems, containing personally identifiable data and treatment allocation.

#### 2.7 Blinding and code-breaking

PRINCIPLE is an open-label trial. The participant, legal representative if applicable, and the recruiting clinician will know the participant's allocation. Therefore, no unblinding or code breaking is required. However, those managing the data will be blind to participant allocation.

© Copyright: The University of Oxford and Oxford University Hospitals NHS Foundation Trust 2019

The trial team and recruiting clinicians will be blinded to emerging results. During the course of the trial, only the unblinding statisticians and the independent members of the Data Monitoring and Safety Committee will have access to the unblinded interim results.

#### 2.8 Baseline Assessments

Once randomised, study medication (if so randomised), and a participant pack will be sent to participants, from their general practice, study team, Public Health England (PHE) or other approved central service (or collected from a general practice or pharmacy). Participants may be offered a swab test as part of standard care. Where possible, and availability of sampling kits allows, one sample will be taken as close to study entry as possible to assess COVID-19 status and other viral aetiologies. While the aim is to have a swab result for all patients, where swabs are unavailable, patients may still participate and be included in the primary intention to treat analysis only.

#### 2.9 Subsequent Visits

There is no requirement for participants to have a face-to-face visit as part of trial participation. Those participants randomised to an unlicensed medication will receive a call from the study team within one day of randomisation, to reaffirm consent, to explain when to call the 24 hour safety phone line, what to do in an emergency, and to answer any other trial questions. This information is also included in the participant information sheet and the participation pack that they will receive. All subsequent measurements consist of self-completed questionnaires online or through telephone calls, and primary care and/or hospital record searches. We will ascertain relevant data from primary care and/or hospital medical records about length of hospital stay, oxygen therapy, and ICU admission and ventilation, if applicable.

Participants will be sent a link to their online diary, which they will be asked to complete for 28 days. They will be asked to rate the severity of symptoms, record contacts with the health services (including hospital admission), record medication use and new infections in the household. It is becoming increasingly apparent the COVID-19 infection may have a considerable negative impact on well-being (7) and so the five questions of WHO-5, validated for measuring wellbeing over time, will be presented at baseline and on days 14 and 28. We will not ask for WHO-5 questions to be completed for participants who lack capacity. We will capture ethnicity and care home residency at baseline and day 28 (if missed at baseline).

All participants receive a call from the trial team on day 2/3 to reaffirm consent, to confirm that they have received a participant pack, and trial medication (if randomised to a trial medication), and to explain that they should complete the daily diary for 28 days even if they feel better or their swab result is negative. The trial team calls participants/study partners on days 7, 14 and 28 if they do not have internet access or have *not* completed their diary for at least 2 consecutive days prior to the call. No more than six contact attempts will be made at each of these follow-up points. For those on unlicensed medication, if the participant/study partner does not answer the calls and hasn't completed online diaries, their GP will be contacted to allow us to monitor any potential side-effects associated with the medication.

© Copyright: The University of Oxford and Oxford University Hospitals NHS Foundation Trust 2019

We will seek consent from participants to contact them on a monthly basis for up to 12 months after enrolment (via email, text message or phone call) to collect information about any ongoing symptoms, hospitalisations and well-being. We will re-consent those already enrolled in the trial.

In addition to the swab being undertaken as part of the national RCGP RSC surveillance programme with PHE, trial participants will also be asked to consent to the trial team accessing a blood sample result. The study team will obtain the result from RCGP RSC/PHE.

The RCGP RSC will report to the central trial office at regular intervals about healthcare contacts in the participant’s clinical records, as they are able to download this information centrally. This will be used as confirmation and a back-up for information obtained directly from study participants and other data sources outlined above. If obtaining data is not possible using this route, the GP surgery will be contacted to request a limited notes review. Participant records will be accessed up to twelve months following enrolment to ascertain follow up data from enrolment to day 28. Data will be collected as close to real time as possible; RCGP RSC, EMIS and NHS Digital and other sources of routinely collected data will be utilised if required To investigate the impact of trial interventions on the longer-term effects of COVID-19, we will use these data collection methods to follow-up participants, for up to 10 years.

##### 2.10 Qualitative Sub-study

A qualitative sub-study will be nested within the trial, to capture data to understand how patients conceptualise their illness and how they respond to taking medication(s) as part of the trial. Once participants have completed the trial, we will interview their respective clinicians to explore their views of taking part in trials during a pandemic. Healthcare professionals will also be asked about their experiences of taking part in the trial. See *supplementary material F* for further details. Participants who lack capacity will not be invited to participate in the qualitative sub study.

##### 2.11 Early Discontinuation/Withdrawal of Participants

Each participant, or their legal representative on the participant’s behalf, has the right to withdraw from the study at any time. For those that lack capacity, expression of dissent in any form will be taken as an indication they do not wish to be included and they will be withdrawn. In addition, the Investigator may discontinue a participant from the study at any time if the Investigator considers it necessary for any reason including:

- Ineligibility (either arising during the study or retrospectively having been overlooked at screening)
- Withdrawal of consent

The reason for withdrawal will be recorded on the CRF. Data that has already been collected about the participant will be kept and used.

##### 2.12 Definition of End of Trial

Last data capture of last participant, when: no further suitable interventions are available and/or COVID-19 is no longer prevalent. March 2022 has been decided as the formal end date at this stage, but this date may need to be amended depending on circumstances prevailing at the time.

© Copyright: The University of Oxford and Oxford University Hospitals NHS Foundation Trust 2019

### 3 TRIAL INTERVENTIONS

IMP information can be found in the relevant ISAs.

In general, re-packaging and issuing of medication can be completed by: the patient's registered GP surgery or treatment and assessment facility; an accredited licensed central facility; an online, community or hospital pharmacy, and The Primary Care Clinical Trials Unit (as approved by the MHRA). Distribution of trial packs to participants will be tracked via courier or call/text message. Clinicians can prescribe trial medications that can be issued in the community and pharmacies can issue medication to the patient by community pharmacy services 'on-line pharmacy' services, NHS volunteers, or it can be collected from the pharmacy by the participant or someone on their behalf.

To record presence of symptoms and severity, as well as adherence to trial treatment, participants will receive a daily email asking them to complete an online diary where they will record their symptoms and medicines use. If incomplete, the trial team will contact the participant and/or their Study Partner to obtain the data. Section 2.9 explains the additional oversight of those participants receiving unlicensed medication. A risk-adapted approach will be used for drug accountability. Accountability logs will be kept by PC-CTU when they ship drug.

### 4 SAFETY REPORTING

All symptoms, medication side-effects and SAEs will be collected from participant daily diaries, calls to participants/Study Partners, medical records, notes reviews and RCGP data downloads. SAE information will be analysed as part of the interim and whole trial analysis and will be reviewed at each Data Safety & Monitoring Committee meeting.

#### 4.1 Procedures for Reporting Adverse Events and Serious Adverse Events

The severity of events and symptoms will be assessed by participants in daily diaries on the following scale: no problem/mild problem/moderate problem/major problem.

| | Participant reported symptom rating |
|---|---|
| No problem | Symptom not experienced |
| Mild problem | Short-lived or mild symptoms; medication may be required. No limitation to usual activity |
| Moderate problem | Moderate limitation in usual activity. Medication may be required. |
| Major problem | Considerable limitation in activity. Medication or medical attention required. |

i. **AE reporting**

AEs will be monitored daily for participants allocated to either hydroxychloroquine or treatments not licensed in the UK, whilst they are taking the drug, to allow successful safety monitoring of

© Copyright: The University of Oxford and Oxford University Hospitals NHS Foundation Trust 2019

these less familiar treatments. Participants will be free to withdraw from taking the treatment if they perceive they have an intolerable AE.

For each treatment not licensed in the UK, the following AEs from the start of medication until the specified follow-up period, will be assessed by a clinician for causality and severity (definitions below): i) pre-defined AEs detailed in the ISA that are rated by the participant as 'moderate' and ii) other reported 'major' AEs.

Participants will also be provided with a Trial Wallet Emergency Card detailing potential side-effects and a 24-hour contact telephone line, manned by a clinical team, enabling them to report any moderate or major AEs they experience whilst taking the drug. The clinician will contact the participant directly within 24 hrs of becoming aware of a major AE reported in their daily diary or on the Freephone number, to advise the participant on the appropriate clinical care, as well as notifying the participant's GP about the event. In the event of a medical emergency, trial participants will be instructed to show this card to the clinician they see.

##### ii. AE Severity Assessment (for assessing clinician):

| | Clinical assessment of severity |
|---|---|
| **GRADE 1 (Mild)** | Short-lived or mild symptoms; medication may be required. No limitation to usual activity |
| **GRADE 2 (Moderate)** | Moderate limitation in usual activity. Medication may be required. |
| **GRADE 3 (Severe)** | Considerable limitation in activity. Medication or medical attention required. |

##### iii. SAEs

Hospitalisation and/or death due to confirmed or possible SARS-Cov-2 infection is a primary outcome, we will collect this data using a risk-adapted approach and will not report such SAEs. SAEs other than hospitalisation or death due to COVID-19 must be reported for all treatments.

SAEs must be reported by the person who has discovered the SAE or nominated delegate within 24 hours of becoming aware of the event. The sponsor or delegate will ensure it is reviewed by the CI or other delegated personnel for relatedness and expectedness as soon as possible taking into account the reporting time for a potential SUSAR according to the relevant competent authority. If the event has not resolved, at the 28 day time point the SAE will be reviewed again to see if resolution has occurred. If the event is considered 'resolved' or 'resolving' no further follow up is required. If not, the event must be followed up until such a time point.

All SAEs that have not resolved by the end of the study, or that have not resolved upon discontinuation of the participant's participation in the study, must be followed until any of the following occurs:

- The event resolves
- The event stabilises
- The event returns to "baseline", if a "baseline" value/status is available

© Copyright: The University of Oxford and Oxford University Hospitals NHS Foundation Trust 2019

• The event can be attributed to agents other than the study intervention or to factors unrelated to study conduct

• It becomes unlikely that any additional information can be obtained (participant or health care practitioner refusal to provide additional information, lost to follow-up after demonstration of due diligence with follow-up efforts)

*See Supplementary Material D for definitions of adverse events*

###### 4.1.1. Other events exempt from immediate reporting as SAEs

Hospitalisations will be defined as at least a one night admission to hospital. Hospitalisation for a pre-existing condition, including elective procedures planned prior to study entry, which has not worsened, does not constitute an SAE, and standard supportive care for the disease under study are not SAEs and do not require SAE reporting.

###### 4.1.2. Procedure for immediate reporting of Serious Adverse Events

- Trial team will complete an SAE report form for all reportable SAEs.
- GP practice/trial team/RCGP will provide additional, missing or follow up information in a timely fashion.
- If necessary the participant may be contacted to provide additional, missing or follow up information as required.

The CI or delegate will review the SAE once reported, collect as much information and report to the Sponsor within the timeframe according to the PC-CTU SOPs.

###### 4.1.3 Expectedness and Causality

For SAEs that require reporting, expectedness of SARs will be determined according to the relevant RSI section of the Summary of Product Characteristics/IB. The RSI will be the current Sponsor and MHRA approved version at the time of the event occurrence.

##### Assessment of Causality

The relationship of each serious adverse event to the trial medication must be determined by a medically qualified individual according to the following definitions:

- **Unrelated** – where an event is not considered to be related to the IMP
- **Possibly** – although a relationship to the IMP cannot be completely ruled out, the nature of the event, the underlying disease, concomitant medication or temporal relationship make other explanations possible.
- **Probably** – the temporal relationship and absence of a more likely explanation suggest the event could be related to the IMP.
- **Definitely** – the known effects of the IMP, its therapeutic class or based on challenge testing suggest that the IMP is the most likely cause.

© Copyright: The University of Oxford and Oxford University Hospitals NHS Foundation Trust 2019

All AEs/SAEs labelled possibly, probably or definitely will be considered as related to the IMP.

##### 4.2 SUSAR Reporting

All SUSARs will be reported by the sponsor delegate to the relevant Competent Authority and to the REC and other parties as applicable. For fatal and life-threatening SUSARS, this will be done no later than seven calendar days after the Sponsor or delegate is first aware of the reaction. Any additional relevant information will be reported within eight calendar days of the initial report. All other SUSARs will be reported within 15 calendar days.

Principal Investigators will be informed of all SUSARs for the relevant IMP for all studies with the same Sponsor, whether or not the event occurred in the current trial.

##### 4.3 Development Safety Update Reports

The DSUR will be developed and submitted annually on the anniversary date that the trial receives Clinical Trial Authorisation +60 days. Due to the nature of this trial and the importance of sharing the science of COVID-19 and the drug, internationally, we expect to produce reports to the UK Government and regulatory agency more frequently upon request.

#### 5 STATISTICS

##### 5.1 Master Statistical Analysis Plan (M-SAP)

Details of the statistical design and methods will be described in a Master Statistical Analysis Plan (M-SAP), in which an appendix to the M-SAP titled “Adaptive Design Report” (ADR) provides complete specifications for the primary analysis and pre-specified adaptive algorithm. In addition, the M-SAP will be accompanied by arm-specific appendices to describe any planned deviations from the M-SAP. A broad overview of the design and primary analyses is provided below.

##### 5.2 Open Adaptive Platform Trial

PRINCIPLE is an open, adaptive, platform trial to evaluate emerging treatments for symptomatic COVID-19-like illness. A “platform trial” is a trial in which multiple treatments for the same disease are tested simultaneously. The backbone of the trial is an adaptive clinical trial design. Pre-specified decision criteria allow for dropping a treatment for futility, declaring a treatment superior, or adding a new treatment to be tested. If at any point a treatment is deemed superior to the Usual Care arm, the superior treatment may replace the Usual Care arm as the new standard of care. Because the process of dropping and adding treatments may be on-going for an indefinite period of time, platform trials may be better conceived of as a process rather than a singular clinical trial. In the context of the COVID-19 pandemic, the trial may continue as long as the pandemic persists.

The PRINCIPLE trial will begin as a two arm, 1:1 randomised trial but will have the capability to add additional interventions over time. The evaluation of any new interventions will be governed by this master protocol and M-SAP (including adaptive algorithm and decision criteria), with any planned deviations from the master protocol and M-SAP to be specified in arm-specific

© Copyright: The University of Oxford and Oxford University Hospitals NHS Foundation Trust 2019

appendices. The inclusion of any new interventions will require additional arm-specific appendices to the master protocol and M-SAP.

##### 5.2.1 Co-Primary Endpoints & Analyses

There are two co-primary endpoints. The first co-primary endpoint is time to recovery from possible COVID-19 infection within 28 days from randomisation, where time to recovery is defined as the first instance that a participant reports feeling recovered. The second co-primary endpoint is hospital admission or death related to possible or confirmed COVID-19 within 28 days from randomisation. Unless otherwise specified in the Intervention Specific Appendices (ISA), the co-primary outcomes will be evaluated using a "gate-keeping" strategy. For a given treatment, the hypothesis for the time to recovery endpoint will be evaluated first, and if the recovery null hypothesis is rejected, the hypothesis for the second co-primary endpoint of hospitalisation/death will be evaluated. This gate-keeping strategy preserves the overall Type I error of the primary endpoints without additional adjustments for multiple hypotheses. In addition, the gate-keeping structure reflects the clinical belief that an intervention is unlikely to demonstrate benefit on the hospitalisation/death endpoint without first demonstrating benefit on the time to recovery endpoint.

The primary outcome of time to recovery is defined as the first instance that a participant reports feeling recovered. The corresponding primary analysis for this outcome is a Bayesian piecewise exponential model, with time to recovery regressed on treatment and stratification covariates (age, comorbidity). Let $\theta_j$ denote the log hazards ratio comparing the hazards of recovery for participants in treatment group *j* versus participants in the Usual Care arm. A corresponding Bayesian posterior distribution will be derived for the estimated log hazards ratio. The first co-primary analysis for intervention *j* will test the following hypothesis:

$$H_{10}: \theta_j \leq 0$$
$$H_{11}: \theta_j > 0$$

If the Bayesian posterior probability of superiority (a log hazards ratio greater than 0 corresponding to quicker recovery) for a treatment versus Usual Care is sufficiently large (e.g. ≥ 0.99), the null hypothesis will be rejected and the intervention will be deemed superior to Usual Care with respect to time to recovery. The exact threshold of the superiority decision criterion (e.g. 0.99) will be determined *a priori* via simulation to control the one-sided Type I error of the study at approximately 0.025, and will be specified in the Adaptive Design Report (Appendix to the M-SAP). The Adaptive Design Report will also specify appropriate methodology for the primary analysis when the Usual Care arm is replaced by a superior treatment, and for when the comparison of a treatment versus Usual Care includes non-concurrent randomisations.

The second co-primary endpoint is hospital admission or death due to possible SARS-CoV-2 infection. The corresponding analysis will be a Bayesian generalised linear model of hospitalisation/death regressed on treatment and stratification covariates (age, comorbidity). Let $\delta_j$ denote the log odds ratio comparing the odds of hospitalisation/death for persons in treatment group *j* versus persons in the Usual Care arm. A corresponding Bayesian posterior distribution will be derived for the estimated log odds ratio. If the first co-primary endpoint hypothesis (for time to recovery) is rejected for intervention *j*, the second co-primary hypothesis for intervention *j* be tested:

© Copyright: The University of Oxford and Oxford University Hospitals NHS Foundation Trust 2019

$$H_{20}: \delta_j \leq 0$$
$$H_{21}: \delta_j > 0$$

If the Bayesian posterior probability of superiority on hospitalisation/death for a treatment versus Usual Care is sufficiently large (e.g. $\geq 0.99$), the null hypothesis will be rejected and the intervention will be deemed superior to Usual Care with respect to hospitalisation/death. The exact threshold of the superiority decision criterion (e.g. 0.99) will be determined *a priori* via simulation to control the one-sided Type I error of the study at approximately 0.025, and will be specified in the M-SAP.

##### 5.2.2 Adaptive Design

The pre-specified design will allow adaptations to the trial based on the observed co-primary endpoint data. These adaptations include the declaration of success or futility of an intervention at an interim analysis, the addition or removal of treatment arms, and changes in the randomisation probabilities. Adaptations will occur at a given interim analysis if pre-specified conditions are satisfied. The adaptive algorithm will be documented in the Adaptive Design Report, including pre-specified criteria for decisions regarding futility or effectiveness of interventions and/or replacing interventions in the trial.

##### 5.2.3 Interim Analyses

Precise timing of the first interim analysis and frequency of subsequent interim analyses will be specified in the Adaptive Design Report, based on both simulations and logistical considerations. At each interim analysis, all enrolled intervention arms will be evaluated for success and futility on both co-primary endpoints using the Bayesian primary analyses. These interim analyses will maintain the gate-keeping sequential order by first evaluating the hypothesis for time to recovery, and if the recovery endpoint null hypothesis is rejected, subsequently evaluating the hypothesis for hospitalisation and/or death. If the Bayesian posterior probability of superiority of a given intervention versus Usual Care is sufficiently large for a given endpoint (e.g. $\geq 0.99$) within the gate-keeping structure, superiority will be declared versus Usual Care with respect to that endpoint.

If the Bayesian posterior probability of a clinically meaningful treatment effect is sufficiently small (e.g. $< 0.01$) for the first co-primary endpoint (time to recovery), the intervention arm may be dropped from the study for futility. If there are no other intervention arms available, the trial may be suspended; otherwise accrual continues to the remaining treatment arms. The exact futility thresholds will be pre-specified in the Adaptive Design Report and determined via simulation.

##### 5.2.4 Allocation & Response Adaptive Randomisation

Initially, randomisation will be fixed 1:1 for a comparison between two trial arms, with stratification by age (less than 65, greater than or equal to 65), and comorbidity (yes/no). If a second experimental intervention arm is added to the study, randomisation allocation will be modified and the additional intervention will be included in the interim analyses (with evaluation for success and futility) as detailed in the Adaptive Design Report. If there are at least 3 arms (2 intervention arms plus Usual Care) in the study, each interim analysis may incorporate modified

© Copyright: The University of Oxford and Oxford University Hospitals NHS Foundation Trust 2019

randomisation probabilities via response adaptive randomisation (RAR). Full details for implementing RAR will be provided in the Adaptive Design Report; the general idea is to allocate more participants to the intervention arms that have the best observed outcomes. Except for the CTU programmer, the rest of the trial team are blinded to the RAR ratios.

##### 5.2.5 Sample Size Justification

Given the open perpetual trial structure, the trial does not have a finite ending based on sample size. Rather, the trial will continue until either superiority or futility is claimed for each intervention, or until the pandemic expires in the population. We estimate that approximately 400 participants per arm (800 participants total if only a single intervention vs. Usual Care) will be required to provide 90% power for detecting a hazard ratio of 1.3 (approximate difference of 2 days in median recovery time). This calculation is based on the assumption of an exponential distribution for time to recovery with a median of 9 days in the Usual Care arm, with some adjustments for missing data and multiple interim analyses. On average, we expect fewer participants to be required when there is a large treatment benefit or complete lack of benefit. For example, if the true hazard ratio is 1.5 (3 day benefit in median time to recovery), on average only 150 subjects per arm are required to provide sufficient power. The primary advantage of the adaptive design is the ability to adapt the sample size to the observed data, thus addressing the primary hypothesis as quickly and as efficiently as possible.

We estimate that approximately 1500 participants per arm (3000 participants total if only a single intervention vs. usual care) will be required to provide 90% power for detecting a 50% reduction in the relative risk of hospitalisation and/or death. This calculation is based on the assumption of an underlying 5% combined hospitalisation and/or death rate in the Usual Care arm, with an intervention lowering the hospitalisation and/or death rate to 2.5%, with some adjustments for the multiple interim analyses. We expect fewer participants to be required to detect a 50% reduction if the event rate in the Usual Care arm is greater than 5%.

##### 5.2.6 Virtual Trial Simulations

Because of the adaptive platform trial structure, there exists no simple formula(s) to calculate power and Type I error (false positive rate). Hence, virtual trial simulations will be used to fully characterize and quantify the power and Type I error of the design. These simulations will be conducted prior to the first interim analysis (with results described in the Adaptive Design Report), and will be used to optimize the adaptive decision criterion and RAR parameters. The simulations will include a comprehensive evaluation of trial performance across a wide range of assumptions (e.g. underlying distribution of outcome in Usual Care arm, treatment effect, accrual rates, etc.). This will include summaries regarding the number of subjects required to make a success or futility conclusions for each intervention. Complete details of the simulations will be provided in the Adaptive Design Report.

##### 5.2.7 Procedure for Accounting for Missing, Unused, and Spurious Data.

Full details of handling missing data will be specified in the M-SAP.

#### 5.3 Primary Analysis Population

© Copyright: The University of Oxford and Oxford University Hospitals NHS Foundation Trust 2019

The primary analysis population is defined as all randomised participants with a COVID-19 positive test, according to the groups they were randomly allocated to as specified in the M-SAP. All other analysis populations will be defined in the M-SAP.

##### 5.4 Procedures for Reporting Unplanned Deviation(s) from the Master Statistical Analysis Plan

Analyses will be carried out in accordance with the M-SAP and corresponding appendices. Any additional analysis that is not specified in the M-SAP/appendices or any unplanned deviation(s) from the M-SAP/appendices will be specified in the Statistical Report. Reasons for these changes will be documented and authorised by the Chief Investigator.

##### 5.5 Qualitative sub-study analysis

Audio-recordings of interviews will be transcribed verbatim and transcripts analysed using thematic analysis. Patient and HCP interviews transcripts will be analysed separately but findings will be compared and triangulated if deemed appropriate. Thematic analysis allows the research team to take a pragmatic approach to data collection, remaining grounded in the data but ensuring that the analysis answers the research objectives. NVivo software will be used to assist with the organisation and coding of data. Codes will be compared with one another to create categories, grouping similar codes together. A thematic framework will be developed to code all data and represent key themes for both sets of interviews.

#### 6 DATA MANAGEMENT

The data management aspects of the study are summarised here with details fully described in the Data Management Plan.

##### 6.1 Source Data

Source documents are where data are first recorded. These include, but are not limited to, hospital records (from which medical history and previous and concurrent medication may be summarised into the CRF), clinical and office charts, laboratory and pharmacy records, diaries, microfiches, radiographs, and correspondence.

If a participant fails to complete data online and after six attempts at contacting the participant/Study Partner, the RCGP RSC may be utilised to obtain missing data. Data collected will include participant identifiable information and will be accessed at the University of Oxford according to PC-CTU Information Governance policies and GDPR. Data will only be held for the duration it is required, this will be reviewed annually.

CRF entries will be considered source data if the CRF is the site of the original recording (e.g. there is no other written or electronic record of data). All documents will be stored safely in confidential conditions. On all study-specific documents, other than the signed consent, the participant will be referred to by the study participant number/code, not by name.

##### 6.2 Access to Data

© Copyright: The University of Oxford and Oxford University Hospitals NHS Foundation Trust 2019

Direct access will be granted to authorised representatives from the Sponsor and host institution for monitoring and/or audit of the study to ensure compliance with regulations.

##### 6.3 Data Recording and Record Keeping

In accordance with the principles of Good Clinical Practice and the recommendations and guidelines issued by regulatory agencies, the design, conduct and analysis of this trial is focussed on issues that might have a material impact on the wellbeing and safety of study participants (in the community with suspected or confirmed SARS-CoV-2 infection) and the reliability of the results that would inform the care for future patients.

The critical factors that influence the ability to deliver these quality objectives are:

- to minimise the burden on thousands of busy clinicians working in an overstretched primary care setting and undertaking research during a major epidemic
- to ensure that suitable patients have access to the trial medication
- to provide information on the study to patients and clinicians in a timely and readily digestible fashion but without impacting adversely on other aspects of the trial or the patient's care
- to collect comprehensive information on the mortality and disease status

In assessing any risks to patient safety and well-being, a key principle is that of proportionality. Risks associated with participation in the trial must be considered in the context of usual care. At present, there are no proven treatments for COVID-19 for clinicians in the community to prescribe safely with a sound evidence base.

Although data entry should be mindful of the desire to maintain integrity and audit trails, in the circumstances of this epidemic, the priority is on the timely entry of data that is sufficient to support reliable analysis and interpretation about treatment effects.

The Investigators will maintain appropriate medical and research records for this trial, in compliance with the requirements of the Medicines for Human Use (Clinical Trial) Regulations 2004, ICH E6 GCP and regulatory and institutional requirements for the protection of confidentiality of volunteers. The Chief Investigator, Principal Investigator, Co-Investigators, clinical team, including Clinical Research Nurses, and other authorised members of the trial team will have access to records. The Investigators will permit authorised representatives of the sponsor, and regulatory agencies to examine (and when required by applicable law, to copy) clinical records for the purposes of quality assurance reviews, audits and evaluation of the study safety and progress. The software used for the trial is described in *supplementary material E*.

#### 7 QUALITY ASSURANCE PROCEDURES

The study will be conducted in accordance with the current approved protocol, GCP, relevant regulations and PC-CTU Standard Operating Procedures. All PIs, coordinating centre staff and site staff will receive training in trial procedures according to GCP where required. Regular monitoring will be performed according to GCP using a risk-based approach. Data will be evaluated for compliance with the protocol and accuracy in relation to source documents where possible.

© Copyright: The University of Oxford and Oxford University Hospitals NHS Foundation Trust 2019

The PC-CTU Trial Management Group will be responsible for the monitoring of all aspects of the trial's conduct and progress and will ensure that the protocol is adhered to and that appropriate action is taken to safeguard participants and the quality of the trial itself. The TMG will be comprised of individuals responsible for the trial's day to day management and will meet regularly throughout the course of the trial.

#### 7.1 Risk assessment and Monitoring

A risk assessment and monitoring plan will be prepared before the study opens and will be reviewed as necessary over the course of the study to reflect significant changes to the protocol or outcomes of monitoring activities. Monitoring will be performed by the PC-CTU Quality Assurance Manager or delegate. The level of monitoring required will be informed by the risk assessment.

#### 7.2 Trial committees

The responsibilities of each group are as follows:

- Data Monitoring and Safety Committee (DMSC) - to review the data at each interim analysis as described in the Statistical Analysis section, as the updates to the randomisation scheme occur in order to ensure that the process is working correctly and to review and monitor the accruing data to ensure the rights, safety and wellbeing of the trial participants. Composition, and roles and responsibilities of the DMSC are detailed in the DMSC charter.
- Trial Steering Committee (TSC) - the Trial Steering Committee ensure the rights, safety and wellbeing of the trial participants. They will make recommendations about how the study is operating, any ethical or safety issues and any data being produced from other relevant studies that might impact the trial. Composition, and roles and responsibilities of the TSC are detailed in the TSC charter.
- Trial Management Group (TMG) - is responsible for the day-to-day running of the trial, including monitoring all aspects of the trial and ensuring that the protocol is being adhered to. It will include Co-Investigators and will meet weekly in the first instance. . Composition, and roles and responsibilities of the TMG are detailed in the TMG charter.
- A core project team (PT) from within the TMG will meet weekly or as required for operational decision making (met daily at the start of the trial).

### 8 PROTOCOL DEVIATIONS

A study related deviation is a departure from the ethically approved study protocol or other study document or process (e.g. consent process or administration of study intervention) or from Good Clinical Practice (GCP) or any applicable regulatory requirements. Any deviations from the protocol will be documented in a protocol deviation form and filed in the study master file.

A PC-CTU SOP is in place describing the procedure for identifying non-compliances, escalation to the central team and assessment of whether a non-compliance /deviation may be a potential Serious Breach.

### 9 SERIOUS BREACHES

© Copyright: The University of Oxford and Oxford University Hospitals NHS Foundation Trust 2019

A "serious breach" is a breach of the protocol or of the conditions or principles of Good Clinical Practice which is likely to affect to a significant degree –

(a) the safety or physical or mental integrity of the trial subjects; or
(b) the scientific value of the research.

In the event that a serious breach is suspected the Sponsor must be contacted within one working day. In collaboration with the CI, the serious breach will be reviewed by the Sponsor and, if appropriate, the Sponsor will report it to the approving REC committee and the relevant NHS host organisation within seven calendar days.

### 10 ETHICAL AND REGULATORY CONSIDERATIONS

#### 10.1 Declaration of Helsinki

The Investigator will ensure that this study is conducted in accordance with the principles of the Declaration of Helsinki.

#### 10.2 Guidelines for Good Clinical Practice

The Investigator will ensure that this trial is conducted in accordance with relevant regulations and with Good Clinical Practice.

#### 10.3 Approvals

Following Sponsor approval, the protocol, informed consent form, participant information sheets and any proposed informing material will be submitted to an appropriate Research Ethics Committee (REC), regulatory authorities, and host institution(s) for written approval. The PI and coordinating centres for each country will ensure and confirm correct regulatory approvals are gained prior to recruitment.

The Investigator will submit and, where necessary, obtain approval from the above parties for all substantial amendments to the original approved documents.

#### 10.4 Other Ethical Considerations

If a particular arm is deemed futile and dropped, no further participants will be randomised to this arm and anyone who is currently on this arm will be informed it has been dropped.
Once a particular intervention has been declared superior and effective, that will become the comparator arm (i.e. standard care).

Some participant's, due to their co-morbidities, will be exempt from prescription charges. Medication is most often sent from PC-CTU and so prescription charges will not apply. All participants will receive a £20 voucher to cover any prescriptions and other expenses they may incur as a consequence of study participation.

Participants who lack capacity to consent for themselves will only be recruited after consultation with their legal representative. Any sign of dissent in any form from the participant who lacks consent will be taken as an indication they do not wish to be involved and they will be withdrawn.

© Copyright: The University of Oxford and Oxford University Hospitals NHS Foundation Trust 2019

Only residents of care homes who lack capacity to consent will be recruited, adults who lack capacity to consent will not be recruited from the wider community.

##### 10.5 Reporting

The CI shall submit once a year throughout the clinical trial, or on request, an Annual Progress Report to the REC, HRA (where required), host organisation, funder (where required) and Sponsor. In addition, an End of Trial notification and final report will be submitted to the MHRA, the REC, host organisation and Sponsor.

##### 10.6 Transparency in Research

Prior to the recruitment of the first participant, the trial will have been registered on a publicly accessible database. Results will be uploaded to the European Clinical Trial (EudraCT) Database within 12 months of the end of trial declaration by the CI or their delegate. Where the trial has been registered on multiple public platforms, the trial information will be kept up to date during the trial, and the CI or their delegate will upload results to all those public registries within 12 months of the end of the trial declaration.

##### 10.7 Participant Confidentiality

The study will comply with the General Data Protection Regulation (GDPR) and Data Protection Act 2018, which require data to be de-identified as soon as it is practical to do so. The processing of the personal data of participants will be minimised by making use of a unique participant study number only on all study documents and any electronic database(s). All documents will be stored securely and only accessible by study staff and authorised personnel. The study staff will safeguard the privacy of participants' personal data.

##### 10.8 Expenses and Benefits

All participants will be reimbursed with a £20 voucher, to cover the payment of a prescription, should they incur this as a result of study participation, and a token of recognition of giving their time and contribution to the study. A proportion of people with the co-morbidities outlined and in the over 50 age-range , are not required to pay for prescriptions. Furthermore, medication is most often sent from the PC-CTU, and so prescription charges will not apply. Participants who complete a telephone interview as part of the qualitative sub-study will be reimbursed with a (second) £20 voucher for their time to participate.

#### 11 FINANCE AND INSURANCE

##### 11.1 Funding

The study is funded by the UKRI/NIHR via an MRC call.
The Department of Health & Social Care have provided the following drugs free of charge for trial Use: Hydroxychloroquine, Favipiravir.

##### 11.2 Insurance

The University has a specialist insurance policy in place, which would operate in the event of any participant suffering harm as a result of their involvement in the research (Newline Underwriting

© Copyright: The University of Oxford and Oxford University Hospitals NHS Foundation Trust 2019

Management Ltd, at Lloyd's of London). NHS indemnity operates in respect of the clinical treatment that is provided.

##### 11.3 Contractual arrangements

Appropriate contractual arrangements will be put in place with all third parties.

#### 12 PUBLICATION POLICY

The Investigators (those listed on the protocol and others to be decided at publication) will be involved in reviewing drafts of the manuscripts, abstracts, press releases and any other publications arising from the study. Authors will acknowledge the study funders. Authorship will be determined in accordance with the ICMJE guidelines and other contributors will be acknowledged.

#### 13 DEVELOPMENT OF A NEW PRODUCT/ PROCESS OR THE GENERATION OF INTELLECTUAL PROPERTY

Ownership of IP generated by employees of the University vests in the University. The University will ensure appropriate arrangements are in place as regards any new IP arising from the trial.

#### 14 ARCHIVING

Archiving will be done according to the UOXF PC-CTU SOP and study specific working instructions.

© Copyright: The University of Oxford and Oxford University Hospitals NHS Foundation Trust 2019

© Copyright: The University of Oxford and Oxford University Hospitals NHS Foundation Trust 2019

21. Chen J, Liu D, Liu L, Liu P, Xu Q, Xia L, et al. [A pilot study of hydroxychloroquine in treatment of patients with moderate COVID-19]. Zhejiang Da Xue Xue Bao Yi Xue Ban. 2020;49(2):215-9.
22. Gautret P, Lagier JC, Parola P, Hoang VT, Meddeb L, Mailhe M, et al. Hydroxychloroquine and azithromycin as a treatment of COVID-19: results of an open-label non-randomized clinical trial. Int J Antimicrob Agents. 2020:105949.
23. Retallack H, Di Lullo E, Arias C, Knopp KA, Laurie MT, Sandoval-Espinosa C, et al. Zika virus cell tropism in the developing human brain and inhibition by azithromycin. Proc Natl Acad Sci U S A. 2016;113(50):14408-13.
24. E Bosseboeuf MA, T Nhan, JJ de Pina, JM Rolain, D Raoult, et al. **Azithromycin inhibits the replication of Zika virus.** J Antivirals Antiretrovirals. 2018;10:6-11.
25. Madrid PB, Panchal RG, Warren TK, Shurtleff AC, Endsley AN, Green CE, et al. Evaluation of Ebola Virus Inhibitors for Drug Repurposing. ACS Infect Dis. 2015;1(7):317-26.
26. Bacharier LB, Guilbert TW, Mauger DT, Boehmer S, Beigelman A, Fitzpatrick AM, et al. Early Administration of Azithromycin and Prevention of Severe Lower Respiratory Tract Illnesses in Preschool Children With a History of Such Illnesses: A Randomized Clinical Trial. JAMA. 2015;314(19):2034-44.
27. Villarino AV, Kanno Y, O'Shea JJ. Mechanisms and consequences of Jak-STAT signaling in the immune system. Nat Immunol. 2017;18(4):374-84.
28. Hodge S, Hodge G, Holmes M, Jersmann H, Reynolds PN. Increased CD8 T-cell granzyme B in COPD is suppressed by treatment with low-dose azithromycin. Respirology. 2015;20(1):95-100.
29. Hodge S, Hodge G, Jersmann H, Matthews G, Ahern J, Holmes M, et al. Azithromycin improves macrophage phagocytic function and expression of mannose receptor in chronic obstructive pulmonary disease. Am J Respir Crit Care Med. 2008;178(2):139-48.
30. Hodge S, Reynolds PN. Low-dose azithromycin improves phagocytosis of bacteria by both alveolar and monocyte-derived macrophages in chronic obstructive pulmonary disease subjects. Respirology. 2012;17(5):802-7.
31. Baines KJ, Wright TK, Gibson PG, Powell H, Hansbro PM, Simpson JL. Azithromycin treatment modifies airway and blood gene expression networks in neutrophilic COPD. ERJ Open Res. 2018;4(4).
32. Mandell LA, Wunderink RG, Anzueto A, Bartlett JG, Campbell GD, Dean NC, et al. Infectious Diseases Society of America/American Thoracic Society consensus guidelines on the management of community-acquired pneumonia in adults. Clin Infect Dis. 2007;44 Suppl 2:S27-72.
33. Kawai S, Ochi M, Nakagawa T, Goto H. Antimicrobial therapy in community-acquired pneumonia among emergency patients in a university hospital in Japan. J Infect Chemother. 2004;10(6):352-8.
34. Gleason PP, Meehan TP, Fine JM, Galusha DH, Fine MJ. Associations between initial antimicrobial therapy and medical outcomes for hospitalized elderly patients with pneumonia. Arch Intern Med. 1999;159(21):2562-72.
35. Wu C, Liu Y, Yang Y, Zhang P, Zhong W, Wang Y, et al. Analysis of therapeutic targets for SARS-CoV-2 and discovery of potential drugs by computational methods. Acta Pharm Sin B. 2020.
36. Kim HS, Luo L, Pflugfelder SC, Li DQ. Doxycycline inhibits TGF-beta1-induced MMP-9 via Smad and MAPK pathways in human corneal epithelial cells. Invest Ophthalmol Vis Sci. 2005;46(3):840-8.
37. Clemens DL, Duryee MJ, Sarmiento C, Chiou A, McGowan JD, Hunter CD, et al. Novel Antioxidant Properties of Doxycycline. Int J Mol Sci. 2018;19(12).
38. Hsu AT, Barrett CD, DeBusk GM, Ellson CD, Gautam S, Talmor DS, et al. Kinetics and Role of Plasma Matrix Metalloproteinase-9 Expression in Acute Lung Injury and the Acute Respiratory Distress Syndrome. Shock. 2015;44(2):128-36.
39. NICE. COVID-19 rapid guideline: managing suspected or confirmed pneumonia in adults in the community. NICE; 2020.
40. Nicolau DV, Bafadhel M. Inhaled corticosteroids in virus pandemics: a treatment for COVID-19? Lancet Respir Med. 2020.
41. BTS/SIGN. BTS/SIGN British Guideline on the Management of Asthma. British Thoracic Society; 2019.
42. GINA. Global Strategy for Asthma Management and Prevention (2019 update). 2019.

© Copyright: The University of Oxford and Oxford University Hospitals NHS Foundation Trust 2019

43. Bloom CI, Elkin SL, Quint JK. Changes in COPD inhaler prescriptions in the United Kingdom, 2000 to 2016. Int J Chron Obstruct Pulmon Dis. 2019;14:279-87.
44. Artigas A, Camprubi-Rimblas M, Tantinya N, Bringue J, Guillamat-Prats R, Matthay MA. Inhalation therapies in acute respiratory distress syndrome. Ann Transl Med. 2017;5(14):293.
45. Ortiz-Diaz E, Li G, Kor D, Gajic O, Akca O, Adesanya A, et al. Preadmission Use of Inhaled Corticosteroids Is Associated With a Reduced Risk of Direct Acute Lung Injury/Acute Respiratory Distress Syndrome. CHEST. 2011;140(4):912A.
46. Festic E, Carr GE, Cartin-Ceba R, Hinds RF, Banner-Goodspeed V, Bansal V, et al. Randomized Clinical Trial of a Combination of an Inhaled Corticosteroid and Beta Agonist in Patients at Risk of Developing the Acute Respiratory Distress Syndrome. Crit Care Med. 2017;45(5):798-805.
47. Yamaya M, Nishimura H, Deng X, Sugawara M, Watanabe O, Nomura K, et al. Inhibitory effects of glycopyrronium, formoterol, and budesonide on coronavirus HCoV-229E replication and cytokine production by primary cultures of human nasal and tracheal epithelial cells. Respir Investig. 2020;58(3):155-68.
48. Merad M, Martin JC. Pathological inflammation in patients with COVID-19: a key role for monocytes and macrophages. Nature Reviews Immunology. 2020;20(6):355-62.
49. Barnes PJ. Inhaled Corticosteroids. Pharmaceuticals (Basel). 2010;3(3):514-40.
50. Peters MC, Sajuthi S, Deford P, Christenson S, Rios CL, Montgomery MT, et al. COVID-19 Related Genes in Sputum Cells in Asthma: Relationship to Demographic Features and Corticosteroids. Am J Respir Crit Care Med. 2020.
51. Inhaled corticosteroids downregulate the SARS-CoV-2 receptor ACE2 in COPD through suppression of type I interferon - preliminary report. 13 June 2020 (pre-print).
52. Lan J, Ge J, Yu J, Shan S, Zhou H, Fan S, et al. Structure of the SARS-CoV-2 spike receptor-binding domain bound to the ACE2 receptor. Nature. 2020;581(7807):215-20.
53. Hoffmann M, Kleine-Weber H, Schroeder S, Krüger N, Herrler T, Erichsen S, et al. SARS-CoV-2 Cell Entry Depends on ACE2 and TMPRSS2 and Is Blocked by a Clinically Proven Protease Inhibitor. Cell. 2020;181(2):271-80.e8.
54. Matsuyama S, Kawase M, Nao N, Shirato K, Ujike M, Kamitani W, et al. The inhaled corticosteroid ciclesonide blocks coronavirus RNA replication by targeting viral NSP15. bioRxiv. 2020:2020.03.11.987016.
55. Yamaya M, Nishimura H, Deng X, Sugawara M, Watanabe O, Nomura K, et al. Inhibitory effects of glycopyrronium, formoterol, and budesonide on coronavirus HCoV-229E replication and cytokine production by primary cultures of human nasal and tracheal epithelial cells. Respir Investig. 2020.
56. Reyes AZ, Hu KA, Teperman J, Wampler Muskardin TL, Tardif J-C, Shah B, et al. Anti-inflammatory therapy for COVID-19 infection: the case for colchicine. Annals of the Rheumatic Diseases. 2020:annrheumdis-2020-219174.
57. Rodriguez-Nava G, Trelles-Garcia DP, Yanez-Bello MA, Chung CW, Trelles-Garcia VP, Friedman HJ. Atorvastatin associated with decreased hazard for death in COVID-19 patients admitted to an ICU: a retrospective cohort study. Crit Care. 2020;24(1):429.
58. Deftereos SG, Giannopoulos G, Vrachatis DA, Siasos GD, Giotaki SG, Gargalianos P, et al. Effect of Colchicine vs Standard Care on Cardiac and Inflammatory Biomarkers and Clinical Outcomes in Patients Hospitalized With Coronavirus Disease 2019: The GRECCO-19 Randomized Clinical Trial. JAMA Netw Open. 2020;3(6):e2013136.
59. Scarsi M, Piantoni S, Colombo E, Airo P, Richini D, Miclini M, et al. Association between treatment with colchicine and improved survival in a single-centre cohort of adult hospitalised patients with COVID-19 pneumonia and acute respiratory distress syndrome. Ann Rheum Dis. 2020;79(10):1286-9.
60. Lopes MIea. Beneficial effects of colchicine for moderate to severe COVID-19: an interim analysis of a randomized, double-blinded, placebo controlled clinical trial. medRxiv preprint. 2020.
61. Chiu L, Chow R, Chiu N, Lo C-H, Aggarwal R, Lee J, et al. Colchicine use in patients with COVID-19: a systematic review and meta-analysis. medRxiv. 2021:2021.02.02.21250960.
62. Tardif J-C, Bouabdallaoui N, L'Allier PL, Gaudet D, Shah B, Pillinger MH, et al. Efficacy of Colchicine in Non-Hospitalized Patients with COVID-19. medRxiv. 2021:2021.01.26.21250494.

© Copyright: The University of Oxford and Oxford University Hospitals NHS Foundation Trust 2019

63. Piantoni S, Colombo E, Airò P, Tincani A, Brucato A, Franceschini F, et al. The rationale for the use of colchicine in COVID-19: comments on the letter by Cumhur Cure M et al. Clin Rheumatol. 2020;39(8):2489-90.
64. Piantoni S, Colombo E, Furloni R, Andreoli L, Brucato A, Imazio M, et al. Anti-inflammatory action of colchicine in hospitalised patients with COVID-19. Response to: 'Colchicine treatment in community healthcare setting to prevent severe COVID-19' by Della-Torre <em>et al</em>. Annals of the Rheumatic Diseases. 2020:annrheumdis-2020-218806.
65. Piantoni S, Patroni A, Toniati P, Furloni R, Franceschini F, Andreoli L, et al. Why not to use colchicine in COVID-19? An oldanti-inflammatory drug for a novel auto-inflammatory disease. Rheumatology. 2020;59(7):1769-70.
66. Prieto-Pena D, Dasgupta B. Biologic agents and small-molecule inhibitors in systemic autoimmune conditions: an update. Pol Arch Intern Med. 2020.
67. Dalbeth N, Lauterio TJ, Wolfe HR. Mechanism of action of colchicine in the treatment of gout. Clin Ther. 2014;36(10):1465-79.
68. Rodrigues TS, de Sá KSG, Ishimoto AY, Becerra A, Oliveira S, Almeida L, et al. Inflammasomes are activated in response to SARS-CoV-2 infection and are associated with COVID-19 severity in patients. J Exp Med. 2021;218(3).
69. Stewart S, Yang KCK, Atkins K, Dalbeth N, Robinson PC. Adverse events during oral colchicine use: a systematic review and meta-analysis of randomised controlled trials. Arthritis Research & Therapy. 2020;22(1):28.
70. Levy M, Spino M, Read SE. Colchicine: a state-of-the-art review. Pharmacotherapy. 1991;11(3):196-211.
71. Chen C, Zhang Y, Huang J, Yin P, Cheng Z, Wu J, et al. Favipiravir versus Arbidol for COVID-19: A Randomized Clinical Trial. medRxiv. 2020:2020.03.17.20037432.
72. Udwadia ZF, Singh P, Barkate H, Patil S, Rangwala S, Pendse A, et al. Efficacy and Safety of Favipiravir, an Oral RNA-Dependent RNA Polymerase Inhibitor, in Mild-to-Moderate COVID-19: A Randomized, Comparative, Open-Label, Multicenter, Phase 3 Clinical Trial. Int J Infect Dis. 2020.
73. Ivashchenko AA, Dmitriev KA, Vostokova NV, Azarova VN, Blinow AA, Egorova AN, et al. AVIFAVIR for Treatment of Patients With Moderate Coronavirus Disease 2019 (COVID-19): Interim Results of a Phase II/III Multicenter Randomized Clinical Trial. Clinical Infectious Diseases. 2020.
74. Khamis F, Al Naabi H, Al Lawati A, Ambusaidi Z, Al Sharji M, Al Barwani U, et al. Randomized controlled open label trial on the use of favipiravir combined with inhaled interferon beta-1b in hospitalized patients with moderate to severe COVID-19 pneumonia. International Journal of Infectious Diseases. 2021;102:538-43.
75. Cai Q, Yang M, Liu D, Chen J, Shu D, Xia J, et al. Experimental Treatment with Favipiravir for COVID-19: An Open-Label Control Study. Engineering. 2020;6(10):1192-8.
76. Pilkington V, Pepperrell T, Hill A. A review of the safety of favipiravir – a potential treatment in the COVID-19 pandemic? Journal of Virus Eradication. 2020;6(2):45-51.
77. Sanders JM, Monogue ML, Jodlowski TZ, Cutrell JB. Pharmacologic Treatments for Coronavirus Disease 2019 (COVID-19): A Review. JAMA. 2020;323(18):1824-36.
78. Coomes EA, Haghbayan H. Favipiravir, an antiviral for COVID-19? Journal of Antimicrobial Chemotherapy. 2020;75(7):2013-4.
79. Arshad U, Pertinez H, Box H, Tatham L, Rajoli RK, Curley P, et al. Prioritisation of potential anti-SARS-CoV-2 drug repurposing opportunities based on ability to achieve adequate plasma and target site concentrations derived from their established human pharmacokinetics. medRxiv. 2020:2020.04.16.20068379.
80. Kaptein SJF, Jacobs S, Langendries L, Seldeslachts L, ter Horst S, Liesenborghs L, et al. Favipiravir at high doses has potent antiviral activity in SARS-CoV-2−infected hamsters, whereas hydroxychloroquine lacks activity. Proceedings of the National Academy of Sciences. 2020;117(43):26955-65.
81. Driouich J-S, Cochin M, Lingas G, Moureau G, Touret F, Petit PR, et al. Favipiravir antiviral efficacy against SARS-CoV-2 in a hamster model. bioRxiv. 2020:2020.07.07.191775.

© Copyright: The University of Oxford and Oxford University Hospitals NHS Foundation Trust 2019

82. Cevik M, Tate M, Lloyd O, Maraolo AE, Schafers J, Ho A. SARS-CoV-2, SARS-CoV, and MERS-CoV viral load dynamics, duration of viral shedding, and infectiousness: a systematic review and meta-analysis. The Lancet Microbe.

83. CDC. https://www.cdc.gov.tw/File/Get/ht8jUiB_MI-aKnlwstwzvw.

84. Sissoko D, Laouenan C, Folkesson E, M'Lebing AB, Beavogui AH, Baize S, et al. Experimental Treatment with Favipiravir for Ebola Virus Disease (the JIKI Trial): A Historically Controlled, Single-Arm Proof-of-Concept Trial in Guinea. PLoS Med. 2016;13(3):e1001967.

© Copyright: The University of Oxford and Oxford University Hospitals NHS Foundation Trust 2019

#### 16 APPENDIX A: SCHEDULE OF PROCEDURES

| **Procedures** | **Participant contacts** | | | | | | | |
|---|---|---|---|---|---|---|---|---|
| | **Visit timing Day 0** | **Day 0** | **Day 0** | **Day 0** | **Daily Day 1-28 incl** | **Day 28-12 months (monthly contact)** | **Day 29-12mths** | **Up to 10 years** |
| | **Screening completed by participant online/phone** | **Eligibility completed by participant online/phone** | **Baseline completed by participant online/phone** | **Eligibility completed by Clinician online/phone** | **Symptom Diaries completed by participant online/phone** | **Contacted by study team if consent provided** | **Retrospective data collection by study team** | **By data extraction from clinical records** |
| Informed consent | X | X | X | X | X | | | |
| Demographics | X | X | X | | | | X | |
| Medical history | X | X | X | X | | | X | |
| Swab as part of the RCGP RSC/PHE national surveillance programme | When available, preferably by self-swabbing at study entry | | | | | | | |
| Concomitant medications | | X | | | | | X | |
| Eligibility assessment | X | X | | | | | | |

© Copyright: The University of Oxford and Oxford University Hospitals NHS Foundation Trust 2019

| | | | | | | | | |
|---|---|---|---|---|---|---|---|---|
| Randomisation | | | | X | | | | |
| Dispensing of trial drugs | | | | X | X | | | |
| Questionnaire | | | | | X | X | | |
| WHO 5 Well Being Index | X | | | | Day 14 and day 28 | X | | |
| Telephone interview (for subset of patient participants) | | | | | X | | | |
| Compliance | | | | | X | | | |
| Adverse event assessments | | | | | X* | | X | |
| Optional SARS-CoV-2 blood test as part of the RCGP RSC/PHE national surveillance programme | | | | | | | X | |
| Evidence of sequalae and health care utilisation | | | | | | X | | X |

© Copyright: The University of Oxford and Oxford University Hospitals NHS Foundation Trust 2019

* Patient reported AEs will be assessed by a clinician for certain IMPs, as specified in the Intervention Specific Appendices. These treatments include HCQ (no longer an active treatment arm) and drugs that are not licensed for use in the UK.

© Copyright: The University of Oxford and Oxford University Hospitals NHS Foundation Trust 2019

#### 17 APPENDIX B: AMENDMENT HISTORY

| Amendment No. | Protocol Version No. | Date issued | Author(s) of changes | Details of Changes made |
|---|---|---|---|---|
| 1 (SA1) | 1.1 | | Emma Ogburn; Chris Butler; Gail Hayward | Inclusion criteria: change 'known heart disease' to 'Known heart disease and/or hypertension'; Exclusion criteria: exclude patients taking the following drugs: penicillamine, amiodarone, ciclosporin, chloroquine. Update section 9.6 to include vision changes and lowering of blood sugar. Update change in Funder and update Investigator list to reflect UKRI funder bid. |
| 2 (SA2) | 2.0 | | Emma Ogburn; Chris Butler; Gail Hayward, Hannah Swayze | Inclusion of TSC; central facility to distribute patient packs; addition of third arm; update of secondary outcomes to include WHO wellbeing questions; qualitative sub study; sign posting to other RCGP RSC study; eligibility confirmation by research nurse. |
| 3 (SA3) | 2.1 | | Hannah Swayze; Chris Butler; Emma Ogburn; Gail Hayward | Trial rationale; secondary outcomes to include blood test; 14 days of covid-19 symptoms; call to participant at day 2; poster |
| *4 (SA4)* | *2.1* | | *No changes to the protocol* | |
| 5 (SA5) | 3.0 | | Hannah Swayze; Chris Butler; Emma Ogburn; Gail Hayward | Updated Azithromycin information; broadening of inclusion criteria; first interim analysis; primary analysis details; care home materials; administrative and typographical updates; study partner letter; recruitment via social media, care homes and pharmacies; GPs prescribe trial medication; eligibility to at least one intervention arm as well as the |

© Copyright: The University of Oxford and Oxford University Hospitals NHS Foundation Trust 2019

| | | | | |
|---|---|---|---|---|
| | | | | Usual Care arm; ICF may be sent to participants. |
| 6 (SA6) | 4.0 | | Chris Butler; Emma Ogburn; Gail Hayward; Ben Saville; Ly-Mee Yu; Hannah Swayze | Updating inclusion criteria; updating the rationale and evidence for safety of hydroxychloroquine; inclusion of a new arm, doxycycline; AE reporting for hydroxychloroquine arm; typographical clarifications. |
| 7 (NS1) | 4.0 | | *No changes to the protocol* | |
| 8 (SA7) | 5.0 | | Chris Butler; Emma Ogburn; Ben Saville; Ly-Mee Yu; Hannah Swayze | Including a second primary outcome, time to recovery, change to sample size estimation, new eligibility criteria: obesity, formatting changes, blood test process. |
| 9 (SA8) | 5.0 | | *No changes to the protocol* | |
| 10 (SA9) | 5.0 | | *No changes to the protocol* | |
| 11 (NS2) | 5.0 | | *No changes to the protocol* | |
| 12 (SA10) | 6.0 | | Chris Butler; Emma Ogburn; Hannah Swayze | Addition of inhaled corticosteroid treatment arm, enrolment to additional trials, long-term follow-up, access to NHS Digital Pillar 2 test data, removal of investigators, additional trial contact with participants for up to 12 months, changes to objectives/outcomes/ time-points, removal of sampling from study |
| 13 (NS3) | 6.1 | | Sharon Tonner | Removal of patient already taking a treatment arm medication as an exclusion |
| 14 (NS4) | 6.1 | | *No changes to the protocol* | |
| 15 (SA11) | 6.2 | | Sharon Tonner, Hannah Swayze | Inclusion of patients who lack capacity to consent, discontinuation of azithromycin arm |
| 16 (SA12) | 7.0 | | Chris Butler; Emma Ogburn; Hannah Swayze; | Addition of colchicine treatment arm. Data management proportional approach. Discontinuation of doxycycline |

© Copyright: The University of Oxford and Oxford University Hospitals NHS Foundation Trust 2019

| | | | | |
|---|---|---|---|---|
| | | | Emily Bongard; Julie Allen; Jienchi Dorward; Oliver Van Hecke | arm. Stylistic and typographical updates. Funding arrangements. Inclusion criteria - to include those aged 18 or over. Rephrasing secondary outcome: daily rating of how well participant feels. Statistical analysis. |
| 17 (SA13) | 7.1 | | Chris Butler; Emma Ogburn; Hannah Swayze; Emily Bongard; Ly-Mee Yu; Jienchi Dorward | Addition of safety monitoring procedures for drugs that are not licensed in the UK. Addition of safety as a secondary endpoint. Addition of favipiravir treatment arm. Stylistic and typographical updates. GP Contact for safety monitoring. In-kind contributions. Using JDR as a recruitment tool. The primary analysis population defined as those with a COVID-19 positive test. Drug accountability for favipiravir. |

Lists details of all protocol amendments whenever a new version of the protocol is produced.

Protocol amendments must be submitted to the Sponsor for approval prior to submission to the REC committee, HRA (where required) or MHRA.

© Copyright: The University of Oxford and Oxford University Hospitals NHS Foundation Trust 2019

### 18 APPENDIX C: USUAL CARE ARM

#### 1. Background and rationale

COVID-19 disproportionately affects people with comorbidities, more severe illness, and who are older. The disease causes considerable morbidity and mortality in this population group in particular, and is having a devastating effect on people's health, and society in the UK and internationally.(2, 3, 6, 8) So far, there are no specific treatments for COVID-19 that have been proven in rigorous clinical trials to be effective and that can be used in the community. Clinicians managing possible COVID-19 in the community will make clinical judgements about best treatment based on the clinical situation, but care is usually supportive to begin with, unless patients deteriorate and require hospital admission https://www.nice.org.uk/guidance/ng163). The National Institute for Health and Care Excellence does not recommend the immediate use of antibiotics unless there are signs of pneumonia (https://www.nice.org.uk/guidance/ng163).

This Usual Care arm will follow current NHS care provision, and provides a control against which the effect of new interventions that are added to usual care can be assessed. If a new trial intervention plus usual care is found to be superior to usual care alone, then the usual care alone arm will be dropped, and the intervention that is found to be most effective will become the standard of care within the trial.

#### 2. Changes to outcome measures

None

#### 3. Detail of intervention

Participants randomised to the usual care arm will receive usual clinical care as per NHS care delivery practice.

##### a. Investigational Medicinal Product (IMP) description

Not applicable

##### b. Storage of IMP

Not applicable

#### 4. Safety reporting

Mechanisms for safety reporting are outlined in the trial protocol.

© Copyright: The University of Oxford and Oxford University Hospitals NHS Foundation Trust 2019

### 19 APPENDIX D: USUAL CARE PLUS HYDROXYCHLOROQUINE ARM (DISCONTINUED)

#### 1. Background and rationale

##### a. Evidence for potential Hydroxychloroquine benefits in COVID-19

A candidate intervention for COVID-19, a drug called hydroxychloroquine, has become available following early evaluation in some studies in China.(9, 10) Hydroxychloroquine is a hydroxylated version of the drug chloroquine.(10, 11) Both agents are commonly in use as anti-malarials, and are used in a variety of auto-immune diseases. They have received significant recent interest as potential modifiers of disease activity in COVID-19. (10, 12, 13) Hydroxychloroquine is already available within the NHS on prescription for other indications, and has a generally benign safety profile.(14) Chloroquine is available to buy in the UK over the counter in some formulations and is used as antimalarial prophylaxis and treatment.

Chloroquine is known to block virus infection by increasing endosomal pH required for virus/cell fusion, as well as interfering with the glycosylation of cellular receptors of SARS-CoV-2.(5) Besides its antiviral activity, chloroquine has an immune-modulating activity, which may synergistically enhance its antiviral effect *in vivo*.(11) Chloroquine is widely distributed in the whole body, including lungs, after oral administration.(10) The $EC_{90}$ value of chloroquine against the 2019-nCoV in Vero E6 cells was 6.90 μM in one study (9) which can be clinically achievable as demonstrated in the plasma of rheumatoid arthritis patients who received 500 mg administration.(14)

Hydroxychloroquine has been found to be effective against intracellular micro-organisms including malaria and intracellular bacteria *Coxiella burnetii* and *Tropheryma Whipplei*.(11) Both chloroquine and hydroxychloroquine have been shown to have *in vitro* antiviral activity against SARS coronavirus in a number of studies.(11) Most recently activity against SARSCOV2 was shown to be greater for hydroxychloroquine than chloroquine (15).

**Key publications that have relevance to the safety and rationale for use of hydroxychloroquine in the PRINCIPLE Trial:**

**1. *The Mahévas study*** was an observational study that assessed whether hydroxychloroquine reduced the need for transfer to ICU in patients already sick enough to be hospitalised.(16) It focussed on sicker patients with hypoxic pneumonia, some requiring ITU care. It did not find a difference in transfers to ICU. So the question and population in the Mahevas study are very different compared to PRINCIPLE. Most importantly, unlike PRINCIPLE, the Mahevas study is ***not a randomised clinical tri***al. Numbers were relatively small (n=181), and it is at high risk of bias due to the observational design.

***Regarding safety***, those receiving hydroxychloroquine were prescribed 600mg per day, whereas the dose in the PRINCIPLE trial is 400mg per day; 18% of those who received hydroxychloroquine in the Mahévas study were also on azithromycin (which can be arrhythmogenic), and this combination is not possible in PRINCIPLE because of the additive risk. Moreover, PRINCIPLE excludes several other drug combinations that could be arrhythmogenic. In the Mahevas study, eight patients (10%) who were taking hydroxychloroquine experienced electrocardiographic changes that required discontinuation of hydroxychloroquine. Critically, those in the control

© Copyright: The University of Oxford and Oxford University Hospitals NHS Foundation Trust 2019

group did not have ECGs done, so we don't know if there was indeed a difference between groups, and we cannot therefore attribute the ECG changes to hydroxychloroquine. COVID-19 itself, or drug interactions, may well have been underlying reasons. The authors state, "Although hydroxychloroquine is considered safe in the context of systemic lupus erythematosus, these adverse events might be explained by the use of high dose hydroxychloroquine in patients older than 75 years with renal impairment and frequent drug interactions. We cannot rule out the possibility that these cardiac effects attributed to hydroxychloroquine were caused by COVID-19, especially given electrocardiograms were unavailable during follow-up in the control group."

**2. *The Tang study*** was a hospital-based, randomised study and included 150 patients; randomisation was done using sealed envelopes.(17) The trial found no difference in the proportion of patients with two sequential negative swab results.

***Regarding safety***,75 participants received hydroxychloroquine 1200 mg daily for 3 days and then 800 mg for either 2 or 3 weeks. Again, the dose used in this study was much higher that the dose being used in PRINCIPLE (initially three times, and subsequently twice as high as PRINCIPLE). However, 63% and 64% of patients in the hydroxychloroquine and control groups respectively also received other antiviral agents. In PRINCIPLE, we are not evaluating the combination of antiviral agents and hydroxychloroquine. Importantly, this study did not find evidence of cardiac arrhythmias associated with hydroxychloroquine use. The authors sate, "Events of cardiac arrhythmia, such as prolonged QT interval were not observed in our trial, possibly because of the relatively mild to moderate disease of patients investigated or the short term period of follow-up."

**3. *The Mehra study*** published in the Lancet on 22.05.2020 reported an association between hydroxychloroquine use and cardiac events and mortality amongst patients hospitalised with COVID-19.(18) The *observational* study design is inherently susceptible to bias, the study data integrity has been queried given the homogeneity of the baseline characteristics, the adequacy of the adjustment for confounders cannot be assessed from the published methods, and the registries used are in a different patient population compared to PRINCIPLE. Patients were much sicker and more advanced in the illness than in PRINCIPLE. The authors themselves state that "*Randomised clinical trials will be required before any conclusion can be reached regarding benefit or harm of these agents (hydroxychloroquine and chloroquine) in COVID-19 patients*." The authors also state "These data do not apply to the use of any treatment regimen used in the ambulatory, out-of-hospital setting." This study has proved hugely controversial on social media, with a number of methodological and data integrity concerns already raised, for example:

1. There were inadequate adjustments for known and measured confounders (disease severity, temporal effects, site effects, dose used).
2. The authors have not adhered to standard practices in the machine learning and statistics community. They have not released their code or data. There is no data/code sharing and availability statement in the paper. The Lancet was among the many signatories on the Wellcome statement on data sharing for COVID 19 studies.
3. There was no ethics review.
4. There was no mention of the countries or hospitals that contributed to the data source, no acknowledgments to their contributions. A request to the authors for information on the contributing centres was denied.
5. Data from Australia are not compatible with government reports (too many cases for

© Copyright: The University of Oxford and Oxford University Hospitals NHS Foundation Trust 2019

just five hospitals, more in-hospital deaths than had occurred in the entire country during the study period). Surgisphere (the data company) have since claimed this was an error of classification.

6. Data from Africa indicate over 40% of all COVID-19 cases and deaths in the continent occurred in *Surgisphere*-associated hospitals which had sophisticated electronic patient data recording, and patient monitoring able to detect and record “non-sustained [at least 6 secs] or sustained ventricular tachycardia or ventricular fibrillation”. This seems unlikely.
7. Unusually small reported variances in baseline variables, interventions and outcomes between continents
8. Mean daily doses of hydroxychloroquine that are 100 mg higher than FDA recommendations, whilst 66% of the data are from North American hospitals.
9. Implausible ratios of chloroquine to hydroxychloroquine use in some continents.
10. The tight 95% confidence intervals reported for the hazard ratios are unlikely. For instance, for the Australian data this would need about double the numbers of recorded deaths that were reported in the paper.

This paper has now been retracted, and the data cannot be verified.

**4.** ***The Geleris study*** was an observational study of 1,376 consecutive COVID-19 patients at a New York hospital to determine whether hydroxychloroquine use was associated with intubation or death, as a primary composite outcome.(19) 811 (58.9%) of these patients received hydroxychloroquine. The authors excluded patients who were intubated, died, or who were transferred to another facility within 24 hours after presentation to the emergency department from the analyses. A propensity score matching model (C-statistic of 0.81) was used to ensure that groups were similar at baseline.

***Regarding safety***, multivariable adjusted analyses with inverse probability weighting revealed no significant association between treatment with hydroxychloroquine and intubation or death (HR 1.04 (95% CI 0.82 – 1.32)). Whilst the patient population in this study is different to that of PRINCIPLE, it is interesting that the findings contrast with those of a recent Lancet study published by Mehra *et al*. One possible reason for the difference is that patients receiving interventions like hydroxychloroquine in the study by Mehra *et al* were sicker than those in the study’s control group. This may have arisen through use of crude measures to account for baseline disease severity (qSOFA score and SpO2 < 94%) in their propensity score matching model, and may also explain the big differences seen in patients requiring mechanical ventilation between controls (7.7%) and those in intervention groups (20-21.6%).

**5.** ***Boulware*** and colleagues conducted a Covid-19 postexposure prophylaxis, placebo controlled randomised trial of hydroxychloroquine in 821 asymptomatic patients; 11.8% of those taking hydroxychloroquine vs 14.3 of those taking placebo experienced a new illness compatible with COVID-19 (absolute difference -2.4%) but this difference was not statistically significant, indicating no evidence of benefit from the hydroxychloroquine. (20)

***Regarding safety***, while side effects were more common with hydroxychloroquine than with placebo (40.1% vs. 16.8%), no serious adverse reactions were reported.

© Copyright: The University of Oxford and Oxford University Hospitals NHS Foundation Trust 2019

**Earlier studies of hydroxychloroquine for COVID-19**

**1. *Chen*** and colleagues conducted a *randomised* controlled trial to test the effectiveness of hydroxychloroquine in 30 adult patients who tested positive for COVID-19 in China.(21) Patients in the treatment group received 400mg of hydroxychloroquine for 5 days, while the control group received usual care. The result of a nasopharyngeal swab on Day 7 was used as the primary outcome. The intention- to- treat analysis revealed that the treatment group did not differ from the control group in the number of patients testing negative for COVID-19 on Day 7 (13 versus 14 patients), nor the duration of illness (all P>0.05).

***Regarding safety*,** the authors report three adverse events in the control group (one patient with abnormal liver function and anaemia, and one patient with abnormal renal function), and four adverse events in the treatment group (two patients with diarrhoea, one with lethargy, and one patient with abnormal liver function tests), which the authors argue were not linked to treatment with HCQ. One patient in the treatment group deteriorated significantly and thus HCQ was stopped on Day 4 of the treatment. This study was under-powered according to their own calculations.

***2. Gautret*** and colleagues presented the results of an open- label, non-randomised trial with 36 patients diagnosed with COVID-19 in French hospitals.(22) Six participants were asymptomatic, 22 had upper respiratory tract infection symptoms, and eight had lower respiratory tract infection symptoms. The twenty patients in the treatment group received HCQ 200mg three times a day for 10 days. Patients declining to take part in the study and not meeting the inclusion criteria were assigned to the control group and received usual care. Six of the patients in the treatment group additionally received azithromycin to prevent bacterial superinfection. The primary outcome was SARS- CoV-2 carriage at Day 6 on nasopharyngeal swabs. Patients treated with hydroxychloroquine were significantly more likely to test negative for SARS- CoV-2 on Day 6 compared with controls (70% versus 12.5% virologically cured, p<0.001). All patients treated with hydroxychloroquine and azithromycin tested negative on Day 6.

***Regarding safety,*** the authors did not report any safety data, stating that this would follow in a subsequent publication. Aside from a lack of adverse event reporting, there are many problems with the study methodology including the non-randomized design, under-powered sample size, lack of intention-to-treat analysis, and absence of medium to long-term follow-up data.

***3. Chen*** and colleagues conducted a *randomised* clinical trial of adult patients admitted to hospital with confirmed COVID-19.(7) Sixty two patients were randomly assigned to usual care (n=31) or hydroxychloroquine (200 mg BD) for five days in addition to usual care (n=31). The authors report that there were 'significant differences' in time to clinical recovery (TTCR) between the two groups, with TTCR defined as the return of body temperature and cough relief, maintained for more than 72 hours. They also report that all four patients who 'progressed to severe disease' were in the control group. The reporting of empirical data by the authors is limited and unclear. They did not include a power calculation, but presumably this study was under-powered to detect differences between groups. No medium to long-term follow-up data is presented.

***Regarding safety,*** the authors report that two mild adverse events occurred (a rash and a headache), both of which were in patients receiving hydroxychloroquine. No patients receiving usual care experienced adverse events.

© Copyright: The University of Oxford and Oxford University Hospitals NHS Foundation Trust 2019

**In summary**

The large scale hospital based Recovery trial has recently announced that they found no benefit from hydroxychloroquine (as yet unpublished). No safety concerns have been reported by the Principle Trial. A post exposure prophylaxis study found no benefit from hydroxychloroquine, but also found no safety concerns. These studies address a different research question and focus on different patient populations in comparison to the Principle Trial. Evidence about early treatment of COPVID-19 in the community is urgently needed: the potential application of the findings of the PRINCIPLE Trial of community treatment is considerable, and the 'reach' of the study is now nation-wide. Our study population are patients in the community and our trial question is about early treatment. Outcome data from studies with sicker hospitalised patients may not apply to our study population

A key, controversial observational study (Mehra et al) reported that those taking hydroxychloroquine had worse outcomes and suffered more cardiac events than those not taking hydroxychloroquine. However, major doubts have been expressed about the data integrity of this study and insufficient detail in the paper to judge the adequacy of the methods employed to adjust for the inevitable confounders in an observational study. Hydroxychloroquine is not a licensed drug for treating COVID-19. Patients doing well are therefore less likely to be prescribed this drug. When a patient is causing their clinical team more concern or their condition is deteriorating, the chances of them being prescribed hydroxychloroquine will be greater. Adjustment for potential confounders has been inadequate in the observational studies. Critically, these studies cannot adjust for the clinician's sense of how the patient is faring over time. The Mehra study has been retracted and can't be relied upon.

The deficiencies and differences in all of these studies highlight the need for well-conducted, adequately powered randomised clinical trials, to provide definitive evidence of the safety and effectiveness of hydroxychloroquine for the early community treatment COVID-19 illness. PRINCIPLE will assess whether hydroxychloroquine is safe and effective if given earlier in the course of illness and in patients with milder symptoms not requiring hospital admission.

#### 2. Eligibility criteria specifically related to hydroxychloroquine

Inclusion criteria:

Exclusion criteria:

- Pregnancy;
- Breastfeeding;
- Known severe hepatic impairment;
- Known severe renal impairment;
- Known porphyria;
- Type 1 diabetes or insulin dependent Type 2 Diabetes mellitus ;
- Known G6PD deficiency;
- Known myasthenia gravis;
- Known severe psoriasis;
- Known severe neurological disorders (especially those with a history of epilepsy—may lower seizure threshold)
- Previous adverse reaction to, or currently taking, hydroxychloroquine or chloroquine

© Copyright: The University of Oxford and Oxford University Hospitals NHS Foundation Trust 2019

Patients currently taking the following drugs: penicillamine, amiodarone, ciclosporin, digoxin: the following antimicrobials; azithromycin, clarithromycin, erythromycin, ciprofloxacin, levofloxacin, moxifloxacin, ketoconazole, itraconazole, or mefloquine: the following antidepressants; amitriptyline, citalopram, desipramine, escitalopram, imipramine, doxepin, fluoxetine, wellbutrin, venlafaxine; the following antipsychotics or mood stabilizers; haloperidol, droperidol, lithium, quetiapine, thioridazine, ziprasidone: methadone: sumatriptan, zolmitriptan

- Known congenital or documented QT prolongation
- Known retinal disease

##### 3. Outcome measures related to hydroxychloroquine

There are no outcome measures related specifically to this usual care plus hydroxychloroquine arm

##### 4. Detail of intervention

Participants randomised to the usual care plus hydroxychloroquine arm will receive usual clinical care as per NHS guidelines, plus a course of oral hydroxychloroquine 200mg twice daily for seven days.

###### a. Investigational Medicinal Product (IMP) description

Hydroxychloroquine sulphate 200 milligram (mg) tablets. The tablets are for oral administration. One tablet (200mg) hydroxychloroquine to be taken twice daily for 7 days by mouth (14 tablets in total).
Special instructions: Each dose should be taken with a meal or glass of milk. Antacids may reduce absorption of hydroxychloroquine so it is advised that a 4-hour interval be observed between taking hydroxychloroquine and an antacid.
This is the standard therapeutic dose for its normal indication in adults which is for the treatment of rheumatoid arthritis, discoid and systemic lupus erythematosus, and dermatological conditions caused or aggravated by sunlight.

The Marketing Authorisation holder is Zentiva Pharma UK Limited Guildford Surrey GU1 4YS United Kingdom. Marketing authorisation number is PL 17780/0748.

###### b. Storage of IMP

: Stored at room temperature in locked cupboards in restricted access rooms in the Nuffield Department of Primary Care Health Sciences; in locked cupboards in restricted access rooms in GP Practices; in Pharmacies.

For hydroxychloroquine, GP practices can order a supply of trial medication from Public Health England using the existing ImmForm process. GPs will be provided with an envelope by the trial team which will be labelled appropriately for trial medication, and they will add the patient's details to this label. This pack, containing instructions on using the medication will be provided to the patient or their representative.

© Copyright: The University of Oxford and Oxford University Hospitals NHS Foundation Trust 2019

##### c. SmPC precautions and concomitant medication

Hydroxychloroquine: Hydroxychloroquine will be used for short-term use (7 days) in this trial. The SmPC and precautions listed below focus on longer term chronic use.

###### *i.* Precautions

Hydroxychloroquine might lower blood sugar levels in some people. If participants develop these symptoms, they will be advised in the Patient Information documents to eat something sweet and seek clinical advice if the symptoms continue.

Hydroxychloroquine occasionally causes blurred vision, which typically resolves once the medication is stopped. Participants will be advised via the Participant Information documents that if they develop any problems with vision, they should stop taking the medication immediately, seek clinical advice, and not drive or operate any heavy machinery.

###### ii. Concomitant medication

Hydroxychloroquine sulfate has been reported to increase plasma digoxin levels. Serum digoxin levels should be closely monitored in participants receiving concomitant treatment.

Hydroxychloroquine sulfate may also be subject to several of the known interactions of chloroquine even though specific reports have not appeared. These include: potentiation of its direct blocking action at the neuromuscular junction by aminoglycoside antibiotics; inhibition of its metabolism by cimetidine which may increase plasma concentration of the antimalarial; antagonism of effect of neostigmine and pyridostigmine; reduction of the antibody response to primary immunisation with intradermal human diploid-cell rabies vaccine.

As with chloroquine, antacids may reduce absorption of hydroxychloroquine so it is advised that a four hour interval be observed between hydroxychloroquine and antacid dosaging.

As hydroxychloroquine may enhance the effects of a hypoglycaemic treatment, a decrease in doses of insulin or antidiabetic drugs may be required.

Halofantrine prolongs the QT interval and should not be administered with other drugs that have the potential to induce cardiac arrhythmias, including hydroxychloroquine. Also, there may be an increased risk of inducing ventricular arrhythmias if hydroxychloroquine is used concomitantly with other arrhythmogenic drugs, such as amiodarone and moxifloxacin.

An increased plasma ciclosporin level was reported when ciclosporin and hydroxychloroquine were co-administered.

Hydroxychloroquine can lower the convulsive threshold. Co-administration of hydroxychloroquine with other antimalarials known to lower the convulsion threshold (e.g. mefloquine) may increase the risk of convulsions. Also, the activity of anti-epileptic drugs might be impaired if co-administered with hydroxychloroquine. In a single-dose interaction study, chloroquine has been reported to reduce the bioavailability of praziquantel. It is not known if there is a similar effect when hydroxychloroquine and praziquantel are co-administered. Per extrapolation, due to the similarities in structure and pharmacokinetic parameters between hydroxychloroquine and chloroquine, a similar effect may be expected for hydroxychloroquine.

© Copyright: The University of Oxford and Oxford University Hospitals NHS Foundation Trust 2019

There is a theoretical risk of inhibition of intra-cellular α-galactosidase activity when hydroxychloroquine is co-administered with agalsidase.

##### iii. Pregnancy and Breastfeeding

A moderate amount of data on pregnant women (between 300 – 1000 pregnancy outcomes), including prospective studies in long-term use with large exposure, have not observed a significant increased risk of congenital malformations or poor pregnancy outcomes. Hydroxychloroquine crosses the placenta. Only limited non-clinical data are available for hydroxychloroquine, data on chloroquine have shown developmental toxicity at high supratherapeutic doses and a potential risk of genotoxicity in some test systems. Therefore, hydroxychloroquine sulfate should be avoided in pregnancy except when, in the judgement of the physician, the individual potential benefits outweigh the potential hazards. Careful consideration should be given to using hydroxychloroquine during lactation, since it has been shown to be excreted in small amounts in human breast milk, and it is known that infants are extremely sensitive to the toxic effects of 4-aminoquinolines.
Pregnancy and breastfeeding are exclusion criteria for the hydroxychloroquine arm of the PRINCIPLE trial.

#### 5. Safety reporting

Hydroxychloroquine: has a well-documented safety profile and is a commonly used medication in a primary care setting (see above).

Common symptoms of hydroxychloroquine include abdominal pain; appetite decreased; diarrhoea; emotional lability; headache; nausea; skin reactions; vision disorders; and vomiting.
Mechanisms for safety reporting are outlined in the trial protocol.

We will call all participants randomised to hydroxychloroquine on day 7 to ask about cardiovascular AEs. Our team of clinicians will review any AEs relating to cardiovascular symptoms from the day 7 call, and assess whether these may be related to hydroxychloroquine. If AEs are thought to be related and it's deemed necessary by the assessing clinician, the participant's GP will be contacted to arrange a face-to-face visit for further clinical evaluation.

© Copyright: The University of Oxford and Oxford University Hospitals NHS Foundation Trust 2019

### 20 APPENDIX E: USUAL CARE PLUS AZITHROMYCIN ARM (DISCONTINUED)

#### 1. Background and rationale

##### a. Evidence for potential Azithromycin benefits in COVID-19

Atypical macrolides, especially Azithromycin, have activities that may be beneficial in the treatment of COVID-19 patients, and especially those in the at-risk or age range of the PRINCIPLE trial.

Firstly, Azithromycin appears to have some anti-viral mechanisms. In COVID-19, Azithromycin appears to inhibit viral replication and therefore reduces shedding. In the small open observational trial of Gautret *et al* the addition of azithromycin to hydroxychloroquine (HCQ) (at 200 tds for 10 days) in 6 of the 14 HCQ subjects of the total 36 COVID-19 patients in the study significantly reduced viral shedding at 3 days to 15% (one subject) versus 70% in the HCQ arm and 95% in the indirect control arm, with no shedding at 6 days in the combination arm versus 50% and 90% respectively.(22) Azithromycin was also used in some Chinese observational and interventional studies.

Azithromycin has also been shown to be active ***in vitro*** against Zika and Ebola viruses,(23-25) and to prevent severe respiratory tract infections when administrated to patients suffering viral infection.(26) Inhibition of viral infections by azithromycin may be linked to its suppressive effect on the production of viral interferon.(27) Longer term administration of low dose azithromycin in COPD has been shown to suppress proinflammatory cytokine production, potentiate macrophage phagocytosis and anti-inflammatory cytokine expression.(28-30) Azithromycin use is also associated with a decrease in the expression of human HLA (human leukocyte antigen) complex molecules in the respiratory tract, including HLA-A, HLA-B, HLA-DPA1, HLA-DRA, HLA-DRB4.(31)

##### b. Importance of treating CAP or CAP risk in the elderly or immuno-compromised

An important secondary pathway to severe illness and death with COVID-19 may be secondary infection and sepsis in the immune-compromised state, especially secondary community or hospital acquired pneumonia. Older people are more susceptible to pneumonia because of comorbidities, a weakened immune system and are therefore more likely to die.(32) The onset of pneumonia in the elderly can often be rapid, and for severe pneumonia, the prognosis is poor: as many as one in five will die.(32) Severe pneumonia is more prevalent the older you are and in those with more serious underlying diseases.(33) The leading cause of death is respiratory insufficiency. Death has been shown to increase in those not responding to initial antimicrobials, and consequently, the initial selection of the agent is important.
Common causative organisms in the elderly admitted to the hospital with pneumonia include *Haemophilus influenza, Staphylococcus aureus, Streptococcus pneumoniae,* and *Mycoplasma pneumoniae*. In severe pneumonia, *S. aureus, Klebsiella pneumoniae*, and *Pseudomonas aeruginosa* have been identified as common causative organisms. Older patients often have polymicrobial infections, which may be a factor in non-responders. Assessment of 12,945 US

© Copyright: The University of Oxford and Oxford University Hospitals NHS Foundation Trust 2019

Medicare inpatients over 65 with pneumonia found that initial treatment with a second-generation cephalosporin plus macrolide ([HR, 0.71; 95% CI, 0.52-0.96), a non-pseudomonas third-generation cephalosporin plus a macrolide (HR, 0.74; 0.60-0.92), or a fluoroquinolone alone (HR, 0.64; 0.43-0.94) was associated with lower 30-day mortality.(34)

For CAP management NICE guidance currently recommends Amoxycillin 500mg tds combined with Clarithromycin 500mg bd for 5 days or, in penicillin sensitive, Clarithromycin 500mg bd for 5 days or Doxycycline 200mg stat then 100mg daily for the next 4 days. They also recommend starting therapy within 4 hours. The identification of the early stages of pneumonia in older patients can prove challenging since traditional symptoms and signs, including fever, may be lacking.

Azithromycin will have at least as broad a spectrum of action as clarithromycin in terms of bacterial infections and the additional potential anti-viral activity which has not been observed for other macrolides like Clarithromycin. It will also cover atypical organisms.

#### 2 Changes to outcome measures

The addition of this usual care plus azithromycin arm will not require any changes to outcome measures

#### 3 Eligibility criteria specifically related to azithromycin

Inclusion criteria: No changes

Exclusion criteria:

- Pregnancy
- Breastfeeding
- Known severe hepatic impairment;
- Known severe renal impairment;
- Known myasthenia gravis;
- Previous adverse reaction to, or currently taking, azithromycin or other macrolides or ketolides
- Patients taking the following drugs: hydroxychloroquine or chloroquine, sotalol, amiodarone, ciclosporin, digoxin, bromocriptine, cabergoline, ergotamine, ergometrine, methysergide or any ergot derivatives.
- Already taking antibiotics for an acute condition
- Known congenital or documented QT prolongation
- Known allergy to soya or peanut due to the risk of hypersensitivity reactions

#### 4 Detail of intervention

Participants randomised to the usual care plus azithromycin arm will receive usual clinical care as per NHS guidelines, plus a course of oral azithromycin 500mg daily for three days. We will use the IMP distribution methods described in the protocol to deliver IMP to participants.

© Copyright: The University of Oxford and Oxford University Hospitals NHS Foundation Trust 2019

##### a. Investigational Medicinal Product (IMP) description

Azithromycin 250mg capsules. Participants in this arm will take 500 mg (two capsules) once daily for 3 days. The capsules are for oral administration.

Special instructions:
Azithromycin must be taken at least 1 hour before or 2 hours after antacids as this affects overall bioavailability. Azithromycin must be taken at least 1 hour before or 2 hours after food.

The marketing authorisation holder is: Teva UK Limited, Brampton Road, Hampden Park, Eastbourne, East Sussex, BN22 9AG, UK.
Marketing authorisation number: PL 00289/1570

##### b. Storage of IMP

Azithromycin: Stored at room temperature in locked cupboards in restricted access rooms in the Nuffield Department of Primary Care Health Sciences; in locked cupboards in restricted access rooms in GP Practices; in Pharmacies.

##### c. SmPC precautions and concomitant medication

###### i. Precautions

Azithromycin is a commonly prescribed antibiotic with an established safety profile. The SmPC advises caution using azithromycin in the following conditions:
Elderly people with proarrhythmic conditions due to the risk of developing cardiac arrhythmia and torsades de pointes including patients with congenital or documented QT prolongation; receiving treatment with other active substances known to prolong QT interval such as anti-arrhythmics (e.g. amiodarone and sotalol), cisapride, and fluoroquinolones such as moxifloxacin and levofloxacin; known hypokalaemia and hypomagnesaemia; significant hepatic or renal impairment; patients with neurological or psychiatric disorders; myasthenia gravis. Azithromycin as other with the use of nearly all antibacterial agents, alters the normal flora of the colon leading to overgrowth of Clostridium difficile which can lead to Clostridium difficile associated diarrhoea.

###### ii. Concomitant medications

Effects of other medicinal products on azithromycin:

*Antacids*

In a pharmacokinetic study investigating the effects of simultaneous administration of antacids and azithromycin, no effect on overall bioavailability was seen, although the peak serum concentrations were reduced by approximately 25%. In patients receiving both azithromycin and antacids, the medicinal products should not be taken simultaneously. Azithromycin must be taken at least 1 hour before or 2 hours after the antacids.

Co-administration of azithromycin prolonged-release granules for oral suspension with a single 20 ml dose of co-magaldrox (aluminium hydroxide and magnesium hydroxide) did not affect the rate and extent of azithromycin absorption.

Co-administration of a 600 mg single dose of azithromycin and 400 mg efavirenz daily for 7 days did not result in any clinically significant pharmacokinetic interactions.

© Copyright: The University of Oxford and Oxford University Hospitals NHS Foundation Trust 2019

*Fluconazole*

Co-administration of a single dose of 1200 mg azithromycin did not alter the pharmacokinetics of a single dose of 800 mg fluconazole. Total exposure and half-life of azithromycin were unchanged by the coadministration of fluconazole, however, a clinically insignificant decrease in $C_{max}$ (18%) of azithromycin was observed.

*Nelfinavir*

Co-administration of azithromycin (1200 mg) and nelfinavir at steady state (750 mg three times daily) resulted in increased azithromycin concentrations. No clinically significant adverse effects were observed and no dose adjustment is required.

*Rifabutin*

Coadministration of azithromycin and rifabutin did not affect the serum concentrations of either medicinal product.

Neutropenia was observed in subjects receiving concomitant treatment of azithromycin and rifabutin. Although neutropenia has been associated with the use of rifabutin, a causal relationship to combination with azithromycin has not been established.

*Terfenadine*

Pharmacokinetic studies have reported no evidence of an interaction between azithromycin and terfenadine. There have been rare cases reported where the possibility of such an interaction could not be entirely excluded; however, there was no specific evidence that such an interaction had occurred.

*Cimetidine*

In a pharmacokinetic study investigating the effects of a single dose of cimetidine, given 2 hours before azithromycin, on the pharmacokinetics of azithromycin, no alteration of azithromycin pharmacokinetics was seen.

Effect of azithromycin on other medicinal products:
*Ergotamine derivatives*

Due to the theoretical possibility of ergotism, the concurrent use of azithromycin with ergot derivatives is not recommended.

*Digoxin and colchicine (P-gp substrates)*

Concomitant administration of macrolide antibiotics, including azithromycin, with P-glycoprotein substrates such as digoxin and colchicine, has been reported to result in increased serum levels of the P-glycoprotein substrate. Therefore, if azithromycin and P-gp substrates such as digoxin are administered concomitantly, the possibility of elevated serum concentrations of the substrate should be considered.

*Coumarin-Type Oral Anticoagulants*

In a pharmacokinetic interaction study, azithromycin did not alter the anticoagulant effect of a single 15-mg dose of warfarin administered to healthy volunteers. There have been reports received in the post-marketing period of potentiated anticoagulation subsequent to co-administration of azithromycin and coumarin-type oral anticoagulants. Although a causal relationship has not been established, consideration should be given to the frequency of

© Copyright: The University of Oxford and Oxford University Hospitals NHS Foundation Trust 2019

monitoring prothrombin time when azithromycin is used in patients receiving coumarin-type oral anticoagulants.

*Cyclosporin*

In a pharmacokinetic study with healthy volunteers that were administered a 500 mg/day oral dose of azithromycin for 3 days and were then administered a single 10 mg/kg oral dose of cyclosporin, the resulting cyclosporin $C_{max}$ and $AUC_{0-5}$ were found to be significantly elevated. Consequently, caution should be exercised before considering concurrent administration of these drugs. If coadministration of these drugs is necessary, cyclosporin levels should be monitored and the dose adjusted accordingly.

*Theophylline*

There is no evidence of a clinically significant pharmacokinetic interaction when azithromycin and theophylline are co-administered to healthy volunteers. As interactions of other macrolides with theophylline have been reported, alertness to signs that indicate a rise in theophylline levels is advised.

*Trimethoprim/sulfamethoxazole*

Coadministration of trimethoprim/sulfamethoxazole DS (160 mg/800 mg) for 7 days with azithromycin 1200 mg on Day 7 had no significant effect on peak concentrations total exposure or urinary excretion of either trimethoprim or sulfamethoxazole. Azithromycin serum concentrations were similar to those seen in other studies.

*Zidovudine*

Single 1000 mg doses and multiple 1200 mg or 600 mg doses of azithromycin had little effect on the plasma pharmacokinetics or urinary excretion of zidovudine or its glucuronide metabolite. However, administration of azithromycin increased the concentrations of phosphorylated zidovudine, the clinically active metabolite, in peripheral blood mononuclear cells. The clinical significance of this finding is unclear, but it may be of benefit to patients.

Azithromycin does not interact significantly with the hepatic cytochrome P450 system. It is not believed to undergo the pharmacokinetic drug interactions as seen with erythromycin and other macrolides. Hepatic cytochrome P450 induction or inactivation via cytochrome-metabolite complex does not occur with azithromycin.

*Astemizole, alfentanil*

There are no known data on interactions with astemizole or alfentanil. Caution is advised in the co-administration of these medicines with azithromycin because of the known enhancing effect of these medicines when used concurrently with the macrolid antibiotic erythromycin.

*Atorvastatin*

Coadministration of atorvastatin (10 mg daily) and azithromycin (500 mg daily) did not alter the plasma concentrations of atorvastatin (based on a HMG CoA-reductase inhibition assay).

However, post-marketing cases of rhabdomyolysis in patients receiving azithromycin with statins have been reported.

*Carbamazepine*

© Copyright: The University of Oxford and Oxford University Hospitals NHS Foundation Trust 2019

In a pharmacokinetic interaction study in healthy volunteers, no significant effect was observed on the plasma levels of carbamazepine or its active metabolite in patients receiving concomitant azithromycin.

*Cisapride*

Cisapride is metabolized in the liver by the enzyme CYP 3A4. Because macrolides inhibit this enzyme, concomitant administration of cisapride may cause the increase of QT interval prolongation, ventricular arrhythmias and torsades de pointes.

*Cetirizine*

In healthy volunteers, coadministration of a 5-day regimen of azithromycin with cetirizine 20 mg at steady-state resulted in no pharmacokinetic interaction and no significant changes in the QT interval.

*Didanosins (Dideoxyinosine)*

Coadministration of 1200 mg/day azithromycin with 400 mg/day didanosine in 6 HIV-positive subjects did not appear to affect the steady-state pharmacokinetics of didanosine as compared with placebo.

*Efavirenz*

Coadministration of a 600 mg single dose of azithromycin and 400 mg efavirenz daily for 7 days did not result in any clinically significant pharmacokinetic interactions.

*Indinavir*

Coadministration of a single dose of 1200 mg azithromycin had no statistically significant effect on the pharmacokinetics of indinavir administered as 800 mg three times daily for 5 days.

*Methylprednisolone*

In a pharmacokinetic interaction study in healthy volunteers, azithromycin had no significant effect on the pharmacokinetics of methylprednisolone.

*Midazolam*

In healthy volunteers, coadministration of azithromycin 500 mg/day for 3 days did not cause clinically significant changes in the pharmacokinetics and pharmacodynamics of a single 15 mg dose of midazolam.

*Sildenafil*

In normal healthy male volunteers, there was no evidence of an effect of azithromycin (500 mg daily for 3 days) on the AUC and $C_{max}$ of sildenafil or its major circulating metabolite.

*Triazolam*

In 14 healthy volunteers, coadministration of azithromycin 500 mg on Day 1 and 250 mg on Day 2 with 0.125 mg triazolam on Day 2 had no significant effect on any of the pharmacokinetic variables for triazolam compared to triazolam and placebo.

© Copyright: The University of Oxford and Oxford University Hospitals NHS Foundation Trust 2019

##### iii. Fertility, pregnancy and lactation

*Pregnancy*

There are no adequate data from the use of azithromycin in pregnant women. In reproduction toxicity studies in animals azithromycin was shown to pass the placenta, but no teratogenic effects were observed. The safety of azithromycin has not been confirmed with regard to the use of the active substance during pregnancy. Therefore azithromycin should only be used during pregnancy if the benefit outweighs the risk.

#### 5 Safety reporting

Mechanisms for safety reporting are outlined in the protocol. In brief, we will collect symptoms and side effects of azithromycin from symptom diaries and participant telephone calls.

Common symptoms of azithromycin include diarrhoea, abdominal pain, nausea and flatulence. It may also cause headache, dizziness, insomnia, altered taste, pins and needles, changes in vision or hearing, rash, itching, joint pains or fatigue.

© Copyright: The University of Oxford and Oxford University Hospitals NHS Foundation Trust 2019

#### 21 APPENDIX F: USUAL CARE PLUS DOXYCYCLINE ARM (DISCONTINUED)

##### 1. Background and rationale

###### a. Evidence for potential doxycycline benefits in COVID-19

Doxycycline may be beneficial in the treatment of COVID-19 patients, and especially those in the at-risk or age range of the PRINCIPLE trial.
The rationale for testing doxycycline is based on three reasons:
Firstly, doxycycline may have direct antiviral activity against SARS-CoV-2 based on computer modelling. Analysing all the proteins encoded by SARS-CoV-2 genes and then predicting potential targets by performing target-based virtual ligand screening, doxycycline ranked in the group of compounds with the highest binding affinity to 3CLpro (3-chymotrypsin-like protease). 3CLpro is the main protease in SARS- CoV-2 which is critical in the life-cycle of the virus (35).

Secondly, doxycycline has known anti-inflammatory effects in various human diseases by inhibiting mitogen-activated protein kinase (MAPK) and SMAD pathways (36), as well as potent antioxidant properties(37). Doxycycline reduces the hyperinflammation associated with severe COVID-19 by antagonising metalloproteinases such as MMP9 that are linked with lung injury, including SARS and ARDS(38).

Lastly, from extensive experience in other infectious diseases, doxycycline has broad antimicrobial activity and is efficacious against a broad spectrum of bacteria including atypical bacteria and other pathogens including intracellular plasmodia, chlamydia, rickettsia, and RNA viruses like Dengue fever and chikungunya.

###### b. Importance of treating CAP or CAP risk in the elderly or immuno-compromised

An important secondary pathway to severe illness and death with COVID-19 may be secondary infection and sepsis in the immune-compromised state, especially secondary community or hospital acquired pneumonia. Older people are more susceptible to pneumonia because of comorbidities, a weakened immune system and are therefore more likely to die.(32) The onset of pneumonia in the elderly can often be rapid, and for severe pneumonia, the prognosis is poor: as many as one in five will die.(32) Severe pneumonia is more prevalent the older you are and in those with more serious underlying diseases.(33) The leading cause of death is respiratory insufficiency. Death has been shown to increase in those not responding to initial antimicrobials, and consequently, the initial selection of the agent is important. Common causative organisms in the elderly admitted to the hospital with pneumonia include *Haemophilus influenza, Staphylococcus aureus, Streptococcus pneumoniae* and less commonly, atypical organisms, such as *Mycoplasma pneumoniae* and *Klebsiella pneumoniae*. All these organisms fall under doxycycline's antimicrobial spectrum.

We are aware that currently NICE, in their COVID-19 rapid guideline, advocates that clinicians offer oral doxycycline for treatment of suspected pneumonia in people who can or wish to be

© Copyright: The University of Oxford and Oxford University Hospitals NHS Foundation Trust 2019

treated in the community if: the likely cause is bacterial or; it is unclear whether the cause is bacterial or viral and symptoms are more concerning or; they are at high risk of complications (older or frail patients, pre-existing comorbidity or have a history of severe illness following previous lung infection).(39) Doxycycline will have at least as broad a spectrum of action as azithromycin in terms of bacterial infections with the potential anti-viral and anti-inflammatory effects.

Doxycycline for acute cough and community acquired pneumonia is recommended in the British National Formulary at a dose of Doxycycline 200mg stat then 100mg daily for the next 4 days. However, its use in COVID-19 is not proven and therefore important to address in this trial. Given the potential anti-inflammatory properties of doxycycline, we will use a slightly extended 7 day course.

#### 2. Changes to outcome measures

The addition of this usual care plus doxycycline arm will not require any changes to outcome measures

#### 3. Eligibility criteria specifically related to doxycycline

Inclusion criteria: No changes
Exclusion criteria:

- Pregnancy
- Breastfeeding
- Myasthenia gravis
- Systemic lupus erythematosus
- Previous adverse reaction to, or currently taking, doxycycline or other tetracyclines
- Sucrose intolerance (i.e. rare hereditary problems of fructose intolerance, glucose galactose malabsorption or sucrose-isomaltase insufficiency)
- Already taking antibiotics for an acute condition
- Patients taking the following drugs: ciclosporin, retinoids (acitretin, alitretinoin, isotretinoin, tretinoin), methotrexate, ergotamine, methoxyflurane, lithium.

#### 4. Detail of intervention

Participants randomised to the usual care plus doxycycline arm will receive usual clinical care as per NHS guidelines, plus a course of oral doxycycline for 7 days. We will use the IMP distribution methods described in the protocol to deliver IMP to participants.

##### a. Investigational Medicinal Product (IMP) description

Doxycycline 100mg capsules. Participants in this arm will take 200mg on the first day (as a single dose or in divided doses with a twelve hour interval) followed by 100mg a day for 6 days (7 day course in total). The capsules are for oral administration.

© Copyright: The University of Oxford and Oxford University Hospitals NHS Foundation Trust 2019

Special instructions:
Capsules should be swallowed whole with plenty of fluid, while sitting or standing. Capsules should be taken during meals, well before going to bed. Due to the risk of photosensitivity, patients should be advised to avoid exposure to sunlight or sun lamps.

The marketing authorisation holder is:

Accord-UK Ltd (Trading style: Accord), Whiddon Valley, Barnstaple, Devon, EX32 8NS
Marketing authorisation number: PL 0142/0407

##### b. Storage of IMP

Doxycycline: Stored at room temperature in locked cupboards in restricted access rooms in the Nuffield Department of Primary Care Health Sciences; in locked cupboards in restricted access rooms in GP Practices; in Pharmacies.

##### c. SmPC precautions and concomitant medication

###### i. Precautions

Doxycycline is a commonly prescribed antibiotic with an established safety profile. The SmPC states that in elderly patients "doxycycline may be prescribed in the usual dose with no special precautions. No dosage adjustment is necessary in the presence of renal impairment".

###### ii. Concomitant medications

*Warfarin*
There have been reports of prolonged prothrombin time in patients taking warfarin and doxycycline. Tetracyclines depress plasma prothrombin activity and reduced dosage of concomitant anti-coagulants may be necessary

#### 5. Safety reporting

Mechanisms for safety reporting are outlined in the protocol. In brief, we will collect symptoms and side effects from symptom diaries and participant telephone calls.

Common side effects of doxycycline include: Angioedema; diarrhoea; headache; Henoch-Schönlein purpura; hypersensitivity; nausea/vomiting; pericarditis; skin and photosensitivity reaction; dyspnoea; hypotension; peripheral oedema; tachycardia.

© Copyright: The University of Oxford and Oxford University Hospitals NHS Foundation Trust 2019

### 22 APPENDIX G: USUAL CARE PLUS INHALED CORTICOSTEROID (ICS) ARM

#### 1. Background and rationale

##### a. Evidence for potential benefits of inhaled corticosteroids in COVID-19 illness

Inhaled corticosteroids (ICS) are a commonly prescribed class of medication throughout the world. They are reasonably cheap and have been used widely for the last 60 years. The inhaled action and type2 pneumocyte target of COVID make ICS a potential therapeutic agent in COVID-19(40). They have been shown to be very effective in improving asthma and COPD care over the long term, where the recommendation is that most, if not all, patients with asthma should be prescribed an inhaled corticosteroid(41),(42) and up to 90% of patients with COPD in the UK are prescribed ICS(43). The rationale of ICS is to reduce the inflammatory process that underlies exacerbations, which can be triggered by viruses in asthma and COPD. Systemic corticosteroids have been found to be effective at reducing mortality amongst hospitalised patients with COVID-19 [46, 47], but it is not known whether pre-hospital treatment with ICS is also beneficial.

Further evidence is as described below:

**Evidence from the ARDS literature**

ICS in patients at risk of acute respiratory distress syndrome (ARDS) have been shown to improve physiology and reduce inflammatory markers(44). In patients admitted to hospital at risk of ARDS or acute lung injury, there was an almost 50% reduction of ARDS in patients that were using ICS pre-admission, even controlling for covariates such as age, gender and chronic respiratory disease(45). Moreover, this ICS effect can also be seen to improve pulmonary physiology(46).

**Potential mechanism of efficacy**

Recently published *in vitro* data suggest a role for ICS inhibition of coronavirus replication in infected epithelial cells(47), whilst there is an indication that there is accelerated hyperinflammation at the onset of SARS-CoV-2 infection(48), which potentially can be modified by anti-inflammatory therapy. This suggests a plausible mechanism for ICS efficacy against COVID-19 in which ICS has a dual role: firstly, toning down the inflammatory “runaway train” (ARDS-like) response affecting a minority of COVID-19 patients; and secondly, inhibiting viral replication. It has long been known that the ICS effect on epithelial cells is as a direct consequence of gene transcription(49), and investigation of gene expression of ACE2 and TMPRSS2 in the sputum of asthmatic patients has very recently demonstrated lower expression of these key receptors in the presence of ICS(50). Furthermore, ICS attenuates expression of the ACE2 receptor in human and murine in vitro and in vivo models(51). This is of relevance as the SARS-CoV-2 mechanism of action is upon direct action of the ACE2 receptor, a receptor highly expressed on epithelial cells in the oral mucosa and type 2 alveolar cells and the serine protease TMPRSS2 for SARS-CoV-2 spike protein priming(52, 53). Furthermore, there is experimental evidence that inhaled corticosteroids inhibit coronavirus replication in vitro(54, 55). SARS-CoV-2 binds to cells via the angiotensin converting enzyme 2 (ACE2) receptor. ACE2 is highly expressed on epithelial cells in the oral mucosa and type 2 alveolar epithelial cells. The use of inhaled corticosteroids as a therapy suggests it would target the cells of interest. Furthermore, the primary action of the inhaled steroids is on the type 2 pneumocytes where viral replication is going to be at its most, where we know that ACE2 receptor expression is high.

© Copyright: The University of Oxford and Oxford University Hospitals NHS Foundation Trust 2019

#### 2. Changes to outcome measures

The addition of this arm will not require any changes to outcome measures.

#### 3. Eligibility criteria specifically related to ICS

Inclusion criteria:

Age criteria: Patients aged ≥65 years, or Patients aged 50-64 years and meeting at least one of the following criteria:

- Known weakened immune system due to a serious illness or medication (e.g. chemotherapy);
- Known heart disease and/or a diagnosis of high blood pressure;
- Known asthma or lung disease;
- Known diabetes;
- Known mild hepatic impairment;
- Known stroke or neurological problem;
- Self-report obesity or body mass index ≥35 kg/m$^2$

Exclusion criteria:

- A known allergy to inhaled corticosteroids
- Any known contraindication to inhaled corticosteroids (as per SmPC, *patients with rare hereditary problems of galactose intolerance, the Lapp lactase deficiency or glucose-galactose malabsorption should not take this medicine. Lactose, the excipient in the product, contains small amounts of milk proteins and can therefore cause allergic reactions*).
- Patient currently prescribed inhaled or systemic corticosteroids
- Unable to administer inhaler

#### 4. Detail of intervention

Participants randomised to the usual care plus ICS arm will receive usual clinical care as per NHS guidelines, plus inhaled corticosteroids for 14 days. We will use the IMP distribution methods described in the protocol to deliver IMP to participants.

##### a. Investigational Medicinal Product (IMP) description

The IMP is the inhaled corticosteroid budesonide (dose 400mcg, Pulmicort turbohaler®). Inhaled budesonide comes in a polyethylene container consisting of a white cover screwed onto a brown bottom plate. Inside this is the inhaler with its main parts: a mouthpiece, a dosing mechanism and a substance store. The device will have 50 actuations of 400mcg/actuation. This product has marketing authorisation in the UK (PL 17901/0164) and is manufactured by AstraZeneca UK Ltd, 600 Capability Green, Luton, LU1 3LU, UK. This IMP will be taken as 2 puffs twice a day for 14 days.

##### b. Storage of IMP

© Copyright: The University of Oxford and Oxford University Hospitals NHS Foundation Trust 2019

Stored at room temperature in locked cupboards in restricted access rooms in the Nuffield Department of Primary Care Health Sciences; in locked cupboards in restricted access rooms in GP practices; in Pharmacies

**c. SmPC precautions and concomitant medication**

**iii. Precautions**

Budesonide is a commonly prescribed inhaled steroid with an established safety profile.

**iv. Concomitant medications**

Largely, there is no restriction to concomitant medications using inhaled budesonide. The SmPC states that concomitant treatment with ketoconazole, HIV protease inhibitors or other potent CYP3A inhibitors may increase systemic budesonide levels, but that this is of little clinical significance for a short term treatment of 2 weeks, which is the duration of IMP use in the trial.

**5. Safety reporting**

Mechanisms for safety reporting are outlined in the protocol. In brief, we will collect symptoms and side effects from symptom diaries and participant telephone calls.

Common and/or potential side effects from IMP include:

- Cough immediately after inhaling
- Mouth and throat pain
- Hoarse voice
- Oral candidiasis (thrush)

These are all reversible upon ceasing IMP.

© Copyright: The University of Oxford and Oxford University Hospitals NHS Foundation Trust 2019

### 23 APPENDIX H: USUAL CARE PLUS COLCHICINE

#### 1. Background and rationale

##### a. Evidence for potential benefits of colchicine in COVID-19 illness

Colchicine is licenced and widely used in the UK for the treatment of acute gout and has been investigated as a possible treatment for COVID-19. Reyes and colleagues (56) have summarised existing clinical evidence for colchicine for COVID-19 thus:

"A retrospective single-centre study of 87 ICU patients with COVID-19 demonstrated a lower risk of death in patients on colchicine (adjusted HR 0.41, 95% CI 0.17 to 0.98).(57) The Greek Effects of Colchicine in COVID-19 (GRECCO-19) trial was the first prospective open-label randomised trial evaluating colchicine versus usual care in early hospitalised patients. This study of 105 patients found a significant reduction in the primary clinical outcome of a two-point deterioration on WHO disease severity scale.(58) An Italian study compared 122 hospitalised patients who received colchicine plus standard-of-care (lopinavir/ritonavir, dexamethasone or hydroxychloroquine) with 140 hospitalised patients receiving standard-of-care alone. Colchicine had a significant mortality benefit versus controls (84% vs 64% survival).(59) A third prospective study randomised 38 hospitalised COVID-19 patients to colchicine or placebo in a double-blinded manner.(60) Patients receiving colchicine had less need for supplemental oxygen at day 7 (6% vs 39%) and were more likely to be discharged at day 10 (94% vs 83%). Colchicine subjects also had greater reduction of CRP, and no increase in serious adverse events."

More recently, a systematic review and meta-analysis (in preprint) supports the notion that colchicine lowers the risk of mortality (HR of 0.25, 95% CI [0.09, 0.66], six studies, n=5,033) However, the summary point estimate from the three included RCTs showed a signal towards mortality benefit that was not statistically significant among patients receiving colchicine versus placebo (OR 0.49, 95% CI [0.20, 1.24]).(61)

The COLCORONA randomised clinical trial has now reported in a pre-print.(62) It randomised 4488 patients to treatment with colchicine (0.5 mg twice daily for 3 days and once daily thereafter) or placebo for 28 days. The primary endpoint occurred in 4.7% of the patients in the colchicine group and 5.8% of those in the placebo group (odds ratio, 0.79; 95.1% confidence interval (CI), 0.61 to 1.03; P=0.08). Among the 4159 patients with PCR-confirmed COVID-19, the primary endpoint occurred in 4.6% and 6.0% of patients in the colchicine and placebo groups, respectively (odds ratio, 0.75; 95% CI, 0.57 to 0.99; P=0.04). In these patients with PCR-confirmed COVID-19, the odds ratios were 0.75 (95% CI, 0.57 to 0.99) for hospitalization due to COVID-19, 0.50 (95% CI, 0.23 to 1.07) for mechanical ventilation, and 0.56 (95% CI, 0.19 to 1.66) for death. Serious adverse events were reported in 4.9% and 6.3% in the colchicine and placebo groups (P=0.05); pneumonia occurred in 2.9% and 4.1% of patients (P=0.02). Diarrhoea was reported in 13.7% and 7.3% in the colchicine and placebo groups (P<0.0001).

This large-scale study of early treatment in those 40 years and over with symptoms of more severe illness or comorbidity suggests that colchicine treatment early on in the illness reduces the need for hospitalisation or COVID-19. However, the study did not assess impact on recovery, so we

© Copyright: The University of Oxford and Oxford University Hospitals NHS Foundation Trust 2019

don't know from this study if colchicine reduced symptom burden. This is important as those receiving colchicine, predictably, experienced more gastrointestinal side-effects. The study did not recruit to target and some of the findings are not statistically significant. However, these findings need replication before this drug can be considered for routine use for COVID-19.

Another Phase 3 trial in Canadian pre-hospital and hospital settings is investigating colchicine paired with aspirin or interferon beta (ACTCOVID). The trial is still in progress (www2.phri.ca/ACT-COVID-19/).

##### *b.* Potential mechanism of action

Colchicine is a broad-spectrum anti-inflammatory agent.(63-66) Colchicine inhibits cellular transport and mitosis by binding to tubulin and preventing its polymerisation as part of the cytoskeleton transport system.(67) Several of the biological therapies that have been studied and/or used in the setting of severe COVID-19 target some of the same pathways as colchicine, including IL-1β (ie, anakinra) and IL-6 (ie, tocilizumab and sarilumab). Colchicine differs from these agents in having pleotropic mechanisms of action, being less potent on any single target, and being an oral agent. Potential benefits of colchicine compared to these biological therapies when used in the midst of cytokine storm, are that colchicine is not immunosuppressive, is not known to increase risk of infection, and is inexpensive.

There is evidence that the inflammasome is activated in COVID-19 and that the degree of activation is correlated with disease severity.(68) Inflammasomes are key components of effective host immune responses to pathogens. Excessive inflammasome activation (specifically NLRP3 inflammasome) is implicated in chronic inflammatory diseases such as inflammatory bowel disease, rheumatoid arthritis and gout, and with ARDS and ALI (acute lung injury) pathology following respiratory viral infections. Additionally, colchicine may have relevance to COVID-19 associated inflammatory pathology that include: inhibition of neutrophil chemotaxis in response to cytokines, inhibition of NFkB activation (a protein complex that controls transcription of DNA, cytokine production and cell survival) or expression, inhibition of neutrophil adhesion to endothelium, inhibition of neutrophil respiratory burst and reactive oxygen species generation, reduced TNF receptor expression on macrophages and endothelial cells and increased TGFβ expression(67) Of note though is that many of these latter actions of colchicine occur at much lower concentrations that are required for NLRP3 inflammasome activation in response to MSU crystals. Symptoms such as fever, joint and muscle ache, and headache may be ameliorated by a general anti-inflammatory action.

Therefore, when used early in the course of COVID-19, colchicine may prevent the progression from inflammatory activation to a hyperinflammatory state. The potential benefits of colchicine may therefore be maximised when used in the community, where earlier treatment could alleviate symptom burden, and prevent disease progression, hospitalisation and adverse outcomes.

#### 2. Changes to outcome measures

The addition of the usual care plus colchicine arm will not require any changes to outcome measures.

© Copyright: The University of Oxford and Oxford University Hospitals NHS Foundation Trust 2019

##### 3. Eligibility criteria specifically related to colchicine

Inclusion criteria: No changes required

Exclusion criteria:

- Hypersensitivity to the active substance or to any of the excipients listed in section (Lactose, Pregelatinised Maize Starch, Stearic Acid, Purified Talc, Purified Water, Ethanol 96%)
- Known or suspected pregnancy
- Breastfeeding
- Women of childbearing potential (premenopausal female that is anatomically and physiologically capable of becoming pregnant*) and not prepared to use highly effective contraception for the 28 day duration of follow up in the study**
- Known blood dyscrasias
- Known severe renal impairment or requiring dialysis
- Known severe hepatic impairment
- Currently taking any of the following drugs: colchicine, clarithromycin, erythromycin, ketoconazole, itraconazole, voriconazole, HIV protease inhibitors (e.g. ritonavir, atazanavir), cobicistat, verapamil, diltiazem, cyclosporin, quinidine, disulfiram, grapefruit juice
- Inflammatory bowel disease or chronic diarrhoea

* As recorded by the participant on the screening form and confirmed on Day 3 telephone call

***Highly effective methods have typical-use failure rates of less than 1% and include male or female sterilisation and long-acting reversible contraceptive (LARC) methods (intrauterine devices and implants). Women using another method of contraception, such as a combined hormonal method, progestogen only pill or injection, are eligible* ***if*** *they are willing to use an additional barrier method (e.g. male condom) for the 28 day duration of follow-up in the trial.*

The Patient Information Sheet Appendix, which the participant must read prior to providing informed consent, will clearly state the exclusion criteria listed above and the participant will be asked if they meet any of these exclusion criteria at the screening stage of the trial. The assessing clinician will then review the participant's responses against their medical record to confirm eligibility.

##### 4. Detail of intervention

Participants randomised to the usual care plus colchicine arm will receive usual clinical care as per NHS guidelines, plus 500 micrograms of colchicine to be taken each day for 14 days. We will use the IMP distribution methods described in the protocol to deliver IMP to participants.

Pharmacokinetic modelling shows that for a given dose of colchicine plasma exposure is greater in older (especially female) people than younger people; the target patient population for this study will be predominantly older subjects. Colchicine is also subject to accumulation in leucocytes- a target cell in this COVID19. Taking these factors into account and given the relatively

© Copyright: The University of Oxford and Oxford University Hospitals NHS Foundation Trust 2019

narrow therapeutic index of colchicine, the dosing regimen will be 500 microgrammes daily for 14 days. A loading dose such as that used in the COLCORONA study and the use of a higher daily dose increases the risk of dose related adverse drug reactions (ADRs). Given the outpatient setting for this study with limited opportunity for laboratory and clinical monitoring, the proposed dose is expected to achieve a balance between clinically meaningful exposure in target cells while minimising the risk of ADRs. Some other studies have treated for longer than 14 days (7) but the natural history of COVID19 is that in most cases the course of disease for that individual has been established within 2 weeks.

It is acknowledged that the total dose of colchicine administered in this study (7 mg) is modestly greater than that recommended for treatment of acute gout (6 mg) but the proposed regimen has been designed to minimise the risk of ADRs.

Note: The British National Formulary advises a maximum total of 6mg per treatment course for acute gout (1mg less than the total for this study). However, the treatment course for gout is given over up to three days at 500mcg 2-4 times per day initially. In the PRINCIPLE Trial, the treatment will be spread over two weeks at a lower daily dose.

We propose a shorter duration and no loading dose compared to the COLCORONA study (62), given the incidence of side-effects found in that study, and that by two weeks, most patients with COVID-19 have either recovered or been hospitalised. Therefore, the window of opportunity for a positive benefit is mainly over two weeks, and a shorter duration without a loading dose will minimize risk of side-effects, while offering potential benefit. There are no dose-findings studies for colchicine in COVID-19. Our proposed dosing regime is based on expert pharmacological opinion and an appraisal of side-effects balanced against potential benefit in a large-scale community study without face-to-face recruitment and monitoring.

**a. Investigational Medicinal Product (IMP) description**

Colchicine 500 microgram (µg) tablets. The tablets are for oral administration. One tablet to be taken daily by mouth for 14 days (14 tablets in total).

Special instructions: Tablets should be swallowed whole with a glass of water.

**Manufacturer:**
The Marketing Authorisation holder is:
Accord-UK Ltd
(Trading style: Accord)
Whiddon Valley
Barnstaple
Devon
EX32 8NS
Marketing authorisation number is: PL 0142/0918

**Labelling and QP release:**
Vertical Pharma Resources Ltd (trading as IPS Pharma), 41 Central Avenue, West Molesey, KT8 2QZ, UK

© Copyright: The University of Oxford and Oxford University Hospitals NHS Foundation Trust 2019

Authorisation number: WDA(H) 32879

**b. Storage of IMP**

Colchicine: This medicine does not need any special storage conditions, but we will ask participants to store the medication at room temperature. The medication will be stored in locked cupboards in restricted access rooms in the Nuffield Department of Primary Care Health Sciences; in locked cupboards in restricted access rooms in GP Practices; in Pharmacies.

**c. SmPC Precautions, concomitant medications, pregnancy and lactation**

**i. SmPC Precautions**

Colchicine is a commonly prescribed drug in UK primary care and has a well-described safety profile due to its regulatory assessments for the authorisation in gout. Typical treatment doses for acute gout are 500 micrograms 2–4 times a day until symptoms relieved, maximum 6 mg per course.

Colchicine is teratogenic in animal studies and contraindicated in patients with severe renal (including patients undergoing haemodialysis) or severe hepatic impairments. Colchicine may cause severe bone marrow depression (agranulocytosis, aplastic anaemia, thrombocytopenia) and blood cell dyscrasia.

Colchicine is potentially toxic with a narrow therapeutic window.
Symptoms of acute overdosage may be delayed (3 hours on average): nausea, vomiting, abdominal pain, haemorrhagic gastroenteritis, volume depletion, electrolyte abnormalities, leucocytosis, hypotension in severe cases. The second phase with life threatening complications develops 24 to 72 hours after drug administration: multisystem organ dysfunction, acute renal failure, confusion, coma, ascending peripheral motor and sensory neuropathy, myocardial depression, pancytopenia, dysrhythmias, respiratory failure, consumption coagulopathy.

Patients at particular risk of toxicity are those with renal or hepatic impairment, gastrointestinal or cardiac disease, and patients at extremes of age. However, colchicine has a good safety profile when used according to the established therapeutic guidelines, and toxicity is rare if the recommended doses are not exceeded.

In PRINCIPLE, we exclude patients with known severe renal and known liver impairment, and have used a cautious dosing regimen, with no loading dose and low daily dose, to minimise risk in participants with other less severe co-morbidities. Our dosing schedule is also shorter in duration than the 30 days used in the large scale, remotely managed COLCORONA trial.(62) In addition, we will mitigate the risk of toxicity by asking each participant taking colchicine the number of tablets remaining via their diary entry to ensure drug accountability.

**ii. Concomitant medications**

Colchicine is a substrate for both CYP3A4 and the transport protein P-gp. In the presence of CYP3A4 or P-gp inhibitors, the concentrations of colchicine in the blood increase. Toxicity,

© Copyright: The University of Oxford and Oxford University Hospitals NHS Foundation Trust 2019

including fatal cases, have been reported during concurrent use of CYP3A4 or P-gp inhibitors such as macrolides (clarithromycin and erythromycin), ciclosporin, ketoconazole, itraconazole, voriconazole, HIV protease inhibitors, calcium channel blockers (**verapami**l and **diltiazem**) and disulfiram.
Colchicine is contraindicated in patients with renal or hepatic impairment who are taking a P-gp inhibitor (e.g. ciclosporin, verapamil or quinidine) or a strong CYP3A4 inhibitor (e.g. ritonavir, atazanavir, indinavir, clarithromycin, telithromycin, itraconazole or ketaconazole).
Therefore, patients using the above medications will not be eligible for enrolment into the colchicine arm of PRINCIPLE.

**iii. Fertility, pregnancy and lactation**

Pregnancy and breast-feeding are exclusions for the colchicine arm.

Women of childbearing potential (premenopausal female that is anatomically and physiologically capable of becoming pregnant) and not prepared to use highly effective contraception for the 28 day duration of follow up in the study are excluded from the trial.

#### 5. Safety reporting

Mechanisms for safety reporting are outlined in the protocol. In brief, we will collect symptoms and side effects from symptom diaries and participant telephone calls.

<u>Common and/or potential side effects that may be associated with colchicine include:</u>

Abdominal pain, diarrhoea*, nausea, vomiting.

A meta-analysis of 35 randomised trials of colchicine versus placebo found that the most common and significant adverse effect was diarrhoea. The only other adverse effect that occurred at a greater frequency than placebo was a set of pooled gastrointestinal symptoms including nausea, vomiting, diarrhoea, abdominal pain, loss of appetite, and bloating.(69, 70)

<u>Rare side effects that may be associated with colchicine:</u>

Agranulocytosis; alopecia; bone marrow disorders; gastrointestinal haemorrhage; kidney injury; liver injury; menstrual cycle irregularities; myopathy; nerve disorders; rash; sperm abnormalities; thrombocytopenia

** Side effects that may be associated with COVID-19*

We will report SAEs as defined in the main protocol for hospitalisation and/or death. Participants record symptoms and adverse events on their diary card. Events rated as a 'major problem' will be assessed by a clinician for potential reporting as an SAE.

**Drug Accountability**

© Copyright: The University of Oxford and Oxford University Hospitals NHS Foundation Trust 2019

We will telephone all participants on Day 3 after randomisation to confirm that they have received their medication. For those receiving a trial treatment, 3 attempts are made to contact the participant to confirm receipt of the medication.

If we are unable to contact patients in the colchicine group, we will confirm and log IMP receipt by checking the patient's daily diary, where they are asked on a daily basis whether they have taken their trial treatment and how many tablets they have left. We can also check via the DHL portal, whether the participant pack containing the medication has been received by the participant, for additional confirmation. IMP receipt will be logged on the central IMP log.

© Copyright: The University of Oxford and Oxford University Hospitals NHS Foundation Trust 2019

### 24 APPENDIX I: USUAL CARE PLUS FAVIPIRAVIR

#### 1. Background and rationale

##### a. Evidence for potential benefits of favipiravir in COVID-19 illness

Small clinical trials assessing favipiravir for treatment of COVID-19 have been published, but results are inconclusive.

A randomised controlled trial in China of 240 adults hospitalised with moderate to severe COVID-19 compared favipiravir (1600mg BD day 1; 600mg BD days 2-7) with umifenovir (200mg TDS).(71) In the per-protocol analysis, there was no difference in the primary outcome of clinical recovery by day 7 between the two arms. In a post-hoc analysis restricted to patients with moderate COVID-19, 70/98 (71.43%) had recovered by day 7 in the favipiravir arm, vs 62/111 (55.86%) in the umifenovir arm ($p = 0.0199$). In other post-hoc analyses, time to cessation of fever ($p < 0.0001$) and cough ($p < 0.0001$) was shorter in the favipiravir arm. Adverse events were similar between the two arms, apart from hyperuricaemia which occurred in 13.8% of participants who received favipiravir vs 2.5% among those receiving umifenovir ($p < 0.001$).(71)

A trial in India of 150 patients with mild to moderate COVID-19 compared favipiravir (1800 mg BD day 1; 800 mg BD days 2-14) with usual care.(72) The primary outcome of time to viral clearance was not significantly different between the two arms (favipiravir 5 days [95% CI 4-7] vs usual care 7 days [95% CI 5-8, P=0.129), but there was a significant decrease in the secondary outcome of time to clinical cure in the favipiravir arm (favipiravir 3 days [95% CI 3- 4] vs usual care 5 days [95% CI 4-6], P=0.030). Adverse events were more common in the favipiravir arm (36% vs 8%); these were mainly hyperuricaemia (16.4%) and abnormal liver function tests (6.8%); 76.9% were mild and 23.1% were moderate.

A trial in Russia among 60 adults hospitalised with moderate COVID-19 compared usual care (n=20) vs lower dose favipiravir (1600 mg BD day 1; 600 mg BID days 2–14) (n=20) vs higher dose favipiravir (1800 mg BD day 1; 800 mg BD days 2–14) (n=20).(73) Viral clearance was higher in the favipiravir arms vs usual care by day 5 (25/40 [62.5%] vs 6/20 [30.0%], $p = 0.018$), although this difference was non-significant by day 10 (37/40 [92.5%] vs 16/20 [80.0%], $p = 0.155$). Temperature normalisation was quicker in the favipiravir arms (2 days [IQR 1–3] vs 4 days [IQR 1–8], $p = .007$). 17.5% of patients in the favipiravir arm experienced adverse drug reactions including gastrointestinal disturbances and raised liver function tests. Two patients discontinued the drug early.

A trial in Oman randomised 89 adults hospitalised with moderate to severe COVID-19 to receive favipiravir (1600 mg OD day 1; 600 mg BD days 2-10) combined with inhaled interferon beta-1b, versus hydroxychloroquine. There was no difference between the two groups with regard to the primary outcomes of inflammatory markers at discharge, length of hospital stay, transfer to ICU, or mortality. Adverse events were not reported.(74)

A smaller, non-randomised study among adults hospitalised with COVID-19 in China compared 35 patients treated with favipiravir (1600 mg BD day 1; 600 mg BD days 2-14) with 45 historical controls who were treated with lopinavir/ritonavir (400mg/100mg BD). All patients also

© Copyright: The University of Oxford and Oxford University Hospitals NHS Foundation Trust 2019

received aerosolised IFN-α. Viral clearance was quicker in the favipiravir cohort (median 4 days, IQR 2.5-9) compared to the lopinavir/ritonavir cohort (11 days, IQR 8-13, p <0.001). Improvements in CT chest imaging at 14 days were higher in the favipiravir group (91.4% vs 62.2%, p = 0.004).(75)

These mixed findings highlight the need for a large randomised controlled trial of favipiravir to treat COVID-19 in the community.

**b. Potential mechanism of efficacy**

Favipiravir is an oral antiviral that is licensed in Japan for use against novel and re-emerging influenzae, and has been used in clinical trials for Ebola (76).(76) Like remdesivir, it is a nucleoside analogue which selectively inhibits viral RNA polymerase, and has been shown to have in vitro activity against a range of RNA viruses (77, 78), including SARS-CoV-2.(9) Favipiravir was one of seven antiviral agents reported to achieve plasma concentrations at least double the reported concentrations required to inhibit 90% of SARS-CoV-2 replication in vitro.(79) In animal models, high dose favipiravir was found to reduce viral titres and lung pathology in SARS-CoV-2 infected hamsters.(80, 81) Given that upper respiratory tract SARS-CoV-2 viral loads peak in the first 3-5 days of illness,(82) antiviral treatments for COVID-19 may be of particular value early in the course of the disease.

#### 2. Changes to outcome measures

The addition of the *usual care plus favipiravir arm* will require the introduction of the secondary outcome measure, safety.

#### 3. Eligibility criteria specifically related to favipiravir

Inclusion criteria: No changes

Exclusion criteria:

- Aged <50 years
- Known or suspected pregnancy
- Breastfeeding
- Women of childbearing potential (premenopausal female that is anatomically and physiologically capable of becoming pregnant*), or male with a partner of childbearing potential, not willing to use highly effective contraceptive** for 28 day duration of the trial.
- Known allergy to favipiravir
- Currently taking favipiravir
- Known history of gout
- Known severe liver disease

* As recorded by the participant on the screening form and confirmed on Day 1 by a call between clinician and participant

© Copyright: The University of Oxford and Oxford University Hospitals NHS Foundation Trust 2019

** Highly effective methods have typical-use failure rates of less than 1% and include male or female sterilisation and long-acting reversible contraceptive (LARC) methods (intrauterine devices and implants) OR If a couple are using another method of contraception, such as a combined hormonal method, progestogen only pill or injection, they are only eligible if they are willing to use an additional barrier method (e.g. male condom) for the 28 day duration of follow-up in the trial.

Note: a barrier method on its own is not sufficient.

The Favipiravir Patient Information Sheet Appendix, which the participant must read prior to providing informed consent, will clearly state the exclusion criteria listed above and the participant will be asked if they meet any of these exclusion criteria at the screening stage of the trial including whether they have any known history of gout or known severe liver disease. The assessing clinician will then review the participant's responses against their medical record to confirm eligibility.

#### 4. Detail of intervention

Participants randomised to the usual care plus favipiravir arm will receive usual clinical care as per NHS guidelines, plus one batch of favipiravir for five days. We will use the IMP distribution methods described in the protocol to deliver IMP to participants.

##### a. Investigational Medicinal Product (IMP) description

Favipiravir 200 milligram (mg) tablets. The tablets are for oral administration. Nine tablets (1800mg) favipiravir to be taken twice a day on day one, and then four tablets (800mg) twice daily for four days (50 tablets in total).

This product is not licensed for use in the UK.

**Manufacturer:**
It is manufactured by FujifilmToyama Chemical Company Ltd., TOYAMA CHEMICAL CO., LTD. 2-5, Nishishinjuku 3-chome, Shinjuku-ku, Tokyo 160-0023, Japan.
Manufacturirng Licence No. NHI Number 87625
MA Approval No from outside the EEA: 22600AMX0053000 Japan

**Importer, Labelling and QP release:**
IPS Pharma, 41 Central Ave, East Molesey, West Molesey KT8 2QZ (32879)

##### b. Storage of IMP

All study medication is to be kept in a dry area, stored at 1° to 30°C (59° to 86°F) and shielded from direct light in locked cupboards in restricted access rooms in the Nuffield Department of Primary Care Health Sciences; in locked cupboards in restricted access rooms in GP practices; in Pharmacies. The IMP is stable at 1-30°C.

##### c. Precautions, concomitant medications, pregnancy and lactation

© Copyright: The University of Oxford and Oxford University Hospitals NHS Foundation Trust 2019

i. **Precautions**

Favipiravir has been used in over 30 clinical trials and has a favourable safety profile.(76) A review of six phase 2 and 3 controlled trials, including 4299 participants and 175 person-years-of-follow-up, found no statistically significant differences in overall proportion of AEs, SAEs, discontinuations due to AEs or LFT elevations between favipiravir and placebo or other treatment arms.(76) However, there was evidence of mild to moderate, asymptomatic, uric acid elevations among patients receiving favipiravir, although these generally returned to normal by 21 days. Overall, follow up times were short (5-21 days) and participants were generally young.

Patients with gout or a history of gout, and patients with hyperuricaemia (blood uric acid level may increase, and symptoms may be aggravated) are excluded.

The participant must avoid excessive exposure to sunlight or artificial ultraviolet light.

ii. **Concomitant medications**

Restrictions to paracetamol use (limiting daily use in adults to no more than 3000 mg/day) have been incorporated into all clinical study protocols. Participants will be advised they can't consume more than 6 paracetamol tablets in 24 hours.

iii. **Pregnancy and breastfeeding**

Evidence from animal models suggests that favipiravir has teratogenic potential, and there are no human studies of its use among pregnant or lactating women. The Japanese drug safety bureau advise that women of child-bearing potential should use effective contraception for up to 7 days after the end of treatment (https://www.cdc.gov.tw/File/Get/ht8jUiB_MI-aKnlwstwzvw). Male patients should use the most effective form of contraception (see eligibility criteria for examples) for up to seven days after the end of treatment if they have a female partner of child-bearing potential. Men should also avoid intercourse with pregnant women.

#### 5. Safety reporting

##### a. Side-effects

Mechanisms for safety reporting are outlined in the protocol. In brief, we will collect symptoms and side effects from symptom diaries and participant telephone calls.

Common and/or potential side effects from IMP include:

- Diarrhoea*[1]
- Nausea[1]
- Headache*[1]
- Urinary Tract Infections[1]
- Vomiting[1]
- Raised liver enzymes
- Elevated uric acid concentrations

*side-effects also seen with COVID-19

© Copyright: The University of Oxford and Oxford University Hospitals NHS Foundation Trust 2019

These are all reversible upon ceasing IMP.

Certain pre-defined moderate (defined above[1]) AEs experienced in the 5 days of favipiravir drug administration, will be reviewed daily and will be assessed by a clinician until resolution.

A systematic review suggests that liver changes do not differ between favipiravir and placebo/other treatments (76). To mitigate the risk of elevated uric acid concentrations, we exclude people with gout. Evidence shows mild to moderate, asymptomatic, uric acid elevations, which return to normal after stopping the medication (76).

We will report SAEs as defined in the main protocol for hospitalisation and/or death.

##### b. Reference Safety Information (RSI)

See section 7.13 of the Investigator Brochure. No serious adverse reactions are considered expected for the purpose of expedited reporting of suspected unexpected serious adverse reactions (SUSAR).

##### c. Drug Accountability

We telephone all participants on Day 3 after randomisation to confirm that they have received their medication and read the instructions on the medication card. For those receiving a trial treatment, 3 attempts are made to contact the participant to confirm receipt of the medication.

If we are unable to contact patients in the favipiravir group, we will confirm and log IMP receipt by checking the patient's daily diary, where they are asked on a daily basis whether they have taken their trial treatment, the number of tablets taken and the number of tablets remaining. We can also check via the DHL portal, whether the participant pack containing the medication has been received by the participant, for additional confirmation. IMP receipt will be logged on the central IMP log.

If a participant decides that they no longer wish to take their medication, we will provide a pre-paid envelope so that they can return the medication to the trial team, via courier.

##### d. Risk/Benefit Assessment

The UK COVID-19 Therapeutics Advisory Panel recommends including favipiravir into the PRINCIPLE platform with an 1800mg loading dose, followed by an 800mg BD maintenance dose, based on a review of efficacy and safety data.

###### i. Risks

Phase I studies and pharmacokinetic studies of favipiravir have indicated increased blood levels of uric acid and elevation of liver aminotransferases in some individuals (76). In addition, animal studies have indicated potential teratogenicity and the drug is distributed in sperm (83). Taking this evidence into account and to ensure patient safety we will exclude known pregnancy, breastfeeding, severe liver disease and known history of gout, and require participants to use adequate contraception for the duration of the treatment and 28 days of follow-up.

###### ii. Benefits

The benefits of favipiravir have been shown in several phase I-III studies to significantly alleviate influenza symptoms with a good safety profile (83). From the studies detailed

© Copyright: The University of Oxford and Oxford University Hospitals NHS Foundation Trust 2019

below, the frequency of adverse events was 386/1472 (26.2%) with favipiravir versus 227/894 (25.4%) with placebo and there were no differences in the incidence of any specific adverse event between groups.

A Phase I/II study in type A or B influenza using favipiravir doses 1800mg bd for 1 day followed by 800mg bd for 4 days (101 patients) or 2400mg/600mg/600mg for 1 day followed by 600mg tds for 4 days (82 patients) or placebo (88 patients).

Two Phase III studies in type A or B influenza using favipiravir doses 1800mg bd for 1 day followed by 800mg bd for 4 days (301 and 526 patients respectively) or placebo (322 and 169 patients respectively).

A global phase III study in type A or B influenza using favipiravir 1200mg/400mg for 1 day followed by 400mg bd for 4 days or oseltamivir 75mg bd for 5 days (377 and 380 patients respectively).

A phase II study in type A or B influenza using favipiravir doses 1000mg bd for 1 day followed by 400mg bd for 4 days (88 patients) or 1200mg bd for 1 day followed by 800mg bd for 4 days (121 patients) or placebo (124 patients).

Favipiravir was also used for Ebola virus disease, especially in the JIKI trial in Guinea (84). Doses (2400mg/2400mg/1200mg on Day 1 followed by 1200mg bd) were significantly higher than proposed for the current study. No drug related grade 3 or 4 clinical events were observed. In 41 of the 48 patients who survived, biochemical abnormalities of renal and liver function rapidly improved on treatment; 7 of 48 patients saw transient rises in one marker but all subsequently normalised despite continuing favipiravir. Biochemical abnormalities in patients who died were attributed to severe viral infection.

In London institutions such as Royal Free Hospital and Great Ormond Street Hospital (GOSH), favipiravir has been used as post-exposure prophylaxis for Ebola virus, for chronic norovirus infection, refractory influenza infection, astrovirus, respiratory syncytial virus and seasonal coronavirus. Occasional asymptomatic elevation of liver aminotransferase levels were observed, but no serious adverse events related to the drug. This includes in patients with immunodeficiencies who have received several months-worth of treatment (FLARE Trial).

Favipiravir has been used clinically in several viral infections and has shown beneficial treatment effects in both influenza and COVID-19 studies, as well as having a good safety profile.

© Copyright: The University of Oxford and Oxford University Hospitals NHS Foundation Trust 2019

### 25. Supplementary Material

#### A. Abbreviations

| | |
|---|---|
| AE | Adverse event |
| AR | Adverse reaction |
| CI | Chief Investigator |
| CRF | Case Report Form |
| CT | Clinical Trials |
| CTA | Clinical Trials Authorisation |
| CTRG | Clinical Trials and Research Governance |
| DMSC | Data Monitoring Committee / Data Monitoring and Safety Committee |
| DSUR | Development Safety Update Report |
| GCP | Good Clinical Practice |
| GP | General Practitioner |
| HRA | Health Research Authority |
| HCP | Healthcare professional |
| IB | Investigators Brochure |
| ICF | Informed Consent Form |
| ICH | International Conference on Harmonisation |
| IMP | Investigational Medicinal Product |
| ISA | Intervention Specific Appendix |
| MHRA | Medicines and Healthcare products Regulatory Agency |
| NHS | National Health Service |
| NIHR | National Institute of Health Research |
| RES | Research Ethics Service |
| PHE | Public Health England |
| PI | Principal Investigator |
| PIL | Participant/ Patient Information Leaflet |
| R&D | NHS Trust Research and Development Department |
| RCGP RSC | Royal College of General Practitioners Research Surveillance Centre |
| REC | Research Ethics Committee |
| RSI | Reference Safety Information |
| SAE | Serious Adverse Event |
| SAR | Serious Adverse Reaction |
| SDV | Source Data Verification |
| SMPC | Summary of Medicinal Product Characteristics |
| SOP | Standard Operating Procedure |
| TSC | Trial Steering Committee |
| SUSAR | Suspected Unexpected Serious Adverse Reactions |

© Copyright: The University of Oxford and Oxford University Hospitals NHS Foundation Trust 2019

| | |
|---|---|
| TMF | Trial Master File |

#### B. Key Trial Contacts

| | |
|---|---|
| **Chief Investigator** | Professor Chris Butler<br>Nuffield Department of Primary Care Health Sciences<br>Gibson Building<br>Radcliffe Observatory Quarter<br>Woodstock Road<br>Oxford<br>OX2 6GG<br> |
| **Sponsor** | Joint Research Office<br>1st floor, Boundary Brook House<br>Churchill Drive,<br>Headington<br>Oxford OX3 7GB<br><br>Tel: +44 (0)1865572224<br>Fax: +44 (0)1865572228 |
| **Funder(s)** | UKRI/NIHR |
| **Clinical Trials Unit** | Primary Care Clinical Trials Unit,<br>Nuffield Department of Primary Care Health Sciences<br>Radcliffe Observatory Quarter<br>Woodstock Road<br>Oxford<br>OX2 6GG<br><br>01865 289296 |
| **Statistician** | Dr Ben Saville,<br>Berry Consultants,<br>Austin, Texas, USA,<br>And<br>Department of Biostatistics,<br>Vanderbilt University School of Medicine,<br>Nashville, Tennessee,<br>USA.<br><br>Dr Ly-Mee Yu<br>Primary Care Clinical Trials Unit,<br>Nuffield Department of Primary Care Health Sciences<br>Radcliffe Observatory Quarter<br>Woodstock Road<br>Oxford<br>OX2 6GG |
| **Committees** | **DMSC Chair:**<br>Prof. Deborah Ashby<br>Chair in Medical Statistics and Clinical Trials<br>Director of the School of Public Health<br>Imperial College London Faculty of Medicine, School of Public Health,<br>153 Medical School |

© Copyright: The University of Oxford and Oxford University Hospitals NHS Foundation Trust 2019

| | |
|---|---|
| | St Mary's Campus<br>Imperial College London<br><br>(0)20 7594 8704<br><br>**DMSC Members:** Prof Simon Gates<br>Cancer Research Clinical Trials Unit (CRCTU)<br>Institute of Cancer and Genomic Sciences<br>University of Birmingham<br>Edgbaston<br>Birmingham<br>B15 2TT<br><br><br>Prof Gordon Taylor<br>College House,<br>University of Exeter, St Luke's Campus,<br>Heavitree Road,<br>Exeter, EX1 2LU, UK<br><br><br>Prof Nick Francis<br>Primary Care and Population Science,<br>University of Southampton, Southampton, UK.<br><br><br>Dr Patrick White<br>7th Floor<br>Capital House<br>Guy's<br>United Kingdom<br> |
| | **TSC Chair**<br>Prof Paul Little,<br>Primary Care and Population Science,<br>University of Southampton, Southampton, UK.<br><br><br>**TSC Members**<br>Prof Philip Hannaford,<br>NHS Professor of Primary Care<br>University of Aberdeen, Aberdeen, UK<br><br><br>Prof Matt Sydes,<br>Professor of Clinical Trials & Methodology,<br>MRC Clinical Trials Unit, University College of London, London, UK<br><br><br>*PPI representatives*<br>Ms Carol Green<br>Mr Tim Mustill |

© Copyright: The University of Oxford and Oxford University Hospitals NHS Foundation Trust 2019

#### C. Objectives and Outcome Measures

| | Objectives | Outcome Measures | Timepoint (s) |
|---|---|---|---|
| Primary | To assess the effectiveness of trial treatments in reducing<br>1) Time to recovery, for patients<br>2) Hospitalisation and/or death. | 1) Time to self-reported recovery, defined as the first instance that a participant reports feeling recovered from possible COVID-19, and<br>2) Hospitalisation and/or death | Within 28 days of randomisation<br>Patient report, Study Partner report, medical records, Daily online symptom scores |
| Secondary | To explore whether trial treatment will affect<br>1) Participant reported illness severity, reported by daily rating of how well participant feels.<br>2) Duration of severe symptoms and symptom recurrence<br>3) Contacts with the health services<br>4) Consumption of antibiotics<br>5) Hospital assessment without admission<br>6) Oxygen administration<br>7) Intensive Care Unit admission<br>8) Mechanical ventilation | 1-3. Participant reports daily and monthly (after 28 days) symptoms.<br>4. Contacts with health services reported by patients and/or captured by reports of patients' medical records if the practice is a member of the RCGP RSC network<br>5. Bi-weekly reports from participants' primary care medical records<br>6-10. Patient report/carer report/medical record in primary and secondary care<br>11. WHO-5 Well Being Index<br>12. Reports of new infections in the household (from daily questionnaire) | Daily online symptom scores.<br>Telephone call or text on days 2, 7, 14 and 28 and once a month for 12 months if data is not obtained through the online diary.<br><br>GP notes review if available through Oxford RCGP RSC network; otherwise, other sources of routinely collected data after 28 days. Medical notes review for up to 10 years.<br><br>HES/ONS/EMIS/Medical record data linkage after 28 days if patients have been assessed in hospital |

© Copyright: The University of Oxford and Oxford University Hospitals NHS Foundation Trust 2019

| | | | |
|---|---|---|---|
| | 9) Duration of hospital admission<br>10) Negative effects on well being<br>11) New infections in household<br>12) To determine if effects are specific to those with a positive test for SARS-CoV-2<br><br>13) To investigate the safety of treatments that are not licensed in the UK | 13. Swab test results will indicate an "Intention to Treat Infected" group within the overall cohort for sub analysis. Blood test results on recovery (optional) for evidence of historic COVID-19<br><br>Evaluation of overall safety of drugs by the monitoring of adverse events (AEs as defined in the ISAs) | Swab result from medical records, the supporting laboratory and/or convalescent blood test result for evidence of historic COVID-19<br><br>WHO 5 Well Being Index at baseline, day 14, and day 28 and monthly for up to 12 months, either via online diary or telephone<br><br>For the duration of the treatment course and a defined period after the treatment finishes (see ISAs) |
| Qualitative sub-study | 1. To explore patients' experiences of consulting, being tested and taking (trial) medication for possible COVID-19.<br>2. To explore healthcare professionals' views of taking part in research during pandemics. | 1. Telephone interviews with patients.<br>2. Telephone interviews with healthcare professionals. | 1. After 28 days.<br>2. Once practice has completed recruitment. |
| Intervention(s) | All trial interventions are detailed in the Appendices. Further interventions may be added or replaced during the course of the trial, subject to suitable interventions becoming available and all necessary approvals being obtained. | | |
| Comparator | In the first instance, this will be a two-arm trial, with the intervention arm being usual care plus a trial drug and the comparator being usual care. There will be no placebo control in this study. Additional arms may be added as the trial progresses. These will be detailed in the Appendices. If an intervention arm is shown to be superior, then this will become the new standard of care. | | |

© Copyright: The University of Oxford and Oxford University Hospitals NHS Foundation Trust 2019

| | |
|---|---|
| | However, the primary analysis of subsequent interventions will correspond to the comparison versus the original Usual Care arm. |

© Copyright: The University of Oxford and Oxford University Hospitals NHS Foundation Trust 2019

#### D. Adverse Events

#### Definitions

| | |
|---|---|
| Adverse Event (AE) | Any untoward medical occurrence in a participant to whom a medicinal product has been administered, including occurrences which are not necessarily caused by or related to that product. |
| Adverse Reaction (AR) | An untoward and unintended response in a participant to an investigational medicinal product which is related to any dose administered to that participant.<br><br>The phrase "response to an investigational medicinal product" means that a causal relationship between a trial medication and an AE is at least a reasonable possibility, i.e. the relationship cannot be ruled out.<br><br>All cases judged by either the reporting medically qualified professional or the Sponsor as having a reasonable suspected causal relationship to the trial medication qualify as adverse reactions. |
| Serious Adverse Event (SAE) | A serious adverse event is any untoward medical occurrence that:<br>• results in death<br>• is life-threatening<br>• requires inpatient hospitalisation or prolongation of existing hospitalisation<br>• results in persistent or significant disability/incapacity<br>• consists of a congenital anomaly or birth defect*.<br><br>Other ‘important medical events’ may also be considered a serious adverse event when, based upon appropriate medical judgement, the event may jeopardise the participant and may require medical or surgical intervention to prevent one of the outcomes listed above.<br><br>NOTE: The term "life-threatening" in the definition of "serious" refers to an event in which the participant was at risk of death at the time of the event; it does not refer to an event which hypothetically might have caused death if it were more severe.<br><br>*NOTE: Pregnancy is not, in itself an SAE. In the event that a participant or his/her partner becomes pregnant whilst taking part in a clinical trial or during a stage where the foetus could have been exposed to the medicinal product (in the case of the active substance or one of its metabolites having a long half-life) the pregnancy should be followed up by the investigator until delivery |

© Copyright: The University of Oxford and Oxford University Hospitals NHS Foundation Trust 2019

| | |
|---|---|
| | for congenital abnormality or birth defect, at which point it would fall within the definition of "serious". |
| Serious Adverse Reaction (SAR) | An adverse event that is both serious and, in the opinion of the reporting Investigator, believed with reasonable probability to be due to one of the trial treatments, based on the information provided. |
| Suspected Unexpected Serious Adverse Reaction (SUSAR) | A serious adverse reaction, the nature and severity of which is not consistent with the Reference Safety Information for the medicinal product in question set out:<br>• in the case of a product with a marketing authorisation, in the approved summary of product characteristics (SmPC) for that product<br>• in the case of any other investigational medicinal product, in the approved investigator's brochure (IB) relating to the trial in question. |

NB: To avoid confusion or misunderstanding the difference between the terms "serious" and "severe", the following note of clarification is provided: "Severe" is often used to describe intensity of a specific event, which may be of relatively minor medical significance. "Seriousness"

#### E. Data Recording and Record Keeping

The data will be entered into the CRFs in an electronic format by the participant, Trial Partner or trial team (using OpenClinica™ database via Sentry). OpenClinica™ is stored on a secure server – data will be entered in a web browser and then transferred to the OpenClinica Database by encrypted (Https) transfer. OpenClinica™ meets FDA part 11B standards. This includes safety data, laboratory data and outcome data. Safety data will also be collected through electronic diaries which are stored on a secure server.

Sentry is an online secure data entry system developed in-house at PC-CTU and hosted at Oxford. It is designed to collect sensitive data, such as participant and Trial Partner contact details, and securely retain them separate form a trial's clinical data. Sentry can also act as a central participant portal to manage online eligibility, eConsent and ePRO - acting as an intermediary between the participant and the clinical databases. Sentry is accessed via a secure HTTPS connection and all stored sensitive data is encrypted at rest to AES-256 standards. Participant and Trial Partner data will be kept and stored securely for as long as it's required by the study and reviewed on annual basis.

#### F. Qualitative Sub-study

With consent, participants will be contacted for a telephone interview within three months after they complete their day 28 follow up. The researcher will provide study information over the telephone and the Interview Patient PIS, and ICF will be available on the study website and emailed to participants if requested.

© Copyright: The University of Oxford and Oxford University Hospitals NHS Foundation Trust 2019

Once a practice has completed patient recruitment and one of their patients has been interviewed, we may ask 1-2 healthcare professionals who would be willing to share their experiences of taking part in the trial. Healthcare professionals will include clinicians and non-clinicians with the main criteria for inclusion in interviews being that HCP participants should have carried out trial activities in their practice. Potential HCP participants will be contacted in person or by email by the practice contact. They will be provided with the Interview HCP Invitation Email, Interview HCP PIS and Interview HCP ICF by email.

Patients recruited to both the intervention and usual care arms will be purposively sampled across the recruiting period with approximately 15-20 patients in each arm (30-40 interviews in total). We will seek to obtain maximum variation in age and symptom severity (as reported in daily diary at baseline). When the research team receives responses from HCPs, they will collect basic demographics to purposively select participants based on practice location, practice size, practice patient recruitment and job role. We aim to complete 20-25 interviews with HCPs.

All participants will only be required to take part in a single interview. Patient participant interviews will follow a semi-structured topic guide (Interview Patient Topic Guide) and ask about reasons for consulting and illness perceptions prior to the consultation, experiences of the consultation, the COVID-19 testing process (if applicable, and result if the participant has been notified) and medication adherence. The topic guide will be informed by the Common Sense Model which describes how people perceive and cope with symptoms of illness.
HCP interviews will follow the Interview HCP Topic Guide and will ask about experiences of carrying out trial activities, recruiting patients and the work required to set up a clinical trial during a pandemic.

Interviews with patient participants are expected to last approximately 30-45 minutes and interviews with HCPs are expected to last 15-30 minutes.

Data Collection:
Each interview will be audio-recorded with the participant's permission. Recordings will allow verbatim transcription of interviews. Transcription will be completed by an independent transcription company. Once transcribed and transcripts are checked, audio-recordings will be deleted. Transcripts will be labelled with a unique participant number and will omit any identifiable data either identifying the participant or their general practice.

© Copyright: The University of Oxford and Oxford University Hospitals NHS Foundation Trust 2019
